# Causal evidence that herpes zoster vaccination prevents a proportion of dementia cases

**DOI:** 10.1101/2023.05.23.23290253

**Authors:** Markus Eyting, Min Xie, Simon Heß, Simon Heß, Pascal Geldsetzer

## Abstract

The root causes of dementia are still largely unclear, and the medical community lacks highly effective preventive and therapeutic pharmaceutical agents for dementia despite large investments into their development. There is growing interest in the question if infectious agents play a role in the development of dementia, with herpesviruses attracting particular attention. To provide causal as opposed to merely correlational evidence on this question, we take advantage of the fact that in Wales eligibility for the herpes zoster vaccine (Zostavax) for shingles prevention was determined based on an individual’s exact date of birth. Those born before September 2 1933 were ineligible and remained ineligible for life, while those born on or after September 2 1933 were eligible to receive the vaccine. By using country-wide data on all vaccinations received, primary and secondary care encounters, death certificates, and patients’ date of birth in weeks, we first show that the percentage of adults who received the vaccine increased from 0.01% among patients who were merely one week too old to be eligible, to 47.2% among those who were just one week younger. Apart from this large difference in the probability of ever receiving the herpes zoster vaccine, there is no plausible reason why those born just one week prior to September 2 1933 should differ systematically from those born one week later. We demonstrate this empirically by showing that there were no systematic differences (e.g., in pre-existing conditions or uptake of other preventive interventions) between adults across the date-of-birth eligibility cutoff, and that there were no other interventions that used the exact same date-of-birth eligibility cutoff as was used for the herpes zoster vaccine program. This unique natural randomization, thus, allows for robust causal, rather than correlational, effect estimation. We first replicate the vaccine’s known effect from clinical trials of reducing the occurrence of shingles. We then show that receiving the herpes zoster vaccine reduced the probability of a new dementia diagnosis over a follow-up period of seven years by 3.5 percentage points (95% CI: 0.6 – 7.1, p=0.019), corresponding to a 19.9% relative reduction in the occurrence of dementia. Besides preventing shingles and dementia, the herpes zoster vaccine had no effects on any other common causes of morbidity and mortality. In exploratory analyses, we find that the protective effects from the vaccine for dementia are far stronger among women than men. Randomized trials are needed to determine the optimal population groups and time interval for administration of the herpes zoster vaccine to prevent or delay dementia, as well as to quantify the magnitude of the causal effect when more precise measures of cognition are used. Our findings strongly suggest an important role of the varicella zoster virus in the etiology of dementia.

## Main

Despite decades of large-scale investments into research on dementia^1^, including hundreds of failed phase 2 and phase 3 clinical trials of pharmaceutical agents for the prevention or treatment of dementia^2, 3^, the root causes of dementia still remain largely unclear^4^. Recently, there has been growing scientific recognition that viruses may play a role in the pathogenesis of dementia^5–7^. Different lines of evidence^8^, including the observation that herpesviruses can seed β-amyloid – a hallmark of Alzheimer’s dementia – in mice^9^, suggest a possible role for herpesviruses in particular in the pathogenesis of dementia. Currently, the US National Institute on Aging is funding a phase 2 proof-of-concept trial to test the effect of an antiviral drug on cognitive and functional ability among patients with mild Alzheimer’s dementia^10^. A second, different, approach to antiviral drugs for targeting herpesviruses is vaccination.

To date, studies in cohort and electronic health record data on the relationship between vaccination receipt (with most studies focusing on influenza vaccination^11^) and dementia have simply compared the occurrence of dementia among those who received a given vaccination versus those who did not. These studies have to assume that all characteristics that differentiate those who are vaccinated from those who are not (and that are also related to dementia) have been perfectly measured and modelled in the analysis, such that no unmeasured factors confound the relationship between vaccination receipt and dementia^12^. This assumption is usually implausible because it has to be assumed that the study perfectly measured factors that are difficult to measure, such as personal motivation or health literacy. It is also an assumption that cannot be empirically verified. Strong indications that these studies suffer from significant confounding is that i) vaccination receipt in these studies is not only associated with dementia but also a host of other health outcomes that are unlikely to be due to the vaccine^13^; ii) the direction and magnitude of the association of dementia with vaccination receipt is highly dependent on the precise analytical specifications^14^; iii) the reported magnitude of association is frequently implausibly large^15–20;^ and iv) for each existing vaccination given in adulthood, studies (often with conflicting evidence of harm or benefit^14^) exist that report an association between receiving the vaccination and dementia^11^.

We employ a fundamentally different approach, called regression discontinuity^21–24^, that takes advantage of the fact that eligibility for the herpes zoster vaccine in Wales was determined based on the exact date of birth of individuals. That is, starting on September 1 2013, those born on or after September 2 1933 were eligible for the vaccine while those born earlier never became eligible^25^ (see Methods for details). By using a rich country-wide dataset that combines information on vaccinations received, all primary and secondary care encounters, as well as death certificates, and that contains patients’ date of birth in weeks, we are able to compare adults who were ineligible for the vaccine because they were born one week before the eligibility cutoff date with those who were born one week later. There is no plausible reason why those born one week before September 2 1933 would systematically differ from those who are born just one week later, as long as September 2 1933 is not used as the date-of-birth eligibility cutoff for other interventions (e.g., another vaccination program or an educational policy) that affect the occurrence of dementia. We provide empirical evidence that no such other interventions exist. In exploiting this unique quasi-experimental setting, we are able to establish the *causal* effect (rather than merely an association) of herpes zoster vaccination on the occurrence of dementia.

We find that adults born one week after the September 2 1933 date-of-birth eligibility cutoff had a 47.2 percentage point higher probability (from 0.01% to 47.2%) of ever receiving the herpes zoster vaccine than those born just one week earlier. We then use this “natural randomization” in a regression discontinuity analysis to first replicate the known finding from clinical trials that receiving the herpes zoster vaccine reduces new diagnoses of shingles. Second, we extend this approach to an outcome – dementia – that was never assessed in clinical trials of the herpes zoster vaccine, and find that receiving the vaccine causes an approximately one-fifth reduction in the probability of a new dementia diagnosis over a seven-year follow-up period. Third, to further substantiate that our findings are not driven by confounding, we show that receiving the herpes zoster vaccine only reduced the occurrence of dementia but not of any other common causes of mortality or morbidity. Similarly, we show that receipt of the herpes zoster vaccine did not lead to increased uptake of other vaccinations or preventive health measures. Fourth, we provide empirical evidence that no other intervention (e.g., health insurance eligibility) in Wales used the identical date of birth (September 2 1933) as eligibility cutoff as was used to define eligibility for the herpes zoster vaccine. Finally, we show in exploratory analyses that the vaccine’s protective effects are far stronger among women than men for all-cause dementia and Alzheimer’s disease, while there was no significant effect heterogeneity by gender for vascular dementia. Our study focuses on the live attenuated herpes zoster vaccine (Zostavax; henceforth simply referred to as “zoster vaccine”) because the newer recombinant subunit zoster vaccine (Shingrix) became available in the UK only after our follow-up period ended^26^.

### A large difference in zoster vaccination receipt because of a mere one-week difference in age

We used the Secure Anonymised Information Linkage (SAIL) Databank^27, 28^, which contains detailed country-wide electronic health record data on primary care visits, as well as records of secondary care, in Wales linked to the country’s death register data. The study population for our primary analyses consisted of all adults born between September 1 1925 and August 31 1942 who were registered with a primary care provider (which is the case for over 98% of adults residing in Wales^29^) at the time of the start of the zoster vaccine program in Wales (on September 1 2013). For all analyses (except those with shingles and postherpetic neuralgia as outcomes), we excluded the 13,783 individuals who had already received a diagnosis of dementia prior to September 1 2013. Basic sociodemographic and clinical characteristics of the resulting sample of 282,541 adults in our primary analysis cohort are shown in **Supplement Table S1**. The Methods section provides more details on how we defined the study population for our analyses.

In Wales, individuals born between September 2 1933 and September 1 1934 (16,595 adults in our data) became eligible for the zoster vaccine on September 1 2013. Eligibility was then progressively extended to younger, but not older, age cohorts on an annual basis based on their exact date of birth (details are provided in the Methods section). In the section “Analyses demonstrating that the zoster vaccine’s effects on dementia are causal”, we provide detailed evidence against the remote possibility that the date of birth of September 2 1933 was used as the eligibility threshold for any other interventions that affect dementia risk than the zoster vaccine program. We find that being born just one week after September 2 1933, and thus being eligible for the zoster vaccine, caused an abrupt increase in the probability of ever receiving the zoster vaccine from 0.01% to 47.2% (p<0.001; **Fig. 1**). This provides a unique opportunity to determine the causal effects of the zoster vaccine because it is by virtue of the design of the vaccination program rollout implausible that individuals just around the date-of-birth eligibility threshold systematically differ from each other by anything but a one-week difference in age and a large difference in the probability of receiving the zoster vaccine. We substantiate this empirically by showing that neither the prevalence of common health outcomes (including having been diagnosed with dementia prior to the vaccination program rollout) nor the prevalence of preventive behaviors (other than zoster vaccine uptake) display a discontinuity at the date-of-birth eligibility threshold for the zoster vaccine (**Fig. 1** and **Supplement Fig. S1** and **S2**). Thus, just like in a randomized clinical trial, the two study groups (one with a low and one with a high probability of receiving the zoster vaccine) are exchangeable with each other on all observed and *unobserved* potential confounding variables^18–20^. Of note, our approach does *not* compare individuals who were eligible for the vaccine and received the vaccine with those who were eligible and did not receive the vaccine. Thus, the fact that not all those who were eligible received a zoster vaccination does not bias our analysis.

**Fig. 1:**
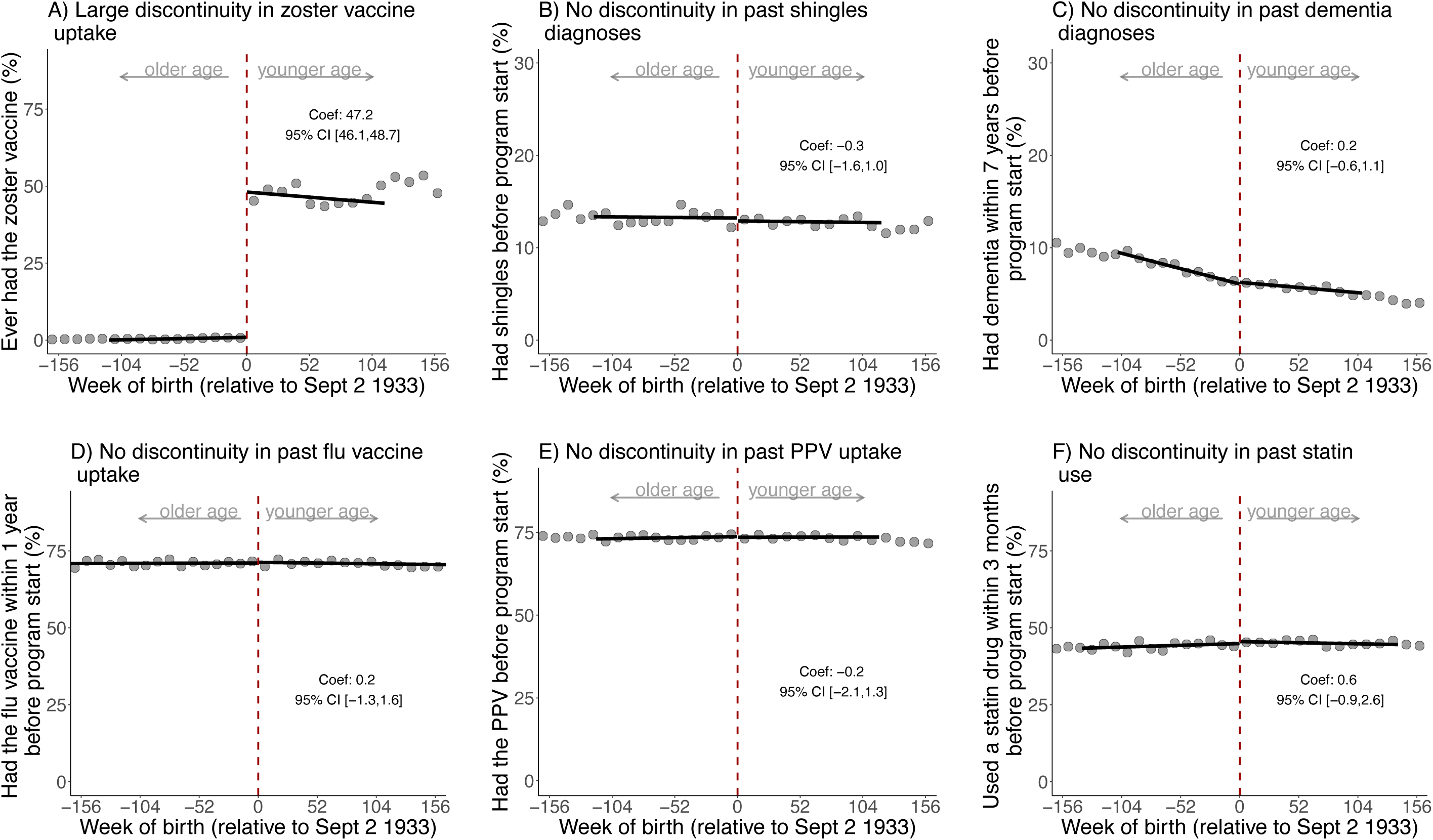
The date-of-birth eligibility cutoff led to a large discontinuity in zoster vaccine receipt but there is baseline exchangeability across the cutoff for uptake of other preventive interventions as well as past shingles and dementia diagnoses.^1, 2^ ^1^ All analyses were run on the same sample as those for the effect of the zoster vaccine on dementia occurrence. The exception is Panel C for which we did not exclude individuals with a diagnosis of dementia prior to the start of the zoster vaccine program. ^2^ Grey dots show the mean value for each 10-week increment in week of birth. Abbreviations: PPV=pneumococcal polysaccharide vaccine

### Replicating the known causal effect that zoster vaccination prevents shingles

Before we use our approach to determine the effect of the zoster vaccine on an outcome never studied in clinical trials of the vaccine, we first demonstrate that this approach successfully reproduces the known causal effect from trials that the vaccine reduces the occurrence of shingles^30^. Specifically, using a regression discontinuity design (a well-established approach for causal inference^18–20)^, we compared the occurrence of shingles between adults close to either side of the date-of-birth eligibility threshold for the zoster vaccine. In line with the approach used by clinical trials of the zoster vaccine^30^, our outcome was whether or not an individual had at least one shingles diagnosis during the follow-up period. During our follow-up period of seven years, 14,465 (among 296,324) adults had at least one diagnosis of shingles. Over the same follow-up time, we find that being eligible for the vaccine reduced the probability of having at least one shingles diagnosis by 1.0 (95% CI: 0.2 – 1.7; p=0.010) percentage points (**Fig. 2**, Panel A). Scaled by the magnitude of the jump in the probability of ever receiving the zoster vaccine at the date-of-birth eligibility threshold (i.e., taking into account that not all those who were eligible took up the vaccine), we find that receiving the zoster vaccine reduced the probability of having at least one shingles diagnosis by 2.3 (95% CI: 0.5 – 3.9; p=0.011) percentage points over the seven-year follow-up period (**Fig. 2**, Panel B). We show that our estimated effect is neither sensitive to the chosen functional form of the regression used to model the relationship of shingles occurrence with week of birth (**Supplement Fig. S3**), the width of the week-of-birth window (“bandwidth”) drawn around the date-of-birth eligibility cutoff (**Supplement Fig. S4**, Panel A), nor to different grace periods (**Fig. 2**, Panel C). With “grace periods” we refer to time periods since the index date after which follow-up time is considered to begin (see Methods for details) to allow for the time needed for a full immune response to develop after vaccine administration. There was also strong indication that the zoster vaccine reduced the probability of having at least one diagnosis of postherpetic neuralgia, although this effect did not reach statistical significance in all specifications (**Supplement Fig. S5**).

**Fig. 2:**
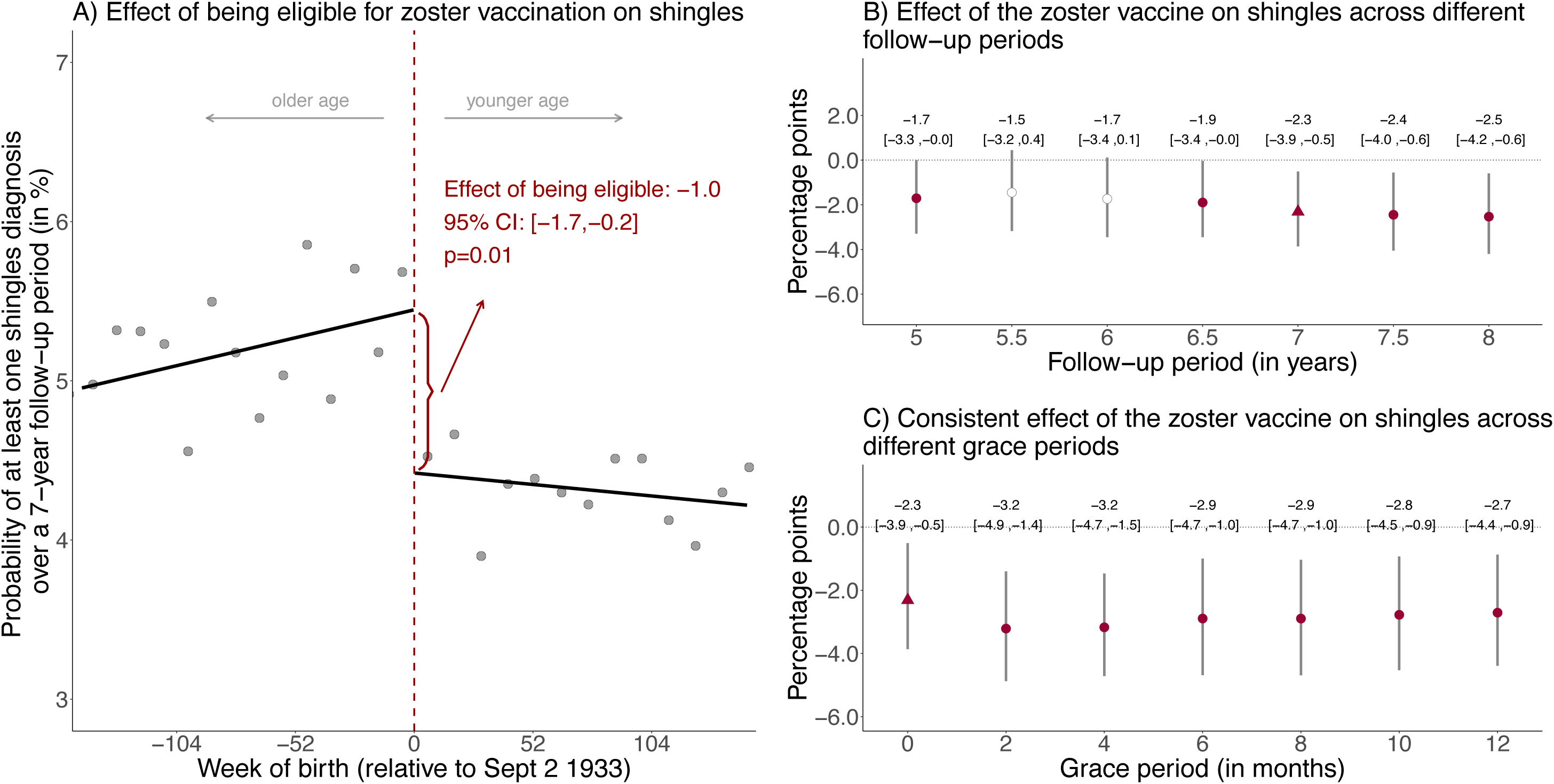
Effect estimates of being eligible (A) and having received the zoster vaccine (B and C) on the probability of having at least one shingles diagnosis during the follow-up period.^1–4,5^ ^1^ Triangles (rather than points) depict our primary specification. ^2^ Red (as opposed to white) fillings denote statistical significance (p<0.05). ^3^ With “grace periods” we refer to time periods since the index date after which follow-up time is considered to begin to allow for the time needed for a full immune response to develop after vaccine administration. ^4^ Grey vertical bars depict 95% confidence intervals. ^5^ Grey dots show the mean value for each 10-week increment in week of birth.

### Zoster vaccination reduces new diagnoses of dementia

Given the neuropathological overlap between dementia types and the difficulty in distinguishing dementia types clinically^31–33^, we defined dementia as dementia of any type or cause in our primary analyses. In exploratory analyses, we analyzed the effect of the zoster vaccine separately for vascular dementia, Alzheimer’s disease, and dementia of unspecified type. We considered an individual to have developed dementia if there was a new diagnosis of dementia in our electronic health record data (which includes all diagnoses made in primary or secondary care) or dementia was listed as a primary or contributory cause of death on the death certificate. The Read and ICD-10-codes used to define dementia are listed in **Supplement Materials**. When using a seven-year follow-up period (ending the follow-up period just prior to the COVID-19 pandemic), 35,307 adults in our sample developed dementia, which compares to 40,063 adults when using our maximum follow-up period of eight years.

Using our regression discontinuity approach, we find that being eligible for the zoster vaccine caused a 1.3 (95% CI: 0.2 – 2.7; p=0.022) percentage point absolute, and 8.5% relative, reduction in the probability of a new dementia diagnosis over our seven-year follow-up period (**Fig. 3**, Panel A). Scaled to account for the fact that not all those who were eligible received the vaccine, we find that actually receiving the zoster vaccine reduced the probability of a new dementia diagnosis by 3.5 (95% CI: 0.6 – 7.1; p=0.019) percentage points, corresponding to a relative reduction of 19.9%. Examining the magnitude of the *absolute* effect over different follow-up periods ranging from four to eight years, we find no indication that the effectiveness of the vaccine for reducing the probability of a new dementia diagnosis wanes over time (**Fig. 3**, Panel B). However, given that the proportion of patients who received a new diagnosis of dementia increased over time as the follow-up period lengthened, the *relative* effect of the vaccine on the probability of receiving a new dementia diagnosis did decrease over time, from 22.5% after five years to 19.9% after seven years and 17.0% after eight years of follow-up. The effect estimates were generally not sensitive to different grace periods (**Fig. 3**, Panel C), the functional form of our regressions (**Supplement Fig. S6**), nor the width of the week-of-birth window (“bandwidth”) drawn around the date-of-birth eligibility cutoff (**Supplement Fig. S4**, Panel B).

**Fig. 3:**
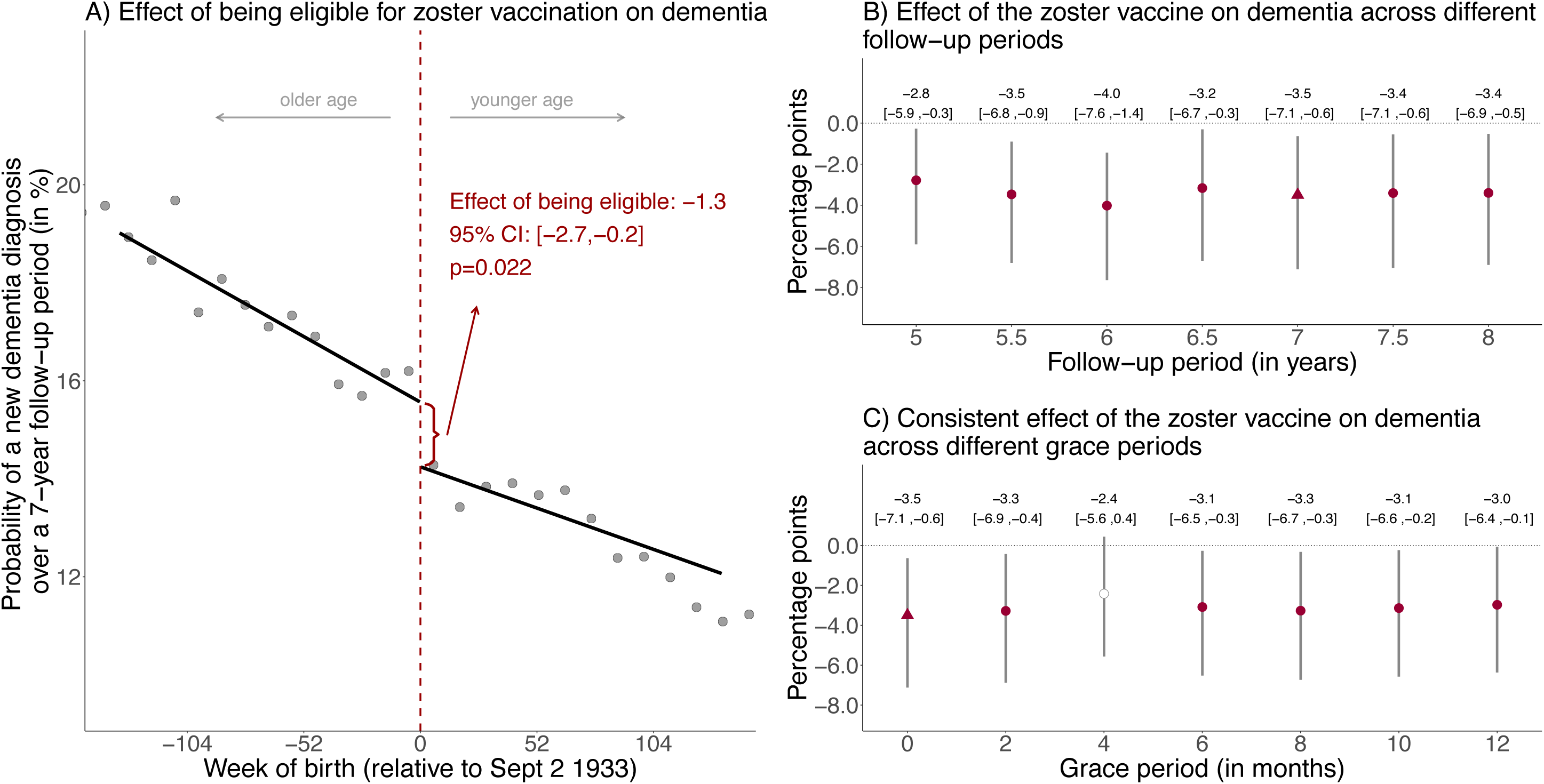
Effect estimates of being eligible (A) and having received the zoster vaccine (B and C) on new diagnoses of dementia.^1,2,3,4,5^ ^1^ Triangles (rather than points) depict our primary specification. ^2^ Red (as opposed to white) fillings denote statistical significance (p<0.05). ^3^ With “grace periods” we refer to time periods since the index date after which follow-up time is considered to begin to allow for the time needed for a full immune response to develop after vaccine administration. ^4^ Grey vertical bars depict 95% confidence intervals. ^5^ Grey dots show the mean value for each 10-week increment in week of birth.

### Analyses demonstrating that the zoster vaccine’s effects on dementia are causal

We conducted a series of analyses to confirm that our regression discontinuity approach indeed yields unbiased causal effects. A confounding factor in our study must be an intervention that used the identical date-of-birth cutoff (September 2 1933) as eligibility criterion as the zoster vaccine program. Such an intervention is unlikely to only affect the risk of developing dementia without also influencing other health outcomes. Thus, one confirmatory type of analysis is to demonstrate that the observed effects of the vaccine are specific to dementia. We, therefore, implemented the same regression discontinuity approach as we have done for shingles and dementia for the ten leading causes of disability-adjusted life years (a composite measure of morbidity and premature mortality^34^) and mortality for the age group 70+ years in Wales in 2019^35^. We show that being eligible for the zoster vaccine did not have an effect on any of these common health outcomes (**Supplement Fig. S7**). The Read and ICD-10-codes for each of these diagnoses are provided in **Supplement Materials**.

To more definitively rule out that another intervention (e.g., a different vaccination program) that affects the risk of developing dementia used exactly the same date-of-birth eligibility cutoff as was used for zoster vaccine eligibility, we undertook three additional types of analysis. First, we demonstrate that the September 2 1933 date-of-birth threshold does not affect the probability of taking up other preventive health interventions, including receiving the influenza vaccine, using a statin or antihypertensive drug, or being screened for breast cancer during our follow-up period (**Supplement Fig. S8**). Similarly, as shown in **Fig. 1** and **Supplement Fig. S2**, there was no difference in the probability of uptake of these preventive health interventions between participants around the September 2 1933 date-of-birth eligibility threshold prior to the start of the zoster vaccine program. Second, we verified that the day-month (i.e., September 2) cutoff used for zoster vaccine eligibility was not also used for other interventions that affect dementia risk. We did so by implementing the identical analysis as for September 1 2013 (the actual date on which the zoster vaccine program started) for September 1 of each of the three years prior to, and after, 2013. Thus, for instance, when shifting the start date of the program to September 1 2012, we compared those around the September 2 1932 eligibility threshold with the follow-up period starting on September 1 2012. To be able to do so while using the same length of follow-up for all comparisons, we had to reduce the follow-up period to five years for this robustness check. As an additional check that allowed us to maintain the length of the seven-year follow-up period used in our primary analyses, we shifted the program start date to September 1 of each of the six years preceding (but not after) 2013. As expected, for both of these checks we only find a significant effect on dementia occurrence for September 1 of 2013 (**Supplement Fig. S9** and **S10**). Third, we carried out the identical age-cohort comparison (that is, comparing cohorts just around the September 2 1933 date-of-birth cutoff) as we do in our primary regression discontinuity analysis, except that we started the follow-up period seven years earlier (on September 1 2006) and ended the follow-up period on August 31 2013 (i.e., before the first group became eligible for the zoster vaccine). In this way, we implemented the same age-cohort comparison and have the same duration of follow-up (without overlapping with the period during which one age cohort was eligible for the zoster vaccine while the other was not) as in our primary analysis. This specification tests our identifying assumption that there were no pre-existing differences in dementia. We find that there is no difference in the seven-year incidence of dementia between age cohorts around the September 2 1933 date-of-birth threshold for the seven-year period prior to the zoster vaccine rollout (**Supplement Fig. S11**). Taken together, these analyses are strong evidence against the possibility that, in theory, the exact day-month-year combination (September 2 1933) that was used as the date-of-birth eligibility threshold for the zoster vaccine rollout could have also been used by another relevant intervention or policy in the past.

### Effect heterogeneity by dementia type and gender

Next, as an exploratory analysis, we examined whether the effect of the zoster vaccine differs by type of dementia. For this analysis, we focused on the three types of dementia recorded in our data: vascular dementia, Alzheimer’s disease, and dementia of unspecified type. Of the 35,307 individuals who were diagnosed with dementia during our seven-year follow-up period, 11,247 were diagnosed with vascular dementia, 14,481 with Alzheimer’s disease, and 12,000 with dementia of unspecified type. 2,421 individuals were diagnosed with both Alzheimer’s disease and vascular dementia. Because shingles occurs more commonly among women than men^36, 37^, and the growing evidence that the pathogenesis of dementia, particularly for Alzheimer’s disease, may differ in important aspects by sex^38–40^, we also investigated whether i) our estimates for the effect of the zoster vaccine on dementia differ significantly between women and men, and ii) any such effect heterogeneity by gender was stronger for Alzheimer’s disease than for vascular dementia and dementia of unspecified type.

Given that some types of dementia in our data are more common than others, we would expect that the absolute effect magnitudes (and their corresponding level of statistical significance) differ by type of dementia. As such, the *relative* effect sizes by dementia type are more informative. We find that the relative effect sizes (Alzheimer’s disease: 17.9%, dementia of unspecified type: 19.1%, and vascular dementia: 18.8%) are similar across the three types of dementia (**Supplement Table S2**). These findings, however, are merely suggestive because there is likely substantial misclassification and overlap between dementia types in our data.

The vaccine’s effect on new diagnoses of dementia was markedly greater among women than men (**Fig. 4** and **Supplement Table S3**, Column 1). In fact, among men, the point estimates were close to zero across all specifications. Nonetheless, the 95% confidence interval for the effect among men included the possibility of a relative protective effect on dementia over a seven-year follow-up period of up to 23.9%. We can, therefore, not exclude the possibility that the vaccine also had a protective effect on dementia among men. When examining the effect heterogeneity by gender separately for each type of dementia, we found that the protective effect of the vaccine for dementia was significantly stronger for women than men for Alzheimer’s disease (p=0.018), but not for vascular dementia (p=0.376) and dementia of unspecified type (p=0.358) (**Supplement Table S3**, Columns 2-4). The magnitude of the jump in vaccine uptake at the September 2 1933 date-of-birth eligibility threshold was similar between men and women (**Supplement Fig. S12**). Likewise, there was no significant difference between men and women in the effect of the zoster vaccine on diagnoses of shingles and postherpetic neuralgia (**Supplement Table S3**, Columns 5-6).

**Fig. 4:**
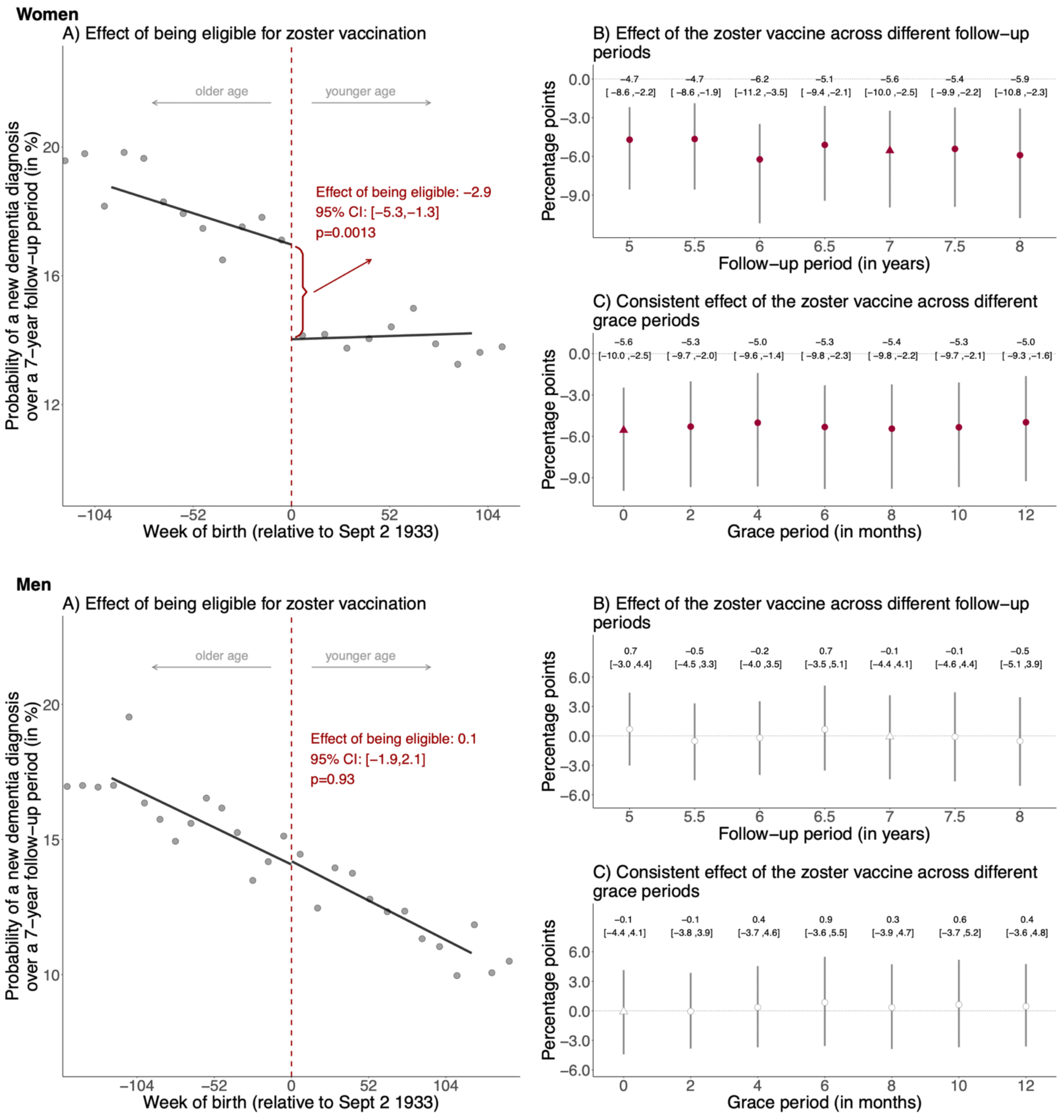
Effect estimates of being eligible (A) and having received the zoster vaccine (B and C) on new diagnoses of dementia of any type, separately for women and men.^1–5^ ^1^ Triangles (rather than points) depict our primary specification. ^2^ Red (as opposed to white) fillings denote statistical significance (p<0.05). ^3^ With “grace periods” we refer to time periods since the index date after which follow-up time is considered to begin to allow for the time needed for a full immune response to develop after vaccine administration. ^4^ Grey vertical bars depict 95% confidence intervals. ^5^ Grey dots show the mean value for each 10-week increment in week of birth.

### Additional robustness checks

Further to the robustness checks already detailed in preceding sections, we conducted four additional analyses to ensure that our findings are robust to different analytical specifications. First, we show that we also find significant causal effects of the zoster vaccine on reducing dementia diagnoses if a diagnosis is defined only as dementia being listed as a primary or contributory cause of death in the death certificate (**Supplement Table S4**, Column 2), as well as when defining dementia solely as a new prescription of a medication (donepezil hydrochloride, galantamine, rivastigmine, or memantine hydrochloride) that is frequently prescribed to slow the progression of Alzheimer’s disease (**Supplement Table S4**, Column 3)^41^. Second, we implemented our analyses when restricting the analysis cohort to the 237,196 (84.0% of the analysis cohort for our primary analyses) patients who visited their primary care provider at least once a year during each of the five years preceding the start of the zoster vaccine rollout. This robustness check aims to determine the causal effect of the zoster vaccine among patients who interact frequently with the health system and may, thus, be more likely to be screened for dementia. The effect sizes among this cohort do not differ significantly from those of our primary analytical cohort (**Supplement Table S4**, Column 4). Third, while not required for unbiasedness of regression discontinuity estimates, in a separate analysis, we adjusted our regressions for indicators of health service utilization during the follow-up period.

These variables were the probability of receiving at least one influenza vaccination and the number of i) primary care visits, ii) outpatient visits, and iii) hospital admissions. The effect sizes remain similar (**Supplement Table S4**, Column 5). Fourth, we compared the effect sizes when the index date (i.e., the date at which the quasi-randomization to intervention or control group occurred) was defined as September 1 2013 for all cohorts, versus when it was defined as the date at which each cohort first became eligible for the zoster vaccine (see Methods for details). The effect sizes do not vary significantly between these two analytical choices (**Supplement Fig. S13** and **Supplement Table S4**, Column 6).

## Discussion

This study found that the zoster vaccine reduced the probability of a new dementia diagnosis by approximately one fifth over a seven-year follow-up period. By taking advantage of the fact that the unique way in which the zoster vaccine was rolled out in Wales constitutes a natural experiment, and meticulously ruling out each possible remaining source of bias, our study provides causal rather than associational evidence. Given that our effect sizes remain stable across a multitude of specifications and analysis choices, it is also improbable that our finding is a result of chance. The evidence provided by this study is, thus, fundamentally different to studies that have simply correlated (with adjustment for, or matching on, certain covariates) vaccine receipt with dementia.

Our rigorous causal approach allows for the conclusion that herpes zoster vaccination is very likely an effective means of preventing or delaying the onset of dementia. Our substantial effect sizes, combined with the relatively low cost of the zoster vaccine^42–44^, imply that the zoster vaccine is both far more effective as well as cost-effective in preventing or delaying dementia than existing pharmaceutical interventions^45–47^. In addition, and arguably even more importantly, the finding that the zoster vaccine reduces the occurrence of dementia could help elucidate the pathogenesis of dementia, which in turn could lead to additional, and potentially even more effective, interventions.

Our findings create an imperative for research in the following areas. First, in addition to confirming our conclusions, randomized trials are needed for determining the optimal timing and frequency of zoster vaccination for dementia prevention. Our results demonstrate that zoster vaccination is effective in preventing or delaying dementia if administered in individuals’ late seventies. We were, however, unable to ascertain whether providing the vaccine at younger ages would result in additional averted dementia cases and whether there is an age above which zoster vaccination has little to no effect on dementia occurrence. In addition, given evidence that the effectiveness of the zoster vaccine for preventing shingles episodes declines over time, which is the case for both the live attenuated and, albeit less so, the recombinant subunit zoster vaccine^48–50^, it may well be the case that the optimal strategy for preventing dementia is regular ‘booster shots’ of the vaccine. Our study provides suggestive evidence that such booster shots may be required as we observed waning effectiveness (on the relative scale) over time for zoster vaccination reducing the probability of a new dementia diagnosis.

Second, our findings strongly suggest that investments into researching the role of the varicella zoster virus and the immune response to the zoster vaccine in the pathogenesis of dementia could provide critical insights into how a significant proportion of dementia cases can be prevented or effectively treated. Our study also suggests that the varicella zoster virus plays a greater role in the pathogenesis of dementia among women than men, particularly for Alzheimer’s disease. Third, research should be conducted to determine if the zoster vaccine reduces, or potentially even reverses, cognitive decline among those with mild cognitive impairment or mild-to-moderate dementia. A clinical trial is underway to test the effect of daily valacyclovir among patients with mild dementia who test positive for herpes simplex virus-1 or herpes simplex virus-2 serum antibodies on change in cognitive and functional ability over a 78-week follow-up period^10^. Similar efforts are required for the zoster vaccine.

Although validation studies on the ability of electronic health record data to reliably ascertain dementia occurrence have generally been encouraging^51–53^, our outcome ascertainment undoubtedly still suffers from some degree of under-detection both in whether and how timely dementia is diagnosed. Crucially, however, for this under-detection in dementia (as well as any false diagnoses of dementia) to bias our relative effect sizes, it would have to be the case that the degree of under-detection of dementia differed substantially between those born just before versus just after the September 2 1933 date-of-birth eligibility threshold for zoster vaccination. There are only two mechanisms through which the zoster vaccination eligibility threshold could have affected the degree to which dementia is underdiagnosed. The first mechanism is that receiving the zoster vaccine presented an opportunity for the health system to diagnose previously undetected dementia. This scenario, however, would bias our effect estimates towards the zoster vaccine *increasing* (rather than having the protective effect observed in this study) dementia occurrence, thus leading us to underestimate the protective effect of the zoster vaccine for dementia. The second mechanism is that zoster vaccination reduced healthcare utilization for shingles episodes, which translated to fewer opportunities for the health system to diagnose dementia. This mechanism cannot plausibly be of sufficient magnitude to significantly bias our findings because both the size of our effect estimates and the occurrence of new diagnoses during the follow-up period were substantially smaller for our shingles than our dementia outcome. Thus, even under the most extreme assumption that every adult in our study population without a dementia diagnosis had undetected dementia throughout the entire follow-up period and that healthcare utilization for shingles episodes presented a certain way for the health system to diagnose undetected dementia, this bias would merely account for a small fraction of the effect of the zoster vaccine on reducing dementia. Providing further reassurance, adjusting our regressions for the frequency of health service utilization (the number of primary care visits, outpatient visits, hospital admissions, and influenza vaccinations received) during the follow-up period did not substantially change our effect estimates.

Our study has several additional limitations. First, we were limited to a maximum follow-up period of adults of eight years. Our study can therefore not inform on the effectiveness of the zoster vaccine for reducing dementia occurrence beyond this time period. Second, we are unable to provide estimates for the effectiveness of the zoster vaccine for reducing dementia occurrence in age groups other than those who were weighted most heavily in our regression discontinuity analyses (primarily those aged 79 to 80 years). Third, the COVID-19 pandemic likely affected the timeliness with which dementia was diagnosed. However, the follow-up period used in our primary analyses ended prior to the start of the COVID-19 pandemic. In addition, because the pandemic affected those born just before versus just after September 2 1933 equally, pandemic-related under-detection of dementia does not bias our relative effect estimates; it may merely have reduced the magnitude of our effect estimates on an absolute scale (and only for the analyses that used a follow-up period of eight years). Fourth, our comparison of effect sizes between dementia types is limited by the difficulty of classifying dementia into types clinically, as evidenced by the fact that around a third of new dementia diagnoses over our seven-year follow-up period were labelled as dementia of unspecified type. Fifth, because the newer recombinant subunit zoster vaccine (“Shingrix”) only became available in the UK in September 2021^26^, which is after our follow-up period ended, our effect estimates apply to Zostavax only.

**Data availability:** The data that support the findings of this study are available from the SAIL Databank^27^. Researchers must request access to the data directly from SAIL. The authors have no permission to share the data.

**Code availability:** All Read and ICD-10 codes to define variables are available in the Supplement. All statistical analysis code (in R) will be made available in a publicly accessible GitHub repository upon acceptance of the manuscript for publication.

## Methods

### Description of the zoster vaccine rollout in Wales

The live attenuated zoster vaccine (Zostavax) was made available to eligible individuals in Wales through a staggered rollout system starting on September 1 2013. Under this system, individuals aged 71 years or older were categorized into three groups on September 1 of each year: i) an ineligible cohort of those aged 71 to 78 years (or 77 years, depending on the year of the program), who would expect to become eligible in the future; ii) a catch-up cohort, consisting of individuals aged 79 years (or 78 years, again depending on the year of the program); and iii) those who were ineligible as they were aged 80 years or older and who would never again become eligible.

Our analysis focused on adults born between September 1 1925 (88 years old at program start) and September 1 1942 (71 years old at program start). Those born between September 1 1925 and September 1 1933 never became eligible, whereas those born between September 2 1933 and September 1 1942 became progressively eligible in a catch-up cohort. Specifically, the vaccine was offered to those born between September 2 1933 and September 1 1934 in the first year of the program (September 1 2013 to August 31 2014); those born between September 2 1934 and September 1 1936 in the second year (September 1 2014 to August 31 2015); those born between September 2 1936 and September 1 1937 in the third year (September 1 2015 to August 31 2016); and those born between September 2 1937 and September 1 1938 in the fourth year (September 1 2016 to August 31 2017). As of April 1 2017, individuals become eligible for the vaccine on their 78^th^ birthday and remain eligible until their 80^th^ birthday. Our analysis principally compared individuals born on or shortly after September 2 1933, to individuals who never became eligible as they were born shortly before September 2 1933.

### Data source

Healthcare in Wales is provided through the Welsh National Health Service (NHS), which is part of the United Kingdom’s single-payer single-provider healthcare system^54^. NHS Wales and the Welsh government have partnered up with Swansea University to create the Secure Anonymised Information Linkage (SAIL) Databank^27, 28^. The SAIL databank includes full electronic health record data for primary care visits in Wales linked to information on hospital-based care as well as the country’s death register data.

SAIL generates a list of all individuals who have ever been registered with a primary care provider in Wales (which is the case for over 98% of adults residing in Wales^29^) from the Welsh Demographic Service Dataset^55^. This dataset also contains individuals’ unique anonymized NHS number, date of birth, anonymized address, primary care provider registration history, as well as the Welsh Index of Multiple Deprivation (the official measure of relative deprivation for small areas in Wales^56^). SAIL then links this universe of individuals to each of the following datasets. Electronic health record data from primary care providers is made available in SAIL through the Welsh Longitudinal General Practice dataset^57^, which contains data from approximately 80% of primary care practices in Wales and 83% of the Welsh population. These electronic health record data use Read codes, which provide detailed information on patients and their care encounters, including diagnoses, clinical signs and observations, symptoms, laboratory tests and results, procedures performed, and administrative items^58^. As specialist care in the NHS is only provided based on a referral from the patient’s primary care provider (i.e., primary care providers are the “gate-keepers” to the wider health system)^54^, referrals to, and diagnoses made in, specialist care are also recorded in the primary care electronic health record data. Additionally, diagnoses made and procedures performed in the hospital setting (as part of inpatient admissions or day-case procedures) are provided in SAIL through linkage to the Patient Episode Database for Wales^59^, which begins in 1991 and contains data for all hospital-based care in Wales as well as hospital-based care provided in England to Welsh residents. Procedures are encoded using OPCS-4 codes^60^ and diagnoses using ICD-10 codes^61^.

Attendance information at any NHS Wales hospital outpatient department is provided through linkage to the Outpatient Database for Wales^62^, which starts in 2004. ICD-10 encoded diagnoses of cancers are identified through linkage to the Welsh Cancer Intelligence and Surveillance Unit^63^, which is the national cancer registry for Wales that records all cancer diagnoses provided to Welsh residents wherever they were diagnosed or treated. This dataset begins in 1994. Finally, cause-of-death data is provided for all Welsh residents (regardless of where they died in the United Kingdom) through linkage to the Annual District Death Extract^64^, which begins in 1996 and includes primary and contributory causes of death from death certificates. Cause-of-death data uses ICD-9 coding until 2001 and ICD-10 coding thereafter.

### Study cohort, follow-up period, and loss to follow-up

Our study population consisted of 296,603 individuals born between September 1 1925 and September 1 1942 who were registered with a primary care provider (which is the case for more than 98% of adults residing in Wales^29^) in Wales on the start date of the zoster vaccine program rollout (September 1 2013). Since we only had access to the date of the Monday of the week in which an individual was born, we were unable to determine whether the individuals born in the cutoff week starting on August 28 1933 were eligible for the zoster vaccine in the first year of its rollout. Therefore, we excluded 279 individuals born in this particular week. Among the remaining individuals, 13,783 had a diagnosis of dementia prior to September 1 2013 and were, thus, excluded from the analyses with dementia occurrence as outcome. The size of our final analysis cohort for all primary analyses for dementia occurrence was, therefore, 282,541. This analysis cohort was also used for all analyses with uptake of other preventive health interventions as well as the leading causes of disability-adjusted life years (DALYs) and mortality in Wales as outcome (henceforth referred to as negative outcome control analyses and detailed under the “Statistical analysis” section below). Our analyses with episodes of shingles and postherpetic neuralgia as outcomes used the same study cohort except that we did not exclude individuals with a dementia diagnosis prior to September 1 2013. With 296,324 individuals, the size of this study cohort was, thus, larger.

We followed these individuals from September 1 2013 to August 31 2021, which allowed for a maximum follow-up period of eight years. In our primary specification, we selected a follow-up period of seven years (i.e., until August 31 2020) because this allowed us to include grace periods of up to 12 months whilst still keeping the follow-up period constant for individuals on either side of the date-of-birth eligibility cutoff. We, however, also show all results for follow-up periods of 5.0, 5.5, 6.0, 6.5, 7.0, 7.5, and 8.0 years. Owing to the unique anonymized NHS number assigned to each patient, we were able to follow individuals across time even if they changed primary care provider. Patients were, thus, only lost to follow-up in our cohort if they emigrated out of Wales or changed to one of the approximately 20% of primary care practices in Wales that did not contribute data to SAIL. Over our seven-year follow-up period, this was the case for 23,049 (8.2%) of adults in our primary analysis cohort, with no significant difference in this proportion between those born just before versus just after the September 2 1933 eligibility threshold.

### Definition of outcomes

Dementia was defined as dementia being named as a primary or contributory cause of death in the death certificate, or a diagnosis of dementia made either in primary care (as recorded in the primary care electronic health record data), specialist care, or hospital-based care. The date of the first recording of dementia across any of these data sources was used to define the date on which the patient was diagnosed with dementia. Because of the neuropathological overlap between dementia types and difficulty in distinguishing dementia types clinically^31–33^, we chose to define dementia as dementia of any type or cause in our primary analyses. In exploratory analyses, we examined the effect of the zoster vaccine separately for vascular dementia, Alzheimer’s disease, and dementia of unspecified type. Because of evidence that the pathogenesis of Alzheimer’s disease may vary by sex^38–40^, we additionally implemented these analyses separately for women and men.

Shingles and postherpetic neuralgia were similarly defined as a diagnosis of shingles or postherpetic neuralgia made in primary or hospital-based care. Again, the date of the first recording of a diagnosis of shingles or postherpetic neuralgia across any of these data sources was used to define the date on which the outcome occurred. The Read and ICD-10 codes used to define dementia, each type of dementia, shingles, postherpetic neuralgia, and each negative control outcome are detailed in the Supplement (**Supplement Materials**).

### Statistical analysis

The two authors who analyzed the data (M.E. and M.X.) have coded all parts of the analysis independently. Occasional minor differences, resulting from different data coding choices, were resolved through discussion.

#### Our regression discontinuity approach

Our statistical approach exploits the fact that, unless another intervention uses the exact same date-of-birth eligibility threshold (September 2 1933) as the zoster vaccine rollout, those who were born just after the date-of-birth eligibility threshold must be exchangeable (i.e., comparable in observable and unobservable characteristics) with those born just before except for being ineligible for the zoster vaccine. We used a regression discontinuity design to analyze our data, which is a well-established method for causal inference in the social sciences^65^. Regression discontinuity analysis estimates expected outcome probabilities just left and just right of the cutoff, to obtain an estimate of the treatment effect. Based on current best practice for regression discontinuity analyses^66^, we used local linear triangular kernel regressions (assigning a higher weight to observations lying closer to the date-of-birth eligibility threshold) in our primary analyses and quadratic polynomials in robustness checks. An important choice in regression discontinuity analyses is the width of the data window (the “bandwidth”) that is drawn around the threshold. Following standard practice, we used a mean squared error (MSE)-optimal bandwidth^67^, which minimizes the mean squared errors of the regression fit, in our primary analyses. In robustness checks, we examined the degree to which our point estimates vary across different bandwidth choices ranging from 0.25 times to two times the MSE-optimal bandwidth. We used robust bias-corrected standard errors for inference^68^.

#### Estimating the effect of being eligible for the zoster vaccine

In the first step, we determined the effect of being eligible for the zoster vaccine (regardless of whether the individual actually received the vaccine) on our outcomes. To do so, we estimated the following regression equation:

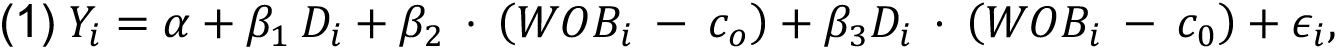

where 𝑌*_i_* is a binary variable equal to one if an individual experienced the outcome (e.g., shingles or dementia). The binary variable 𝐷*_i_* indicates eligibility for the zoster vaccine and is equal to one if an individual was born on or after the cutoff date of September 2 1933. The term (𝑊𝑂𝐵*_i_* – 𝑐_0_) indicates an individual’s week of birth centered around the cutoff date. The interaction term 𝐷*_i_* ⋅ (𝑊𝑂𝐵*_i_* – 𝑐_0_) allows for the slope of the regression line to differ on either side of the threshold. The parameter 𝛽_1_ identifies the absolute effect of being eligible for the vaccine on the outcome. Wherever we report relative effects, we calculated these by dividing the absolute effect estimate 𝛽_1_ by the mean outcome just left of the date-of-birth eligibility threshold, i.e., the estimate of 𝛼.

#### Estimating the effect of actually receiving the zoster vaccine

In the second step, we estimated the effect of actually receiving the zoster vaccine on our outcomes. This effect is commonly referred to as the complier average causal effect (CACE) in the econometrics literature^21^. As is standard practice^21^, we used a so-called fuzzy regression discontinuity design to estimate the CACE. Fuzzy regression discontinuity analysis takes into account the fact that the vaccine is not deterministically assigned at the week-of-birth cutoff. Instead, a proportion of ineligible individuals still received the vaccine and a proportion of eligible individuals did not receive the vaccine. To account for this fuzziness in the assignment, the fuzzy regression discontinuity design employs an instrumental variable approach, with the instrumental variable being the binary variable that indicates whether or not an individual was eligible to receive the vaccine, i.e., is born on or after September 2 1933. As we verify in our plot of vaccine receipt by week of birth (**Fig. 1**, Panel A), individuals who were born just after the date-of-birth eligibility threshold had a far higher probability of receiving the zoster vaccine than those born just before the threshold. Other than the abrupt change in the probability of receiving the zoster vaccine, there is no other difference in characteristics that affect the probability of our outcomes occurring between those born just after versus just before the date-of-birth eligibility threshold. Thus, the indicator variable for the date-of-birth eligibility threshold is a valid instrumental variable to identify the causal effect of receipt of the zoster vaccine on our outcomes. To compare the probability of experiencing the outcome between those who actually received the zoster vaccine versus those who did not, the instrumental variable estimation scales the effect size for being eligible for the zoster vaccine by the size of the abrupt change in the probability of receiving the vaccine at the date-of-birth eligibility threshold. The size of the jump is estimated via the following first-stage regression equation:

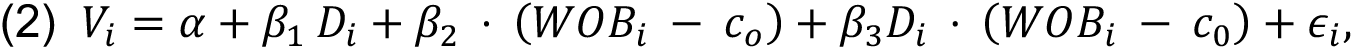

where 𝑉*_i_* is a binary variable indicating if the individual received the zoster vaccine and 𝛽_1_ identifies the discontinuous increase in vaccine receipt at the date-of-birth eligibility threshold. All other parameters are the same as in regression equation (1).

To compute relative effect sizes for the effect of actually receiving the zoster vaccine, we divided the CACE estimate obtained from the instrumental variable estimation described above by the mean outcome among unvaccinated compliers (those who do not receive the vaccine because they are not eligible) just at the threshold. Since, among the ineligible group, compliers are not distinguishable from never-takers (those who do not take the vaccine irrespective of their eligibility), their mean outcome must be estimated. To do this, we followed standard practice^23^. By construction, the mean outcome among vaccinated patients at the date-of-birth threshold is approximately equal to the population-weighted average of the mean outcomes among vaccinated compliers (those who only receive the vaccine because they are eligible) and eligible always-takers (those who would always receive the vaccine irrespective of their eligibility). This relationship can be solved for the mean outcome among vaccinated compliers because all missing unknown quantities in this relationship can be computed from our data: the mean outcome of eligible always-takers can be computed as the mean outcome among vaccinated individuals just left of the threshold; the population share of always-takers can be computed as the share of vaccinated individuals left of the threshold; and the population share of vaccinated compliers corresponds to the treatment effect of the first stage regression (equation 2 above). Finally, we subtracted the estimate for the CACE from the mean outcome among vaccinated compliers to obtain the mean outcome among unvaccinated compliers.

#### Empirical tests that the key assumption of regression discontinuity is met

The key assumption made by regression discontinuity designs is the continuity assumption^21^. In our setting, the continuity assumption is that if September 2 1933 had not been used as the date-of-birth eligibility threshold for the zoster vaccine program, then the probability of a new dementia diagnosis during our follow-up period would be identical for individuals born just before versus just after September 2 1933. Two scenarios could violate this assumption. First, the continuity assumption would be violated if week of birth in our data was recorded with systematic bias such that individuals with a differential risk of dementia are systematically more likely to be categorized to one side of the date-of-birth vaccine eligibility threshold. Given that vaccine prescribers and administrators in our data are not able to change the recorded date of birth for a patient, it is not possible that this concern is a source bias in our analysis. If this concern was a source of bias, then we would expect to see bunching in the number of patients with a week of birth just on one side of the September 2 1933 threshold. As shown in **Supplement Fig. S14**, this is not the case. We also formally tested for bunching using the McCrary density test^69^, which confirms (p=0.28) that there is no evidence of such bunching in the week-of-birth variable in our data. Second, the continuity would be violated if the exact same date-of-birth eligibility threshold (September 2 1933) as for the zoster vaccine was also used for other interventions (e.g., other vaccinations or an educational policy) that affect the probability of being diagnosed with dementia during our follow-up period. We conducted a series of robustness checks (described in detail in the next section) to verify that no such competing interventions exist.

#### Robustness checks to confirm that our findings are causal

Our analysis can only be confounded if the confounding variable changes abruptly at the September 2 1933 date-of-birth eligibility threshold such that individuals very close to either side of this threshold would no longer be exchangeable with each other. The only plausible scenario of such a confounding variable would be the existence of an intervention that used the exact same date-of-birth eligibility threshold as the zoster vaccine rollout and that also affected the probability of a dementia diagnosis during our follow-up period. We conducted three analyses to demonstrate that the existence of such an intervention is extremely unlikely, by establishing that measures of outcomes and behaviors that would be affected by such an intervention are smooth across the date-of-birth eligibility cutoff.

First, across a range of birthdates around the September 2 1933 eligibility threshold, we plotted the probability of having received the following diagnoses or interventions prior to the start of the zoster vaccine program (on September 1 2013): diagnosis of shingles, influenza vaccine receipt in the preceding 12 months, receipt of the pneumococcal vaccine as an adult, current statin use (defined as a new or repeat prescription of a statin in the three months preceding program start), current use of an antihypertensive medication (defined as a new or repeat prescription of an antihypertensive drug in the three months preceding program start), participation in breast cancer screening (defined as the proportion of women with a record of referral to, attendance at, or a report from “breast cancer screening” or mammography), and each of the top ten leading causes of disability-adjusted life years (DALYs) and mortality for Wales in 2019 as estimated by the Global Burden of Disease Project^35^. The Read codes for each of these variables are provided in **Supplement Materials**. As shown in **Fig. 1** and **Supplement Fig. S1** and **S2**, none of these variables displayed a significant jump at the September 2 1933 eligibility threshold. As is the case for balance tables in clinical trials, these plots provide reassurance that individuals close to either side of the September 2 1933 eligibility threshold are exchangeable with each other.

Second, we conducted the same analysis as we did for individuals with birthdays on either side of the September 2 1933 threshold also for people with birthdays around September 2 of each of the three years of birth preceding and succeeding 1933. For example, when moving the start date of the program to September 1 2011, we started the follow-up period on September 1 2011 and compared individuals around the September 2 1931 eligibility threshold. In order to ensure the same length of follow-up in each of these comparisons, we had to reduce the follow-up period to five years for this set of analyses. Thus, as an additional check, we only shifted the start date of the program to September 1 of each of the six years preceding (but not succeeding) 2013, which allowed us to maintain the same seven-year follow-up period as in our primary analysis. If another intervention that affects dementia risk also used the September 2 threshold to define eligibility, then we would expect to observe effects on dementia incidence for these comparisons of individuals just around the September 2 thresholds of other birth years.

As shown in **Supplement Fig. S9** and **S10**, the only year of birth for which the September 2 threshold has an effect on dementia incidence is 1933 (i.e., the year used as eligibility criterion by the zoster vaccine program).

Third, we conducted the identical comparison of individuals around the September 2 1933 date-of-birth threshold as in our primary analysis, except for starting the follow-up period seven years prior to the start of the zoster vaccine program rollout. If there was an intervention that used the September 2 1933 date-of-birth eligibility threshold but was implemented before the rollout of the zoster vaccine program, then we would expect to see an effect of the September 2 1933 threshold on dementia incidence in this analysis. As shown in **Fig. 1**, Panel C, and **Supplement Fig. S11**, there is no evidence of any such effect.

We conducted two additional analyses to further confirm that our observed effects on dementia incidence are indeed causal. First, we verified that the effects that we observed in our analyses for dementia incidence are specific to dementia. If an intervention that used the exact same date-of-birth eligibility threshold as the zoster vaccine program indeed existed, it would be unlikely to only affect dementia risk without also having an influence on other health outcomes. We, thus, conducted the same analysis as with dementia incidence as outcome but for each of the ten leading causes of DALYs and mortality in Wales in 2019 for the age group 70+ years^35^. The results of this analysis are shown in **Supplement Fig. S7** and demonstrate that the September 2 1933 threshold has no effect on any of these common health outcomes other than dementia. Second, we adjusted our regressions for the following variables as assessed during our seven-year follow-up period: the number of primary care visits, outpatient visits, hospital admissions, and influenza vaccinations received. If receipt of the zoster vaccine presented an opportunity for the health system to additionally provide other preventive health interventions to the patient, then we may expect this adjustment to alter our effect sizes. The effect sizes, however, remained very similar (**Supplement Table S4**, Column 5). To provide further reassurance in this regard, we also examined the effect of zoster vaccine receipt on the probability of taking up preventive health measures (receipt of at least one influenza vaccine, statin use, use of an antihypertensive drug, and participation in breast cancer screening) during the follow-up period. None of these analyses showed significant effects (**Supplement Fig. S8**).

#### Robustness checks to different analytical specifications

We conducted a series of additional robustness checks to ensure that our results are not substantially affected by a specific analytical choice that we made in cases for which other possible choices could have been justified. First, instead of starting the follow-up period for all individuals on September 1 2013, we adjusted the follow-up period to account for the staggered rollout of the program by beginning the follow-up period for each individual on the date on which they first became eligible for the zoster vaccine (as detailed in the section “Description of the zoster vaccine rollout in Wales”). We controlled for cohort fixed effects in these analyses to account for the one-to two-year (depending on the year of the program) differences between cohorts in the calendar year in which this moving follow-up window started. That is, we defined one cohort fixed effect for ineligible individuals and the first catch-up cohort and then included additional cohort fixed effects for each group of patients who became eligible at the same time. The effects in this specification were very similar to those in our primary analysis (**Supplement Fig. S13**). Second, we implemented the same analysis as our primary analysis but when restricting the sample to those 237,196 (84.0% of the analysis cohort for our primary analyses) patients who had made at least one visit to their primary care provider during each of the five years preceding the start date of the zoster vaccine program. The rationale for this robustness check is that it estimates the causal effect of the zoster vaccine among the group of patients that is likely to be screened more regularly for dementia given that they interact frequently with the health system. Our effect sizes are similar among this group as in our full sample (**Supplement Table S4**, Column 4). Third, we varied our definition of a new diagnosis of dementia by implementing our analysis separately for each of these two definitions of dementia: i) dementia being named as a primary or contributory cause of death on the death certificate; and ii) a new prescription of donepezil hydrochloride, galantamine, rivastigmine, or memantine hydrochloride. We observed significant protective effects from the zoster vaccine on each of these two different outcome definitions (**Supplement Table S4**, Columns 2 and 3). Fourth, we show all results for our primary analysis with follow-up periods of 5.0., 5.5, 6.0, 6.5, 7.0, 7.5, and 8.0 years, grace periods (i.e., time periods since the index date after which follow-up time is considered to begin) of 0, 2, 4, 6, 8, 10, and 12 months, and bandwidth choices of 0.25, 0.50, 0.75, 1.00, 1.25, 1.50, 1.75, and 2.00 times the MSE-optimal bandwidth. Our results were consistent across these different specifications. Fifth, we verified that our results are similar when using a local second-order polynomial specification instead of local linear regression (**Supplement Fig. S6**).

### Ethics

Approval was granted by the Information Governance Review Panel (IGRP, application number: 1306). Composed of government, regulatory and professional agencies, the IGRP oversees and approves applications to use the SAIL databank.

## Data Availability

The data that support the findings of this study are available from the SAIL Databank. Researchers must request access to the data directly from SAIL. The authors have no permission to share the data. All analysis code will be posted in a publicly accessibly repository upon acceptance of the manuscript for publication in a peer-reviewed journal.

https://saildatabank.com/data/apply-to-work-with-the-data/

## Acknowledgements

This study makes use of anonymized data held in the SAIL Databank. We would like to acknowledge all the data providers who made anonymized data available for research. The responsibility for the interpretation of the data supplied by SAIL is the authors’ alone. SAIL bears no responsibility for the further analysis or interpretation of their data, over and above that published by SAIL.

## Author contributions

M.E. and M.X. contributed equally to this work. M.E. co-conceived the study, devised the methodology, analyzed and processed the data, created data visualizations, interpreted the results, wrote the methods section of the original draft, and reviewed and edited the original draft. M.X. co-conceived the study, devised the methodology, analyzed and processed the data, created data visualizations, interpreted the results, and reviewed and edited the original draft. S.H. devised the methodology, interpreted the results, and reviewed and edited the original draft. P.G. conceived the overall project, acquired funding, co-conceived the study, devised the methodology, was responsible for administration and supervision, interpreted the results, and wrote the original draft.

## Additional Information

Supplementary Information is available for this paper. Correspondence and requests for materials should be addressed to Pascal Geldsetzer.

## References

1. Pickett, J. & Brayne, C. The scale and profile of global dementia research funding. Lancet 394, 1888–1889 (2019).

2. Cummings, J. L., Morstorf, T. & Zhong, K. Alzheimer’s disease drug-development pipeline: few candidates, frequent failures. Alzheimers Res Ther 6, 37 (2014).

3. Cummings, J. et al. Alzheimer’s disease drug development pipeline: 2022. Alzheimers Dement (N Y) 8, e12295 (2022).

4. Gouilly, D. et al. Beyond the amyloid cascade: An update of Alzheimer’s disease pathophysiology. Rev Neurol (Paris) S0035-3787(23)00870–6 (2023) doi:10.1016/j.neurol.2022.12.006.

5. Itzhaki, R. F. et al. Microbes and Alzheimer’s Disease. J Alzheimers Dis 51, 979–984 (2016).

6. Devanand, D. P. Viral Hypothesis and Antiviral Treatment in Alzheimer’s Disease. Curr Neurol Neurosci Rep 18, 55 (2018).

7. Moir, R. D., Lathe, R. & Tanzi, R. E. The antimicrobial protection hypothesis of Alzheimer’s disease. Alzheimers Dement 14, 1602–1614 (2018).

8. Itzhaki, R. F. Overwhelming Evidence for a Major Role for Herpes Simplex Virus Type 1 (HSV1) in Alzheimer’s Disease (AD); Underwhelming Evidence against. Vaccines (Basel) 9, 679 (2021).

9. Eimer, W. A. et al. Alzheimer’s Disease-Associated β-Amyloid Is Rapidly Seeded by Herpesviridae to Protect against Brain Infection. Neuron 99, 56–63.e3 (2018).

10. Devanand, D. P. et al. Antiviral therapy: Valacyclovir Treatment of Alzheimer’s Disease (VALAD) Trial: protocol for a randomised, double-blind,placebo-controlled, treatment trial. BMJ Open 10, e032112 (2020).

11. Wu, X. et al. Adult Vaccination as a Protective Factor for Dementia: A Meta-Analysis and Systematic Review of Population-Based Observational Studies. Front Immunol 13, 872542 (2022).

12. Hernan, M. A. & Robins, J. M. Causal Inference: What if. (Chapman & Hall/CRC, 2020).

13. Schnier, C., Janbek, J., Lathe, R. & Haas, J. Reduced dementia incidence after varicella zoster vaccination in Wales 2013-2020. Alzheimers Dement (N Y) 8, e12293 (2022).

14. Douros, A., Ante, Z., Suissa, S. & Brassard, P. Common vaccines and the risk of incident dementia: a population-based cohort study. J Infect Dis jiac484 (2022) doi:10.1093/infdis/jiac484.

15. Verreault, R., Laurin, D., Lindsay, J. & De Serres, G. Past exposure to vaccines and subsequent risk of Alzheimer’s disease. CMAJ 165, 1495–1498 (2001).

16. Wiemken, T. L. et al. Comparison of rates of dementia among older adult recipients of two, one, or no vaccinations. J Am Geriatr Soc 70, 1157–1168 (2022).

17. Tyas, S. L., Manfreda, J., Strain, L. A. & Montgomery, P. R. Risk factors for Alzheimer’s disease: a population-based, longitudinal study in Manitoba, Canada. Int J Epidemiol 30, 590–597 (2001).

18. Kim, J. I. et al. Intravesical Bacillus Calmette-Guérin Treatment Is Inversely Associated With the Risk of Developing Alzheimer Disease or Other Dementia Among Patients With Non-muscle-invasive Bladder Cancer. Clin Genitourin Cancer 19, e409–e416 (2021).

19. Gofrit, O. N. et al. Bacillus Calmette-Guérin (BCG) therapy lowers the incidence of Alzheimer’s disease in bladder cancer patients. PLoS One 14, e0224433 (2019).

20. Ukraintseva, S., et al. P4-436: Vaccination Against Pneumonia in the Elderly May Reduce Alzheimer’s Risk. Alzheimer’s & Dementia 15, P1469–P1470 (2019).

21. Imbens, G. W. & Lemieux, T. Regression discontinuity designs: A guide to practice. Journal of Econometrics 142, 615–635 (2008).

22. Moscoe, E., Bor, J. & Bärnighausen, T. Regression discontinuity designs are underutilized in medicine, epidemiology, and public health: a review of current and best practice. J Clin Epidemiol 68, 122–33 (2015).

23. Bor, J., Moscoe, E., Mutevedzi, P., Newell, M. L. & Barnighausen, T. Regression discontinuity designs in epidemiology: causal inference without randomized trials. Epidemiology 25, 729–37 (2014).

24. Venkataramani, A. S., Bor, J. & Jena, A. B. Regression discontinuity designs in healthcare research. BMJ 352, i1216 (2016).

25. UK Health Security Agency. Shingles: guidance and vaccination programme. Shingles: guidance and vaccination programme https://www.gov.uk/government/collections/shingles-vaccination-programme (2021).

26. Pan, C. X., Lee, M. S. & Nambudiri, V. E. Global herpes zoster incidence, burden of disease, and vaccine availability: a narrative review. Ther Adv Vaccines Immunother 10, 25151355221084536 (2022).

27. Swansea University. SAIL Databank. https://saildatabank.com/.

28. Ford, D. V. et al. The SAIL Databank: building a national architecture for e-health research and evaluation. BMC Health Serv Res 9, 157 (2009).

29. NHS Digital. Attribution Data Set GP-Registered Populations Scaled to ONS Population Estimates -2011. https://digital.nhs.uk/data-and-information/publications/statistical/attribution-dataset-gp-registered-populations/attribution-data-set-gp-registered-populations-scaled-to-ons-population-estimates-2011.

30. Oxman, M. N. et al. A vaccine to prevent herpes zoster and postherpetic neuralgia in older adults. N Engl J Med 352, 2271–2284 (2005).

31. Alzheimer’s Association. 2022 Alzheimer’s disease facts and figures. Alzheimers Dement 18, 700–789 (2022).

32. Boyle, P. A. et al. Attributable risk of Alzheimer’s dementia attributed to age-related neuropathologies. Ann Neurol 85, 114–124 (2019).

33. Boyle, P. A. et al. Person-specific contribution of neuropathologies to cognitive loss in old age. Ann Neurol 83, 74–83 (2018).

34. Salomon, J. A. Disability-Adjusted Life Years. in Encyclopedia of Health Economics (ed. Culyer, A. J.) 200–203 (Elsevier, 2014). doi:10.1016/B978-0-12-375678-7.00511-3.

35. Global Burden of Disease Project 2019. GBD Compare. http://vizhub.healthdata.org/gbd-compare (2020).

36. Fleming, D. M., Cross, K. W., Cobb, W. A. & Chapman, R. S. Gender difference in the incidence of shingles. Epidemiol Infect 132, 1–5 (2004).

37. Marra, F., Parhar, K., Huang, B. & Vadlamudi, N. Risk Factors for Herpes Zoster Infection: A Meta-Analysis. Open Forum Infect Dis 7, ofaa005 (2020).

38. Ferretti, M. T. et al. Sex and gender differences in Alzheimer’s disease: current challenges and implications for clinical practice: Position paper of the Dementia and Cognitive Disorders Panel of the European Academy of Neurology. Eur J Neurol 27, 928–943 (2020).

39. Ferretti, M. T. et al. Sex differences in Alzheimer disease - the gateway to precision medicine. Nat Rev Neurol 14, 457–469 (2018).

40. Laws, K. R., Irvine, K. & Gale, T. M. Sex differences in Alzheimer’s disease. Curr Opin Psychiatry 31, 133–139 (2018).

41. National Institute for Health and Care Excellence. Dementia: assessment, management and support for people living with dementia and their carers. https://www.nice.org.uk/guidance/ng97 (2018).

42. Centers for Disease Control and Prevention. Archived CDC Vaccine Price List as of June 1, 2018. Adult Vaccine Price List https://www.cdc.gov/vaccines/programs/vfc/awardees/vaccine-management/price-list/2018/2018-06-01.html (2022).

43. Singhal, P. K. & Zhang, D. Costs of adult vaccination in medical settings and pharmacies: an observational study. J Manag Care Spec Pharm 20, 930–936 (2014).

44. Prosser, L. A. et al. A Cost-Effectiveness Analysis of Vaccination for Prevention of Herpes Zoster and Related Complications: Input for National Recommendations. Ann Intern Med 170, 380–388 (2019).

45. Ross, E. L., Weinberg, M. S. & Arnold, S. E. Cost-effectiveness of Aducanumab and Donanemab for Early Alzheimer Disease in the US. JAMA Neurol 79, 478–487 (2022).

46. Lin, G. et al. Beta-Amyloid Antibodies for Early Alzheimer’s Disease: Effectiveness and Value; Draft Evidence Report. https://icer.org/assessment/alzheimers-disease-2022/#timeline (2022).

47. Huo, Z. et al. Cost-effectiveness of pharmacological therapies for people with Alzheimer’s disease and other dementias: a systematic review and meta-analysis. Cost Eff Resour Alloc 20, 19 (2022).

48. Morrison, V. A. et al. Long-term persistence of zoster vaccine efficacy. Clin Infect Dis 60, 900–909 (2015).

49. Baxter, R. et al. Long-Term Effectiveness of the Live Zoster Vaccine in Preventing Shingles: A Cohort Study. Am J Epidemiol 187, 161–169 (2018).

50. Strezova, A. et al. Long-term Protection Against Herpes Zoster by the Adjuvanted Recombinant Zoster Vaccine: Interim Efficacy, Immunogenicity, and Safety Results up to 10 Years After Initial Vaccination. Open Forum Infect Dis 9, ofac485 (2022).

51. McGuinness, L. A., Warren-Gash, C., Moorhouse, L. R. & Thomas, S. L. The validity of dementia diagnoses in routinely collected electronic health records in the United Kingdom: A systematic review. Pharmacoepidemiol Drug Saf 28, 244–255 (2019).

52. Wilkinson, T. et al. Identifying dementia cases with routinely collected health data: A systematic review. Alzheimers Dement 14, 1038–1051 (2018).

53. Wilkinson, T. et al. Identifying dementia outcomes in UK Biobank: a validation study of primary care, hospital admissions and mortality data. Eur J Epidemiol 34, 557–565 (2019).

54. Anderson, M., et al. United Kingdom: Health System Review. Health Syst Transit 24, 1–194 (2022).

55. Digital Health and Care Wales. Welsh Demographic Service Dataset (WDSD). https://web.www.healthdatagateway.org/dataset/cea328df-abe5-48fb-8bcb-c0a5b6377446# (2022).

56. Statistical Directorate. Welsh Index of Multiple Deprivation 2011: Summary Report. https://www.gov.wales/welsh-index-multiple-deprivation-full-index-update-ranks-2011 (2011).

57. SAIL Databank. Welsh Longitudinal General Practice Dataset (WLGP) - Welsh Primary Care. https://web.www.healthdatagateway.org/dataset/33fc3ffd-aa4c-4a16-a32f-0c900aaea3d2 (2023).

58. Benson, T. The history of the Read Codes: the inaugural James Read Memorial Lecture 2011. Inform Prim Care 19, 173–182 (2011).

59. Digital Health and Care Wales. Patient Episode Database for Wales. Digital Health and Care Wales https://dhcw.nhs.wales/information-services/health-intelligence/pedw-data-online/ (2020).

60. NHS England. OPCS Classification of Interventions and Procedures. https://www.datadictionary.nhs.uk/supporting_information/opcs_classification_of_intervention s_and_procedures.html (2023).

61. World Health Organization. ICD-10 : international statistical classification of diseases and related health problems : tenth revision. https://apps.who.int/iris/handle/10665/42980 (2004).

62. Digital Health and Care Wales. Outpatient Database for Wales (OPDW). https://web.www.healthdatagateway.org/dataset/d331159b-b286-4ab9-8b36-db39123ec229 (2022).

63. Public Health Wales. Welsh Cancer Intelligence and Surveillance Unit (WCISU). Public Health Wales https://phw.nhs.wales/services-and-teams/welsh-cancer-intelligence-and-surveillance-unit-wcisu/ (2023).

64. Digital Health and Care Wales. Annual District Death Extract (ADDE). https://web.www.healthdatagateway.org/dataset/15cf4241-abad-4dcc-95b0-8cd7c02be999 (2023).

65. Villamizar-Villegas, M., Pinzon-Puerto, F. A. & Ruiz-Sanchez, M. A. A comprehensive history of regression discontinuity designs: An empirical survey of the last 60 years. Journal of Economic Surveys **Epub ahead of print**, (2022).

66. Gelman, A. & Imbens, G. Why High-Order Polynomials Should Not Be Used in Regression Discontinuity Designs. Journal of Business & Economic Statistics 1–10 (2017) doi:10.1080/07350015.2017.1366909.

67. Imbens, G. & Kalyanaraman, K. Optimal Bandwidth Choice for the Regression Discontinuity Estimator. The Review of Economic Studies 79, 933–959 (2012).

68. Calonico, S., Cattaneo, M. D. & Titiunik, R. Robust Nonparametric Confidence Intervals for Regression-Discontinuity Designs. Econometrica 82, 2295–2326 (2014).

69. McCrary, J. Manipulation of the running variable in the regression discontinuity design: A density test. Journal of Econometrics 142, 698–714 (2008).

