## Supplement for "Causal evidence that herpes zoster vaccination prevents a proportion of dementia cases"

This PDF file includes:
- Materials
- Figs. S1 to S16
- Tables S1 to S4

#### Materials

Below we list Read version 2 (used for all care processes in primary care electronic health record data) and ICD-10 (used for diagnoses in secondary care records and death certificates) codes for all key variables in our analysis.

Herpes zoster vaccination

| Read code | | | | | | ICD-10 code |
|---|---|---|---|---|---|---|
| 65FY. | 65FY.1 | 65FY.11 | 65FY0 | n4v1. | n4v4. | Not applicable |

***Primary and secondary outcomes***

Dementia of any type

| Read code | | | | | | | | | | ICD-10 code | | | | | | | | | |
|---|---|---|---|---|---|---|---|---|---|---|---|---|---|---|---|---|---|---|---|
| 1461 | 38C13 | 3AE3. | 3AE4. | 3AE5. | 3AE6. | 66h.. | 6AB.. | 8BM02 | 8CMG2 | F00 | F00, | F00. | F00` | F000 | F001 | F002 | F003 | F004 | F007 |
| 8CMZ. | 8CMZ0 | 8CMZ1 | 8CMZ2 | 8CMZ3 | 8CSA. | 9hD.. | 9hD0. | 9hD1. | 9Ou.. | F008 | F009 | F00A | F00X | F01 | F01. | F010 | F011 | F012 | F013 |
| 9Ou1. | 9Ou2. | 9Ou3. | 9Ou4. | 9Ou5. | A411. | A4110 | E00.. | E00..1 | E00..11 | F018 | F019 | F01X | F02 | F02. | F020 | F021 | F022 | F023 | F025 |
| E00..2 | E00..97 | E000. | E001. | E0010 | E0011 | E0012 | E0013 | E001z | E002. | F028 | F029 | F02X | F03 | F03. | F030 | F031 | F032 | F033 | F034 |
| E0020 | E0021 | E002z | E003. | E004. | E004.1 | E004.11 | E0040 | E0041 | E0042 | F038 | F039 | F03X | F051 | G30 | G30. | G300 | G301 | G308 | G309 |
| E0043 | E004z | E012. | E012.1 | E0120 | E02y1 | E041. | Eu00. | Eu000 | Eu00011 | G30D | G30X | G310 | G311 | G318 | I673 | | | | |
| Eu001 | Eu00112 | Eu0012 | Eu002 | Eu00z | Eu00z1 | Eu00z11 | Eu01. | Eu010 | Eu011 | | | | | | | | | | |
| Eu012 | Eu013 | Eu01y | Eu01z | Eu02. | Eu020 | Eu021 | Eu022 | Eu023 | Eu025 | | | | | | | | | | |
| Eu02y | Eu02z | Eu02z13 | Eu02z14 | Eu02z3 | Eu02z4 | Eu041 | Eu106 | Eu1061 | Eu107 | | | | | | | | | | |
| Eu10711 | F110. | F1100 | F1101 | F111. | F112. | F116. | F118. | F1181 | F11x2 | | | | | | | | | | |
| F11x7 | F11x9 | F11y2 | F21y2 | Fyu30 | | | | | | | | | | | | | | | |

Vascular dementia

| Read code | | | | | | | | | | ICD-10 code | | | | | | | | | |
|---|---|---|---|---|---|---|---|---|---|---|---|---|---|---|---|---|---|---|---|
| E004. | E004.1 | E004.11 | E0040 | E0041 | E0042 | E0043 | E004z | Eu01. | Eu010 | F01 | F01. | F010 | F011 | F012 | F013 | F018 | F019 | F01X | I673 |
| Eu011 | Eu01y | Eu01z | F11x2 | F21y2 | | | | | | | | | | | | | | | |

Alzheimer’s disease

| Read code | | | | | | | | | | ICD-10 code | | | | | | | | | |
|---|---|---|---|---|---|---|---|---|---|---|---|---|---|---|---|---|---|---|---|
| Eu00. | Eu000 | Eu00011 | Eu001 | Eu00112 | Eu0012 | Eu002 | Eu00z | Eu00z1 | Eu00z11 | F00 | F00, | F00. | F00` | F000 | F001 | F002 | F003 | F004 | F007 |
| F110. | F1100 | F1101 | F112. | Fyu30 | | | | | | F008 | F009 | F00A | F00X | G30 | G30. | G300 | G301 | G308 | G309 |
| | | | | | | | | | | G30D | G30X | | | | | | | | |

Dementia of unspecified type

| Read code | | | | | | | | | | ICD-10 code | | | | | | | | | |
|---|---|---|---|---|---|---|---|---|---|---|---|---|---|---|---|---|---|---|---|
| 1461 | 38C13 | 3AE3. | 3AE4. | 3AE5. | 3AE6. | 66h.. | 6AB.. | 8BM02 | 8CMG2 | F02 | F02. | F020 | F021 | F022 | F023 | F025 | F028 | F029 | F02X |
| 8CMZ. | 8CMZ0 | 8CMZ1 | 8CMZ2 | 8CMZ3 | 8CSA. | 9hD.. | 9hD0. | 9hD1. | 9Ou.. | F03 | F03. | F030 | F031 | F032 | F033 | F034 | F038 | F039 | F03X |
| 9Ou1. | 9Ou2. | 9Ou3. | 9Ou4. | 9Ou5. | A411. | A4110 | E00.. | E00..1 | E00..11 | F051 | G310 | G311 | G318 | | | | | | |
| E00..2 | E00..97 | E000. | E001. | E0010 | E0011 | E0012 | E0013 | E001z | E002. | | | | | | | | | | |
| E0020 | E0021 | E002z | E003. | E012. | E012.1 | E0120 | E02y1 | E041. | Eu012 | | | | | | | | | | |
| Eu013 | Eu02. | Eu020 | Eu021 | Eu022 | Eu023 | Eu025 | Eu02y | Eu02z | Eu02z13 | | | | | | | | | | |
| Eu02z14 | Eu02z3 | Eu02z4 | Eu041 | Eu106 | Eu1061 | Eu107 | Eu10711 | F111. | F116. | | | | | | | | | | |
| F118. | F1181 | F11x7 | F11x9 | F11y2 | | | | | | | | | | | | | | | |

New prescription of donepezil hydrochloride, galantamine, rivastigmine, or memantine hydrochloride

| Read code | | | | | | | | | | ICD-10 code |
|---|---|---|---|---|---|---|---|---|---|---|
| dB1.. | dB11. | dB12. | dB13. | dB14. | dB15. | dB16. | dB17. | dB18. | dB19. | Not applicable |
| dy1.. | dy11. | dy12. | dy13. | dy14. | dy15. | dy16. | dy1y. | dy1z. | dy21. | |
| dy22. | dy23. | dy24. | dy25. | dy26. | dy27. | dy28. | dy29. | dy2A. | dy2B. | |
| dy2C. | dy2D. | dy2E. | dy2F. | dy2J. | dy2K. | dy2L. | dy2M. | dy2N. | dy2O. | |
| dy2R. | dy2S. | dy31. | dy32. | dy33. | dy34. | dy35. | dy36. | dy37. | dy38. | |
| dy39. | dy3B. | dy3C. | dy3D. | dy3K. | dy3L. | dy3M. | dy3t. | dy3u. | dy3v. | |
| dy3w. | dy3x. | dy3y. | dy3z. | | | | | | | |

#### Shingles

| Read code | ICD-10 code |
|---|---|
| A53.. A53..1 A53..11 A530. A531. A531.1 A531.11 A5310 A5311 A53111 | B020 B021 B022 B023 B027 B028 B029 B02X |
| A5312 A5313 A5314 A5315 A53151 A531511 A531z A532. A5320 A5321 | |
| A5322 A5323 A5324 A532z A53x. A53x0 A53x1 A53xz A53y. A53z. | |
| AyuA4 AyuA5 F0112 F0309 F300. F3744 F5016 HNG0041 HNG0060 | |

#### Postherpetic neuralgia

| Read code | ICD-10 code |
|---|---|
| A531. A5312 A5313 A5315 A53151 A531511 F300. | B022 |

### ***Outcomes for negative control outcome analyses***

#### Ischemic heart disease

| Read code | ICD-10 code |
|---|---|
| G3... G3...11 G3...13 G3...3 G30.. G30..12 G30..14 G30..15 G30..4 G30..5 | I21 I21. I210 I211 I212 I213 I214 I218 I219 I21X |
| G30..7 G300. G301. G3010 G3011 G301z G302. G303. G304. G305. | I22 I22. I220 I221 I228 I229 I22X I230 I231 I232 |
| G306. G307. G3070 G3071 G308. G309. G30A. G30B. G30X. G30X0 | I233 I234 I235 I236 I238 I240 I241 I248 I249 I24X |
| G30y. G30y0 G30y2 G30yz G30z. G31.. G310. G311. G311.11 G311.13 | |
| G311.14 G311.3 G3110 G3111 G3112 G3113 G3114 G3115 G311z G312. | |
| G31y. G31y0 G31y099 G31y1 G31y2 G31y3 G31yz G32.. G32..11 G35.. | |
| G350. G351. G353. G35X. G36.. G360. G361. G362. G363. G364. | |
| G365. G366. G37.. G38.. G380. G381. G384. G38z. G39.. G3y.. | |
| G3z.. | |

#### Chronic obstructive pulmonary disease

| Read code | ICD-10 code |
|---|---|
| H3... H31.. H310. H3100 H310z H311. H3110 H3111 H311z H312. | J410 J411 J418 J41X J42 J42X J43 J43. J430 J431 |
| H3120 H3121 H3123 H312z H313. H31y. H31y1 H31yz H31z. H32.. | J432 J438 J439 J43X J44 J44. J440 J441 J444 J445 |
| H320. H3200 H3201 H3202 H3203 H320z H321. H322. H32y. H32y0 | J448 J449 J44X |
| H32y1 H32y2 H32yz H32z. H36.. H37.. H38.. H3y.. H3z.. | |

Stroke

| Read code | | | | | | | | | | ICD-10 code | | | | | | | | | |
|---|---|---|---|---|---|---|---|---|---|---|---|---|---|---|---|---|---|---|---|
| F423. | F4231 | F4238 | F423811 | G60.. | G600. | G601. | G602. | G603. | G604. | H341 | I600 | I601 | I602 | I603 | I604 | I605 | I606 | I607 | I608 |
| G605. | G606. | G60X. | G60z. | G61.. | G61..1 | G61..11 | G61..12 | G61..2-5( | G610. | I609 | I60X | I610 | I611 | I612 | I613 | I614 | I615 | I616 | I618 |
| G611. | G612. | G613. | G614. | G615. | G616. | G617. | G618. | G619. | G61X. | I619 | I61X | I63 | I630 | I631 | I632 | I633 | I634 | I635 | I636 |
| G61X0 | G61X1 | G61z. | G63.. | G630. | G631. | G631.1 | G632. | G633. | G634. | I638 | I639 | I63X | I64 | I64. | I640 | I644 | I649 | I64X | |
| G63y. | G63y0 | G63y1 | G63z. | G64.. | G64..1 | G64..11 | G64..2 | G640. | G6400 | | | | | | | | | | |
| G641. | G6410 | G64z. | G64z.12 | G64z.2 | G64z0 | G64z1 | G64z2 | G64z3 | G64z4 | | | | | | | | | | |
| G66.. | G66..1 | G66..11 | G66..12 | G66..13 | G66..2 | G66..3 | | G660. | G661. | | | | | | | | | | |
| G662. | G663. | G664. | G665. | G666. | G667. | G668. | G669. | | | | | | | | | | | | |

#### Lower respiratory tract infection

| Read code | | | | | | | | | |
|---|---|---|---|---|---|---|---|---|---|
| 16L.. | 1J72. | 1J72.1 | 1J72.11 | 1JX.. | 1W0.. | A33.. | A330. | A331. | A33y. |
| A33yz | A33z. | A521. | A54x4 | A730. | A7850 | A795. | A7951 | A7952 | A7953 |
| A7954 | A7955 | A7y00 | AB24. | AB405 | Ayu39 | Ayu3A | Ayu3U | AyuDC | H0... |
| H06.. | H060. | H060.1 | H060.11 | H0600 | H0601 | H0602 | H0603 | H0604 | H0605 |
| H0606 | H0607 | H0608 | H0609 | H060A | H060B | H060C | H060D | H060E | H060F |
| H060v | H060w | H060x | H060z | H061. | H0610 | H0611 | H0612 | H0613 | H0614 |
| H0615 | H0616 | H0617 | H061z | H062. | H06z. | H06z0 | H06z01 | H06z011 | H06z1 |
| H06z11 | H06z111 | H06z112 | H06z12 | H06z2 | H07.. | H2... | H2...SHH | H20.. | H200. |
| H201. | H202. | H203. | H204. | H20y. | H20y0 | H20z. | H21.. | H22.. | H22..1 |
| H220. | H221. | H222. | H223. | H2230 | H224. | H22y. | H22y0 | H22y1 | H22y2 |
| H22y3 | H22yX | H22yz | H22z. | H23.. | H230. | H231. | H232. | H233. | H23z. |
| H24.. | H240. | H241. | H242. | H243. | H246. | H247. | H2470 | H247z | H24y. |
| H24y0 | H24y2 | H24y3 | H24y4 | H24y7 | H24yz | H24z. | H25.. | H25..1 | H25..11 |
| H25..99 | H26.. | H26..1 | H26..11 | H260. | H2600 | H261. | H262. | H263. | H27.. |
| H270. | H2700 | H2701 | H270z | H271. | H2710 | H2711 | H271z | H27y. | H27y0 |
| H27y1 | H27yz | H27z. | H27z.1 | H27z.11 | H27z.12 | H27z.2 | H28.. | H29.. | H2A.. |
| H2A..1 | H2A..11 | H2B.. | H2C.. | H2F.. | H2Fy. | H2Fz. | H2y.. | H2z.. | H2z..98 |
| H30.. | H30..1 | H30..11 | H30..12 | H30..2 | H300. | H301. | H302. | H30z. | H31.. |
| H310. | H3100 | H3101 | H310z | H311. | H3110 | H3111 | H311z | H312. | H3120 |
| H312011 | H3121 | H3122 | H312250( | H3123 | H312z | H313. | H31y. | H31y0 | H31y1 |
| H31yz | H31z. | H3y.. | H3y0. | H3y1. | H3z.. | H3z..11 | H47.. | H47..1 | H47..11 |
| H47..99 | H470. | H470.1 | H470.11 | H4700 | H4701 | H4702 | H4703 | H470312 | H470z |
| H50.. | H500. | H5000 | H5001 | H5003 | H5004 | H5005 | H500z | H501. | H5010 |
| H5011 | H5012 | H5013 | H5014 | H5015 | H5016 | H501z | H50z. | H511. | H5110 |
| H5111 | H5112 | H511z | H53.. | H530. | H5300 | H5301 | H5302 | H5303 | H530z |
| H5400 | H5401 | H571. | Hyu1. | Hyu10 | Hyu11 | Q310. | Q3100 | Q3101 | Q3102 |
| Q3103 | Q3104 | Q3105 | Q3106 | Q310y | Q310z | SP131 | SP132 | | |

| ICD-10 code | | | | | | | | | |
|---|---|---|---|---|---|---|---|---|---|
| A370 | A371 | A378 | A379 | A37X | B012 | B052 | B250 | J041 | J042 |
| J100 | J101 | J108 | J10X | J11 | J110 | J111 | J118 | J11X | J120 |
| J121 | J122 | J123 | J128 | J129 | J13 | J13X | J14 | J14X | J150 |
| J151 | J152 | J153 | J154 | J155 | J156 | J157 | J158 | J159 | J160 |
| J168 | J17 | J170 | J171 | J172 | J173 | J178 | J17X | J18 | J180 |
| J181 | J182 | J188 | J189 | J18O | J18X | J200 | J201 | J202 | J203 |
| J204 | J205 | J206 | J207 | J208 | J209 | J20X | J210 | J211 | J218 |
| J219 | J21X | J22 | J22# | J220 | J229 | J22X | J22Y | J440 | J851 |
| J860 | J869 | P230 | P231 | P232 | P233 | P234 | P235 | P236 | P238 |
| P239 | U049 | | | | | | | | |

#### Lung cancer

| Read code |
|---|
| Not applicable |

| ICD-10 code | | | | | | | | | |
|---|---|---|---|---|---|---|---|---|---|
| C33 | C33X | C34 | C34, | C340 | C341 | C342 | C343 | C344 | C348 |
| C349 | C34X | | | | | | | | |

Pancreatic cancer

| Read code | ICD-10 code |
|---|---|
| Not applicable | C25 C250 C251 C252 C253 C254 C257 C258 C259 |

Colorectal cancer

| Read code | ICD-10 code |
|---|---|
| Not applicable | C18 C18. C180 C181 C182 C183 C184 C185 C186 C187 C188 C189 C18X C19 C19X C20 C20X C210 C211 C212 C218 |

Breast cancer

| Read code | ICD-10 code |
|---|---|
| Not applicable | C50 C500 C501 C502 C503 C504 C505 C506 C508 C509 C50X |

<u>Falls</u>

| Read code | | | | | | | | | |
|---|---|---|---|---|---|---|---|---|---|
| TC... | TC...1 | TC...11 | TC0.. | TC00. | TC000 | TC001 | TC00z | TC01. | TC010 |
| TC011 | TC01z | TC02. | TC020 | TC021 | TC02z | TC0z. | TC1.. | TC10. | TC11. |
| TC1z. | TC2.. | TC20. | TC21. | TC22. | TC23. | TC24. | TC27. | TC28. | TC29. |
| TC2z. | TC3.. | TC30. | TC300 | TC302 | TC305 | TC30z | TC31. | TC32. | TC320 |
| TC321 | TC32z | TC3y. | TC3y0 | TC3y1 | TC3y2 | TC3y3 | TC3y4 | TC3y6 | TC3yz |
| TC3z. | TC4.. | TC40. | TC41. | TC42. | TC420 | TC421 | TC42z | TC4y. | TC4y0 |
| TC4y1 | TC4y2 | TC4y3 | TC4yz | TC4z. | TC5.. | TC50. | TC51. | TC52. | TC53. |
| TC5z. | TC6.. | TC60. | TC600 | TC60y | TC60z | TC6y. | TC6y0 | TC6y1 | TC6yz |
| TC6z. | TC7.. | TCLO16[illegible] | TCy.. | TCy0. | TCyz. | TCz.. | | | |

| ICD-10 code | | | | | | | | | |
|---|---|---|---|---|---|---|---|---|---|
| W000 | W001 | W002 | W003 | W004 | W005 | W006 | W007 | W008 | W009 |
| W00X | W01 | W01. | W010 | W011 | W012 | W013 | W014 | W015 | W016 |
| W017 | W018 | W019 | W01X | W020 | W021 | W022 | W023 | W024 | W025 |
| W026 | W027 | W028 | W029 | W02X | W03 | W03. | W030 | W031 | W032 |
| W033 | W034 | W035 | W036 | W037 | W038 | W039 | W03X | W04. | W040 |
| W041 | W042 | W043 | W044 | W045 | W046 | W048 | W049 | W04X | W05 |
| W050 | W051 | W052 | W053 | W054 | W055 | W056 | W058 | W059 | W05X |
| W06 | W060 | W061 | W062 | W063 | W064 | W065 | W068 | W069 | W06X |
| W07 | W070 | W071 | W072 | W073 | W074 | W075 | W076 | W077 | W078 |
| W079 | W07X | W080 | W081 | W082 | W083 | W084 | W085 | W086 | W087 |
| W088 | W089 | W08X | W090 | W091 | W092 | W093 | W094 | W095 | W097 |
| W098 | W099 | W09X | W10 | W100 | W101 | W102 | W103 | W104 | W105 |
| W106 | W107 | W108 | W109 | W10X | W11 | W110 | W111 | W112 | W113 |
| W114 | W115 | W116 | W117 | W118 | W119 | W11X | W120 | W121 | W122 |
| W123 | W124 | W125 | W126 | W128 | W129 | W13 | W130 | W131 | W132 |
| W133 | W134 | W135 | W136 | W137 | W138 | W139 | W13X | W140 | W141 |
| W142 | W143 | W144 | W145 | W147 | W148 | W149 | W14X | W150 | W152 |
| W153 | W154 | W155 | W156 | W158 | W159 | W160 | W161 | W162 | W163 |
| W164 | W165 | W166 | W168 | W169 | W170 | W171 | W172 | W173 | W174 |
| W175 | W176 | W177 | W178 | W179 | W17X | W18 | W180 | W181 | W182 |
| W183 | W184 | W185 | W186 | W187 | W188 | W189 | W18X | W19 | W19. |
| W190 | W191 | W192 | W193 | W194 | W195 | W196 | W197 | W198 | W199 |
| W19O | W19X | | | | | | | | |

<u>Lower back pain</u>

| Read code | | | | | | | | | |
|---|---|---|---|---|---|---|---|---|---|
| 16C2. | 16C3. | 16C4. | 16C5. | 16C6. | 16C8. | 16C9. | 16CA. | 16CZ. | N14.. |
| N140. | N140.1 | N140.11 | N1400 | N1401 | N1402 | N1403 | N1404 | N1407 | N1408 |
| N140A | N140z | N141. | N141.1 | N141.11 | N142. | N142.1 | N142.11 | N142.12 | N142.13 |
| N142.14 | N142.3 | N142.4 | N1420 | N143. | N143.1 | N143.11 | N144. | N1440 | N1441 |
| N144z | N145. | N145.1 | N145.11 | N145.12 | N145.2 | N146. | N1460 | N1461 | N1463 |
| N14631 | N146311 | N1464 | N1465 | N1466 | N146z | N146z1 | N146z11 | N147. | N1470 |
| N1471 | N1472 | N14721 | N147211 | N147z | N148. | N1480 | N1481 | N1482 | N1483 |
| N1484 | N1485 | N1486 | N1487 | N1488 | N1489 | N148A | N148B | N148C | N149. |
| N14A. | N14X. | N14y. | N14y.1 | N14z. | N14z.2 | | | | |

| ICD-10 code | | | | | | | | | |
|---|---|---|---|---|---|---|---|---|---|
| G544 | M470 | M471 | M472 | M478 | M480 | M481 | M482 | M483 | M484 |
| M485 | M498 | M510 | M511 | M512 | M513 | M514 | M518 | M533 | M538 |
| M540 | M541 | M543 | M544 | M990 | M991 | M992 | M993 | M994 | M995 |
| M996 | M997 | M998 | | | | | | | |

Diabetes:
Please see separate pdf file (Supplement Materials 2).

#### *Preventive interventions other than zoster vaccination*[1]

[1] ICD-10 codes are not applicable for these interventions.

Influenza vaccination

| Read code | | | | | | | | | |
|---|---|---|---|---|---|---|---|---|---|
| 65E.. | 65E0. | 65E00 | 65E1. | 65E10 | 65E2. | 65E20 | 65E21 | 65E22 | 65E23 |
| 65E24 | 65E3. | 65E30 | 65E4. | 65E40 | 65E5. | 65E6. | 65E7. | 65E8. | 65E9. |
| 65EA. | 65EB. | 65EC. | 65ED. | 65ED0 | 65ED1 | 65ED2 | 65ED3 | 65ED4 | 65ED5 |
| 65ED6 | 65ED7 | 65ED8 | 65ED9 | 65EDA | 65EE. | 65EE0 | 65EE1 | n47.. | n471. |
| n472. | n473. | n476. | n477. | n47a. | n47A. | n47b. | n47B. | n47c. | n47C. |
| n47d. | n47D. | n47e. | n47E. | n47f. | n47F. | n47g. | n47G. | n47h. | n47H. |
| n47i. | n47I. | n47j. | n47k. | n47l. | n47m. | n47n. | n47o. | n47p. | n47q. |
| n47r. | n47u. | n47v. | n47z. | | | | | | |

Pneumococcal vaccination (PPV-23)

| Read code | | | | | | | | | |
|---|---|---|---|---|---|---|---|---|---|
| 6572. | 65720 | 657K. | 657L. | 657M. | 657N. | 657P. | 657R. | n4b.. | n4b1. |
| n4b2. | n4b4. | n4b9. | | | | | | | |

Statin use

| Read code | | | | | | | | | |
|---|---|---|---|---|---|---|---|---|---|
| bxd.. | bxd1. | bxd2. | bxd3. | bxd4. | bxd5. | bxd6. | bxd7. | bxd8. | bxd9. |
| bxdA. | bxdB. | bxdC. | bxdD. | bxdE. | bxdF. | bxdH. | bxdI. | bxdJ. | bxdK. |
| bxdu. | bxdv. | bxdw. | bxdx. | bxdy. | bxdz. | bxe.. | bxe1. | bxe2. | bxe3. |
| bxe4. | bxe5. | bxe6. | bxe7. | bxe8. | bxg1. | bxg2. | bxg3. | bxg4. | bxg5. |
| bxgz. | bxi.. | bxi1. | bxi2. | bxi3. | bxi4. | bxi5. | bxi6. | bxi7. | bxi8. |
| bxi9. | bxiA. | bxiB. | bxiy. | bxiz. | bxj1. | bxj2. | bxj3. | bxj4. | bxj5. |
| bxj6. | bxj7. | bxj8. | bxjz. | bxk1. | bxk2. | bxk3. | bxk4. | bxkw. | bxkx. |
| bxky. | bxkz. | | | | | | | | |

Use of antihypertensive medications
Please see separate pdf file (Supplement Materials 2).

Breast cancer screening

| Read code | | | | | | | | | |
|---|---|---|---|---|---|---|---|---|---|
| 1A81. | 1A82. | 1A84. | 5371 | 5372 | 5373 | 5376 | 54L3. | 56B1. | 585C. |
| 5861 | 6862 | 68620 | 68621 | 6862Z | 6865 | 7P0F. | 7P0F0 | 7P0F1 | 7P0F2 |
| 7P0Fy | 7P0Fz | 8HTh. | 8HTl. | 9N0A. | 9N1y9 | 9Np2. | 9OH9. | 9OHZ. | 9Oq8. |
| L001. | R138. | R1380 | R138z | ZV761 | | | | | |

#### Figs. S1 to S16

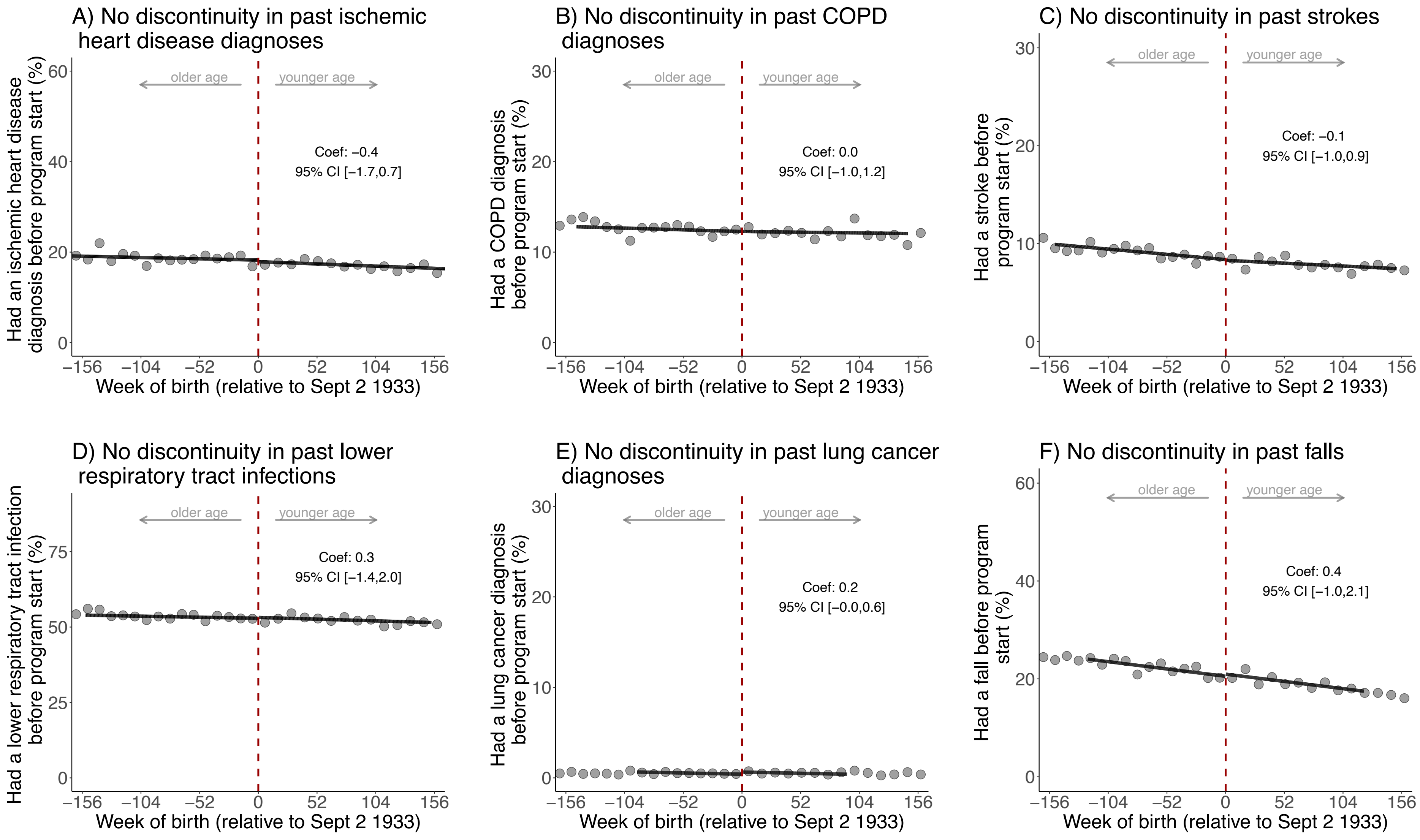

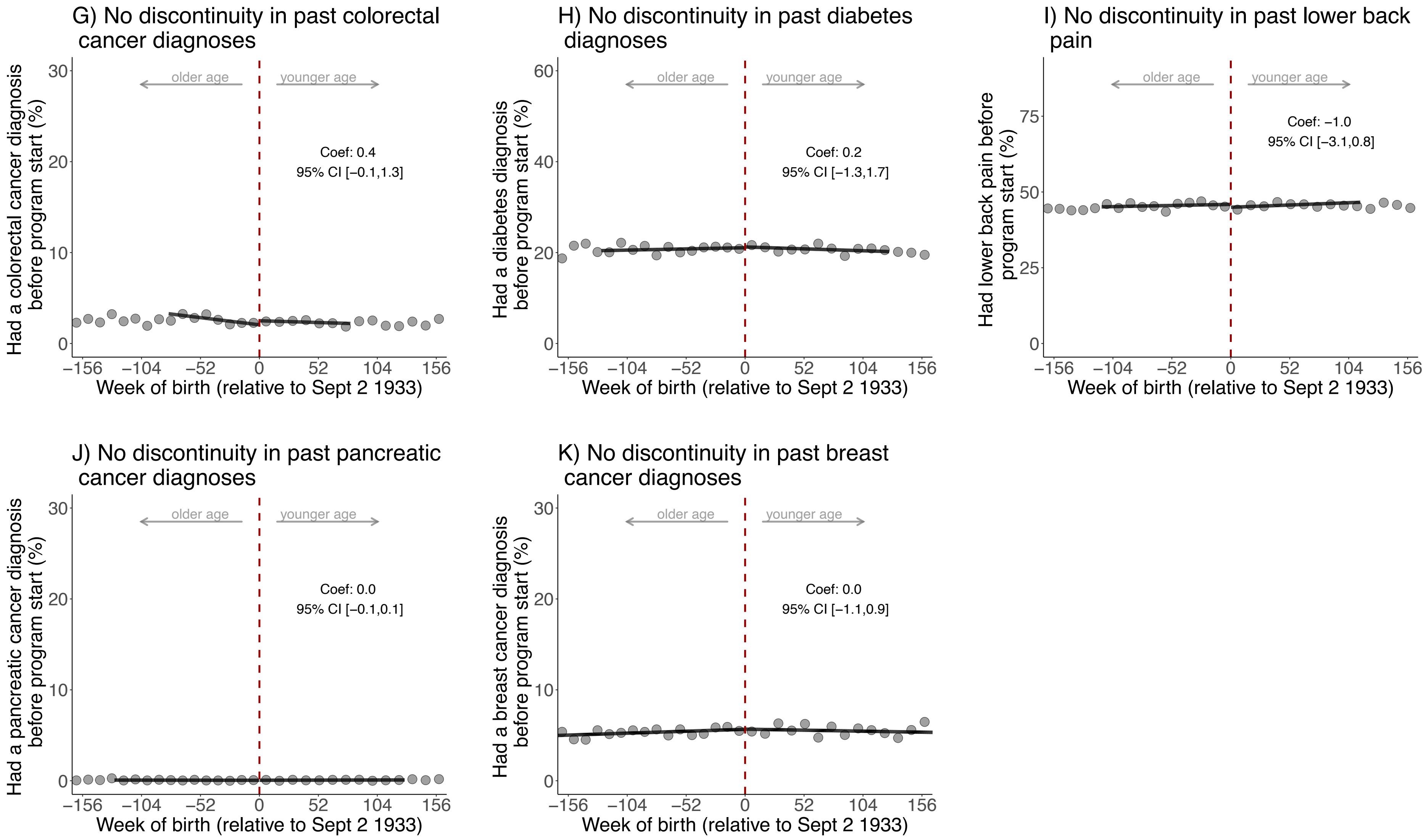

**Fig. S1**: There is exchangeability at baseline across the date-of-birth eligibility cutoff for the ten (other than dementia) leading causes of disability-adjusted life years and mortality in Wales in 2019.[1,2]

[1] Grey dots show the mean value for each 10-week increment in week of birth.

[2] The analysis of breast cancer diagnoses was restricted to women only.

Abbreviations: Coef=coefficient; CI=confidence interval; COPD=chronic obstructive pulmonary disease

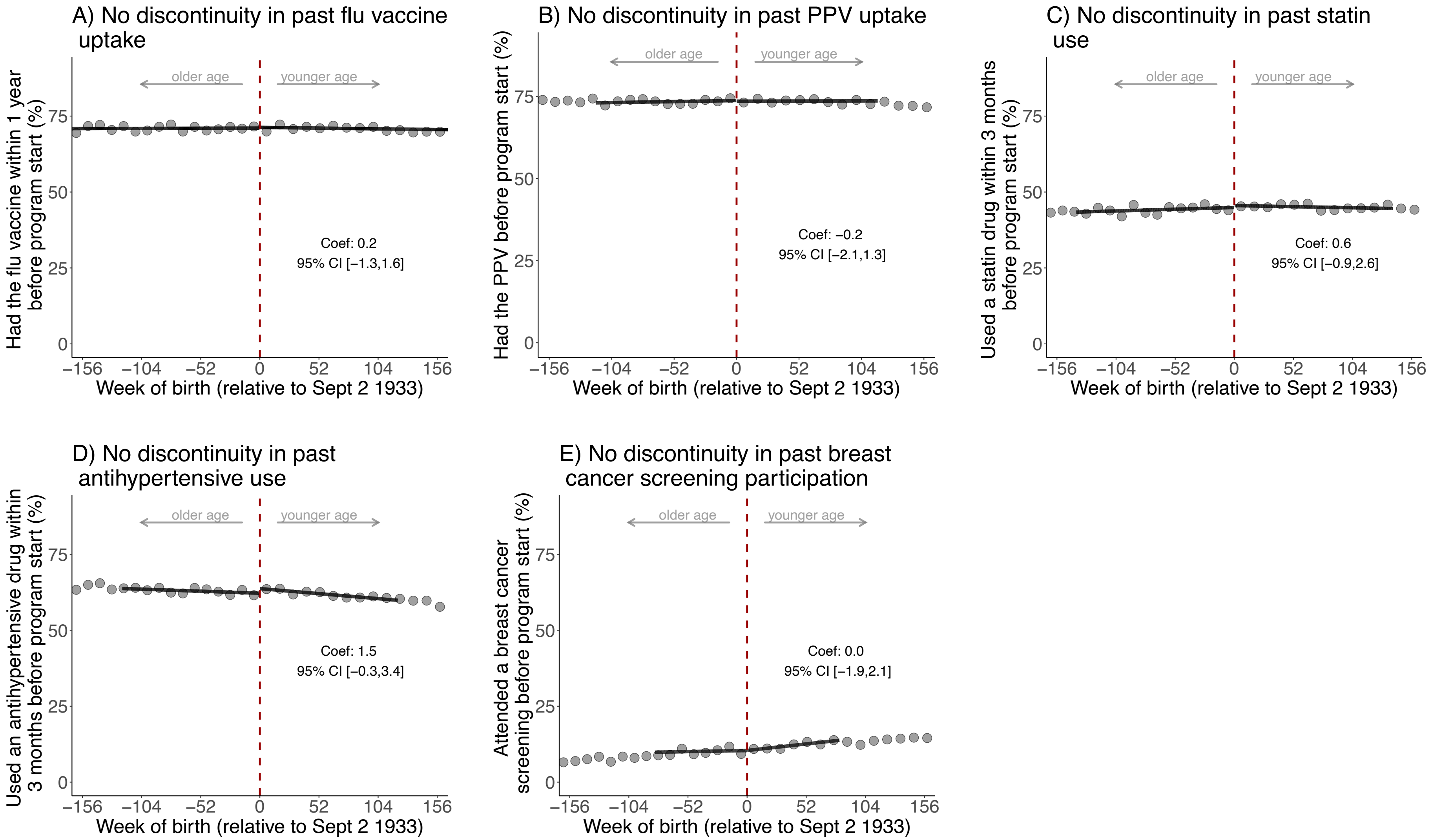

**Fig. S2**: No abrupt change at the date-of-birth eligibility cutoff in the probability of having taken up preventive health measures before the start date of the zoster vaccine program.[1,2,3]

[1] Panels A, B, and C are also shown in Fig. 1 in the main manuscript. They have been repeated here for comprehensiveness as flu vaccine uptake, PPV uptake, and statin use also constitute preventive health measures.

[2] Breast cancer screening participation was defined as having a record of referral to, attendance at, or a report from “breast cancer screening” or mammography. The analysis of breast cancer screening participation was restricted to women only.

[3] Grey dots show the mean value for each 10-week increment in week of birth.

Abbreviations: PPV=pneumococcal polysaccharide vaccine.

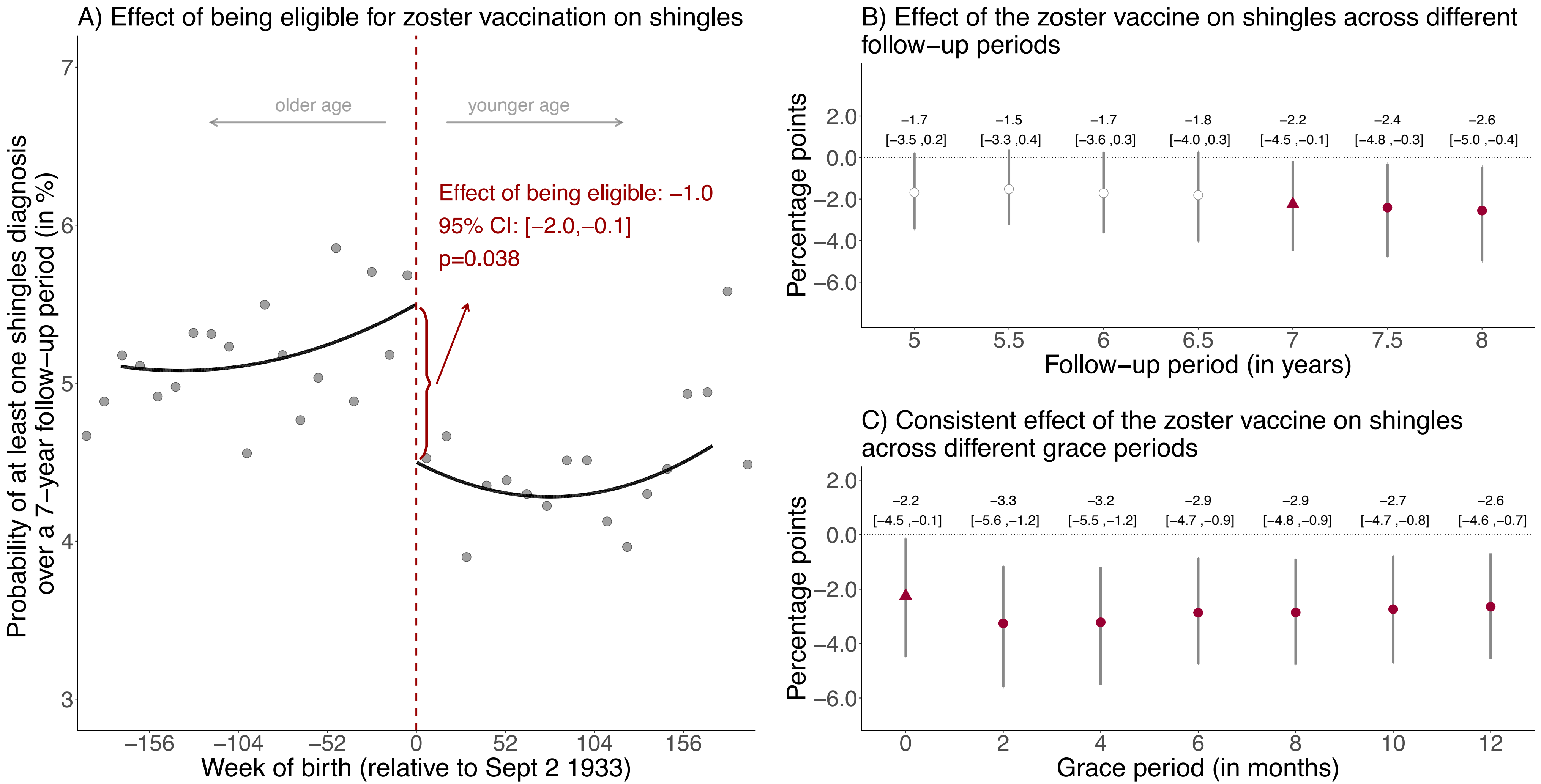

**Fig. S3**: Effect estimates of being eligible (A) and having received the zoster vaccine (B and C) on having at least one shingles diagnosis using local squared regression instead of local linear regression.[1,2,3,4,5]

[1] Triangles (rather than points) depict our primary specification.
[2] Red (as opposed to white) fillings denote statistical significance (p<0.05).
[3] With "grace periods" we refer to time periods since the index date after which follow-up time is considered to begin to allow for the time needed for a full immune response to develop after vaccine administration.
[4] Grey vertical bars depict 95% confidence intervals.
[5] Grey dots show the mean value for each 10-week increment in week of birth.

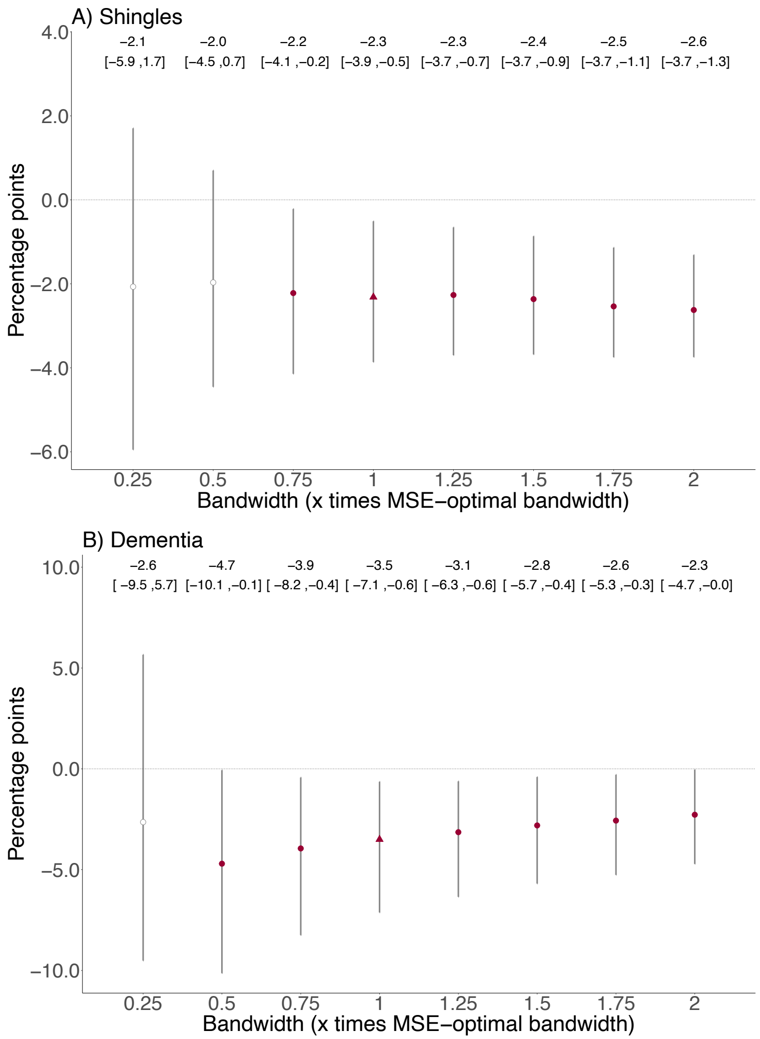

**Fig. S4**: Point estimates for the effect of the zoster vaccine on at least one shingles diagnosis and new dementia diagnoses across different bandwidth specifications.[1,2,3,4,5]

[1] The bandwidth is the window (in weeks) in participants' date of birth that is drawn around the September 2 1933 eligibility threshold. The shorter the bandwidth is, the wider is the 95% confidence interval.

[2] 95% confidence intervals are not symmetrical around the point estimate because we used robust bias-corrected confidence intervals[1].

[3] The MSE-optimal bandwidth is 90.6 and 93.0 weeks on either side of the threshold for the dementia and shingles outcome, respectively.

[4] White points depict statistically insignificant point estimates (p>0.05).

[5] Grey vertical bars depict 95% confidence intervals.

Abbreviations: MSE=mean-squared error

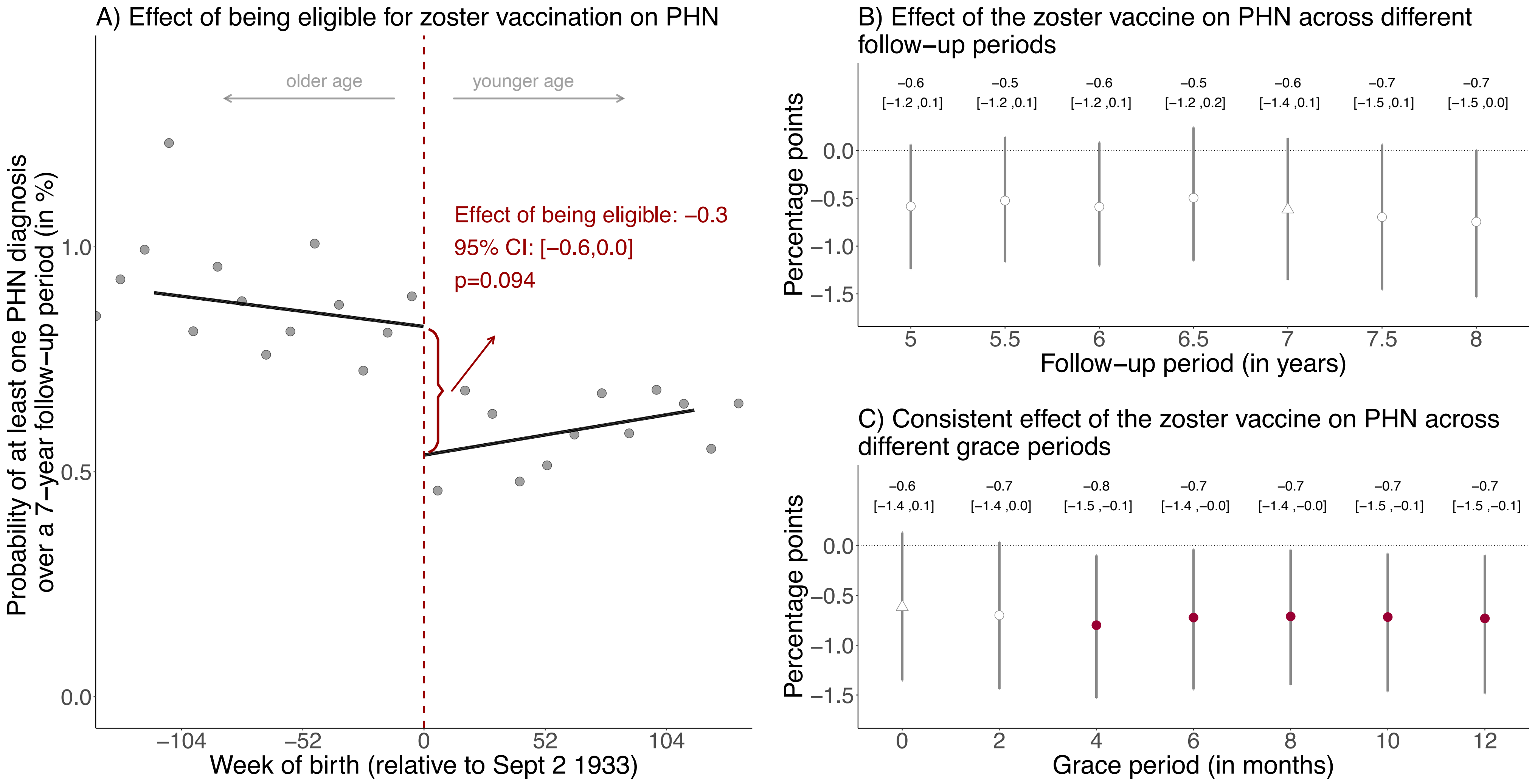

**Fig. S5**: Effect estimates of being eligible (A) and having received the zoster vaccine (B and C) on having at least one diagnosis of postherpetic neuralgia (PHN) during the follow-up period.[1,2,3,4,5]

[1] Triangles (rather than points) depict our primary specification.

[2] Red (as opposed to white) fillings denote statistical significance ($p<0.05$).

[3] With "grace periods" we refer to time periods since the index date after which follow-up time is considered to begin to allow for the time needed for a full immune response to develop after vaccine administration.

[4] Grey vertical bars depict 95% confidence intervals.

[5] Grey dots show the mean value for each 10-week increment in week of birth.

Abbreviations: PHN=postherpetic neuralgia

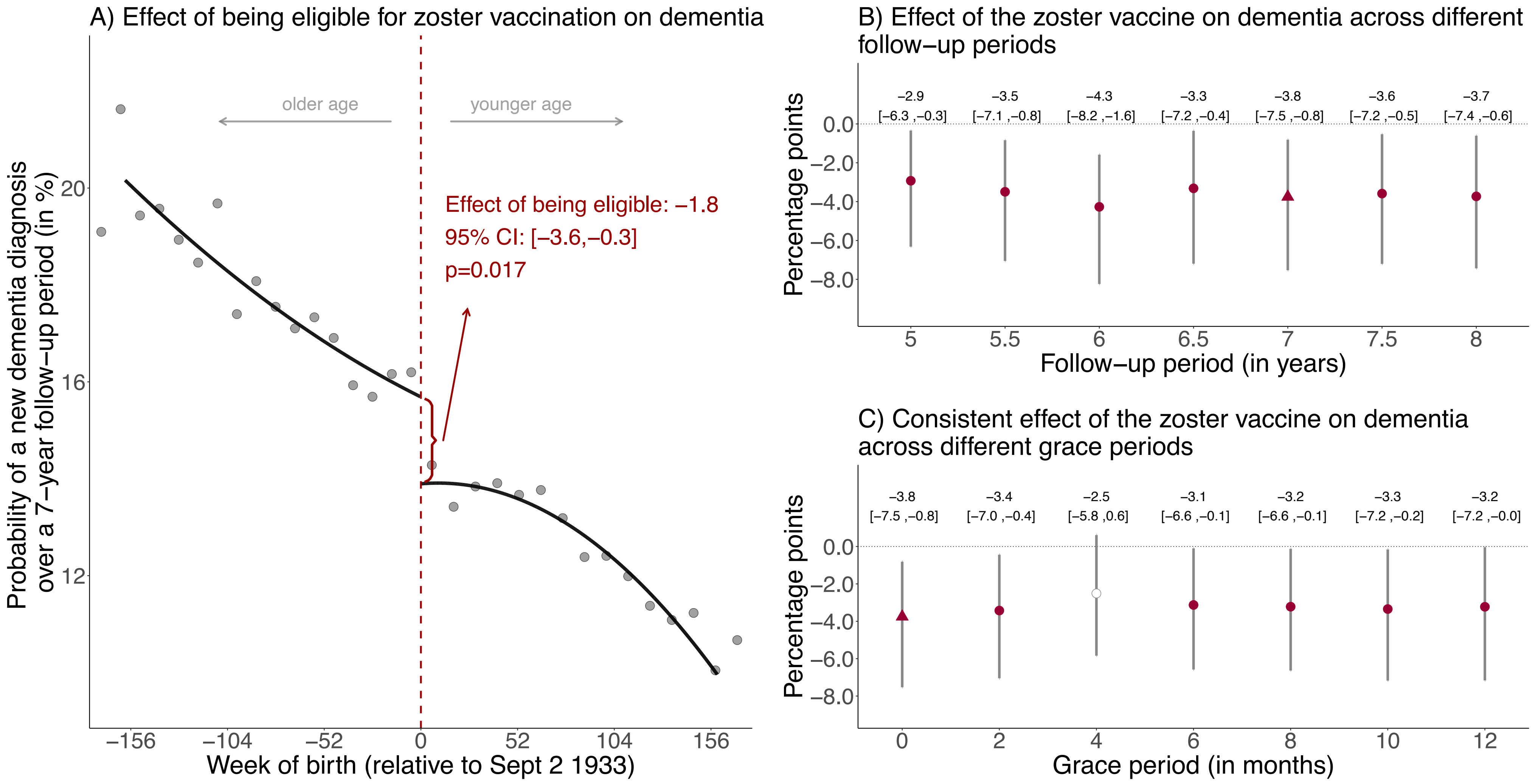

**Fig. S6**: Effect estimates of being eligible (A) and having received the zoster vaccine (B and C) on new diagnoses of dementia using local squared regression instead of local linear regression.[1,2,3,4,5]

[1] Triangles (rather than points) depict our primary specification.
[2] Red (as opposed to white) fillings denote statistical significance (p<0.05).
[3] With "grace periods" we refer to time periods since the index date after which follow-up time is considered to begin to allow for the time needed for a full immune response to develop after vaccine administration.
[4] Grey vertical bars depict 95% confidence intervals.
[5] Grey dots show the mean value for each 10-week increment in week of birth.

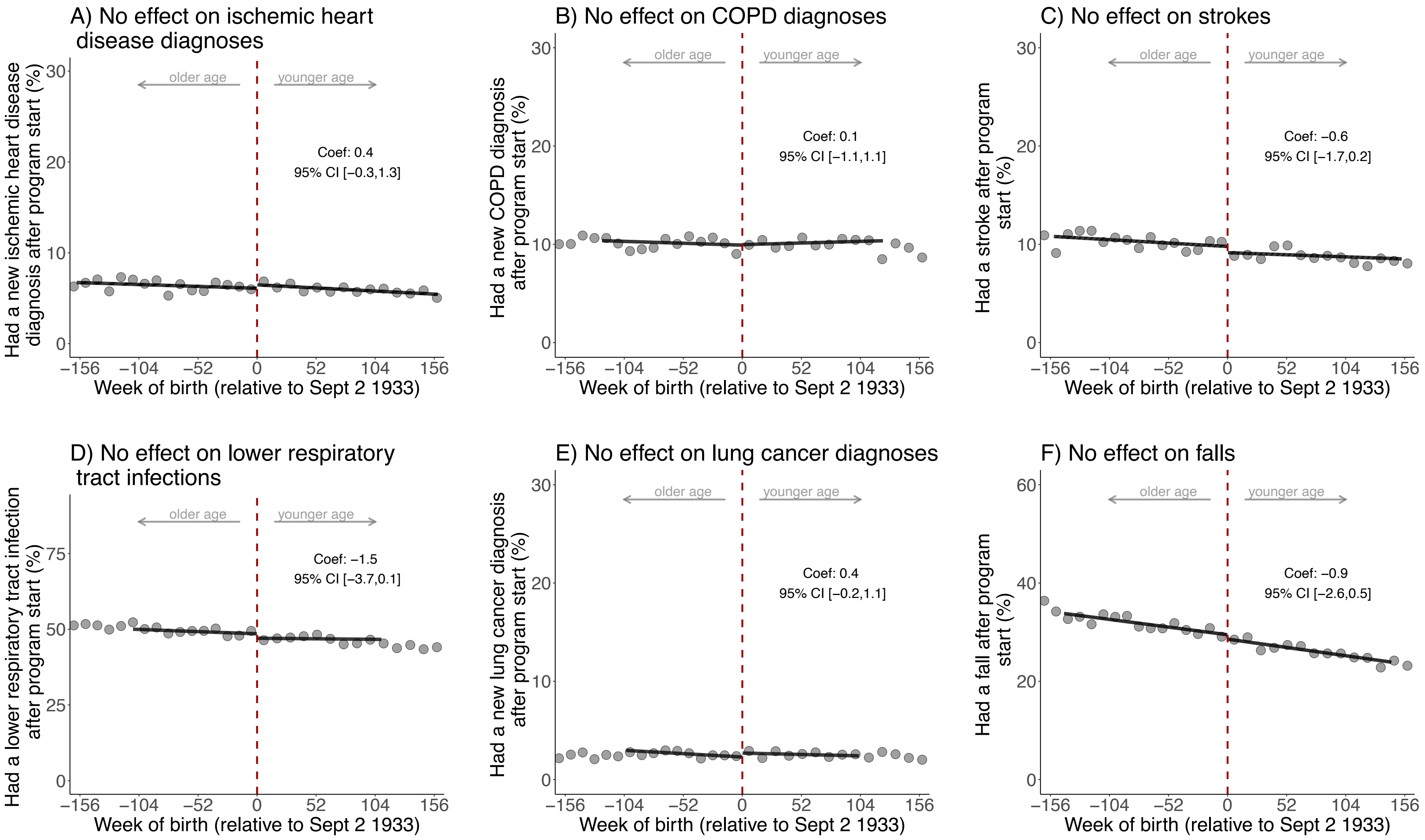
A) No effect on ischemic heart disease diagnoses
Had a new ischemic heart disease diagnosis after program start (%)
older age
younger age
Coef: 0.4
95% CI [−0.3,1.3]
Week of birth (relative to Sept 2 1933)
B) No effect on COPD diagnoses
Had a new COPD diagnosis after program start (%)
Coef: 0.1
95% CI [−1.1,1.1]
C) No effect on strokes
Had a stroke after program start (%)
Coef: −0.6
95% CI [−1.7,0.2]
D) No effect on lower respiratory tract infections
Had a lower respiratory tract infection after program start (%)
Coef: −1.5
95% CI [−3.7,0.1]
E) No effect on lung cancer diagnoses
Had a new lung cancer diagnosis after program start (%)
Coef: 0.4
95% CI [−0.2,1.1]
F) No effect on falls
Had a fall after program start (%)
Coef: −0.9
95% CI [−2.6,0.5]

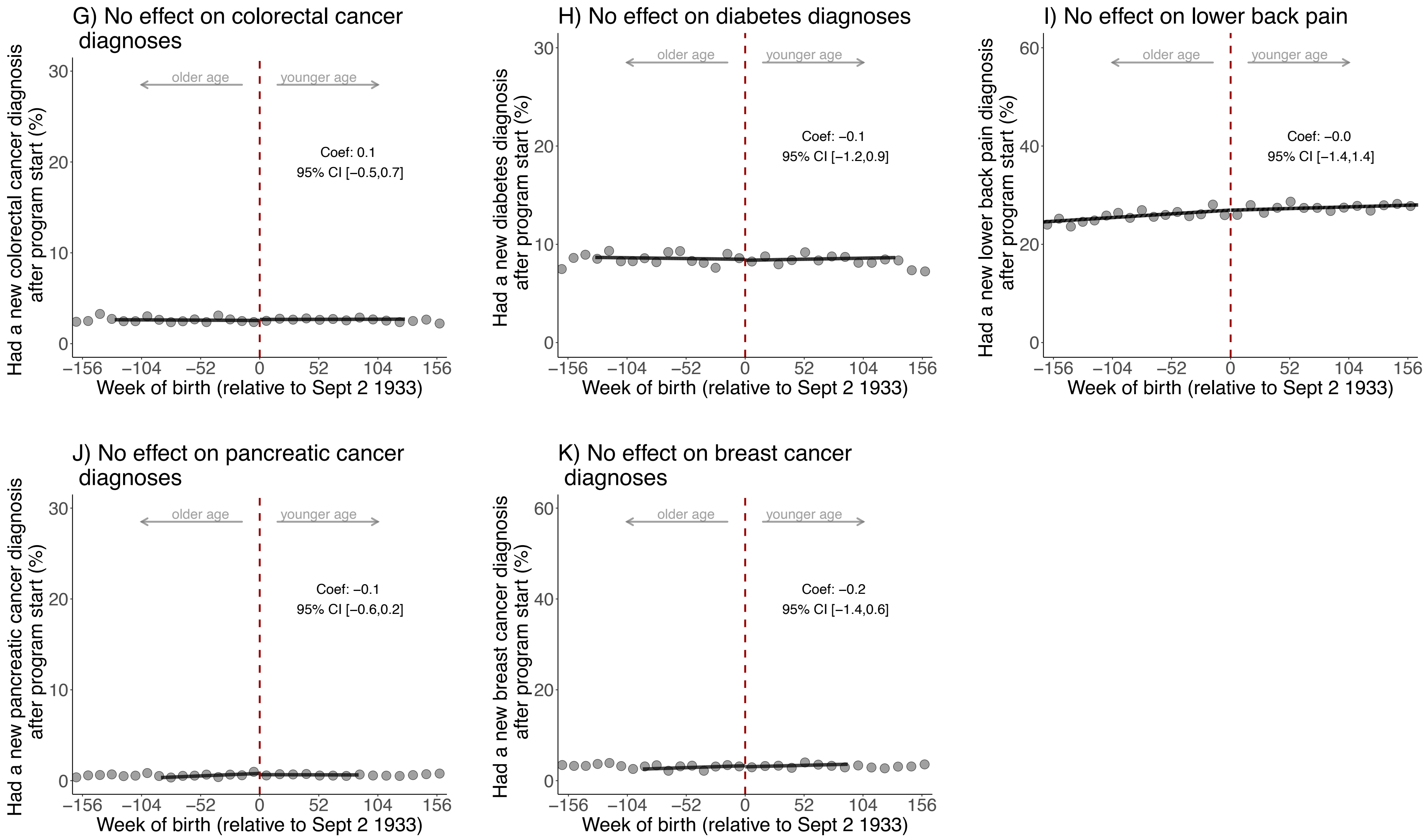

**Fig. S7**: No statistically significant effects of being eligible for the zoster vaccine on each of the ten (other than dementia) leading causes of disability-adjusted life years and mortality in Wales.[1,2,3]

[1] These analyses used the same follow-up period (September 2 1933 to September 1 2020) as our primary analyses for dementia shown in the main manuscript.

[2] The analysis of breast cancer diagnoses was restricted to women only.

[3] Grey dots show the mean value for each 10-week increment in week of birth.

Abbreviations: Coef=coefficient; CI=confidence interval; COPD=chronic obstructive pulmonary disease

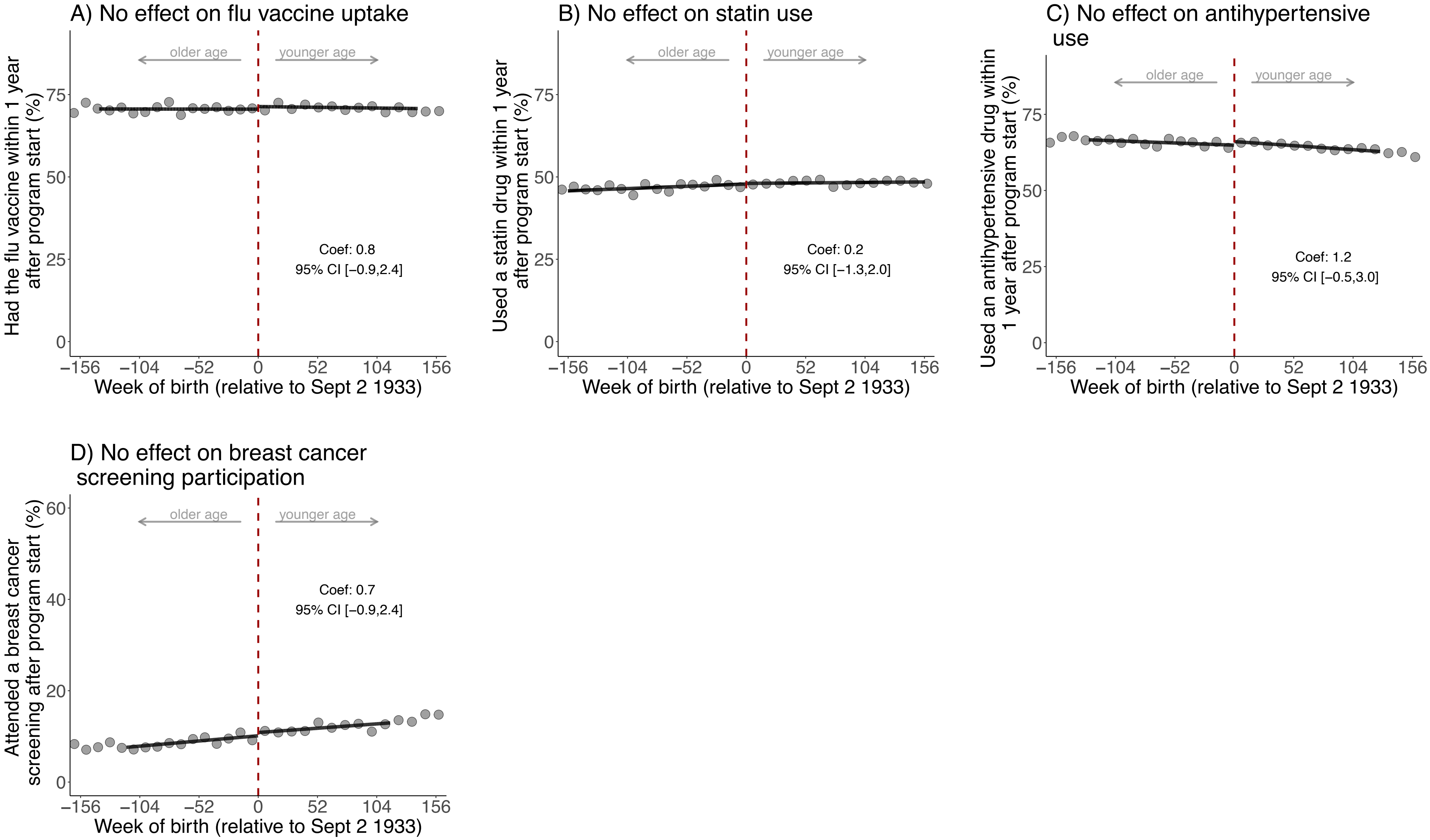

**Fig. S8**: No statistically significant effects of being eligible for the zoster vaccine on preventive health actions.[1,2,3]

[1] These analyses used the same follow-up period (September 2 1933 to September 1 2020) as our primary analyses with dementia as an outcome.

[2] Breast cancer screening participation was defined as having a record of referral to, attendance at, or a report from "breast cancer screening" or mammography. The analysis of breast cancer screening participation was restricted to women only.

[3] Grey dots show the mean value for each 10-week increment in week of birth.

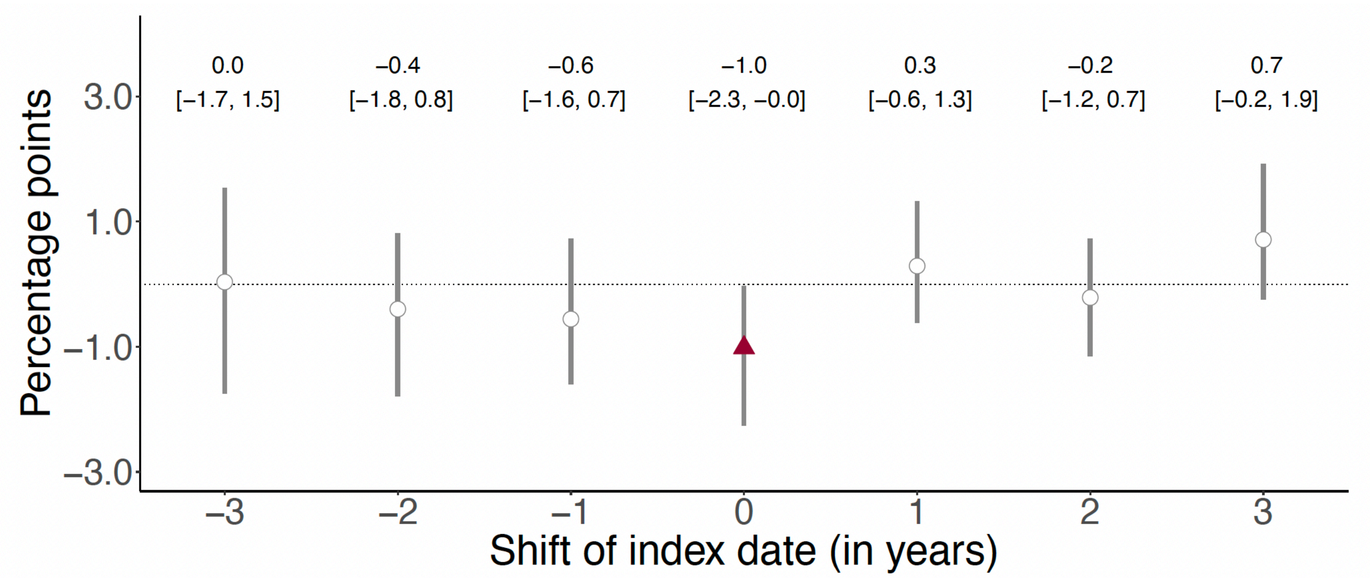

**Fig. S9**: The September 2 date-of-birth threshold only has an effect on the occurrence of dementia over a five-year follow-up period in the year (2013) for which that date was used as eligibility threshold for zoster vaccination.[1,2,3,4]

[1] This analysis implements the identical analysis as for September 1 2013 (the date on which the zoster vaccine program started), for September 1 of each of the three years prior to and after 2013. For example, when moving the start date of the program to "-2" (i.e., September 1 2011), we started the follow-up period on September 1 2011 and compared individuals around the September 2 1931 eligibility threshold. The purpose of this analysis is to verify that the day-month (i.e., September 2) cutoff used for zoster vaccine eligibility was not also used for other interventions that affect dementia risk.

[2] This analysis used a five-year follow-up period to allow each comparison to have the same length of follow-up.

[3] White points depict statistically insignificant point estimates (p>0.05).

[4] Grey vertical bars depict 95% confidence intervals.

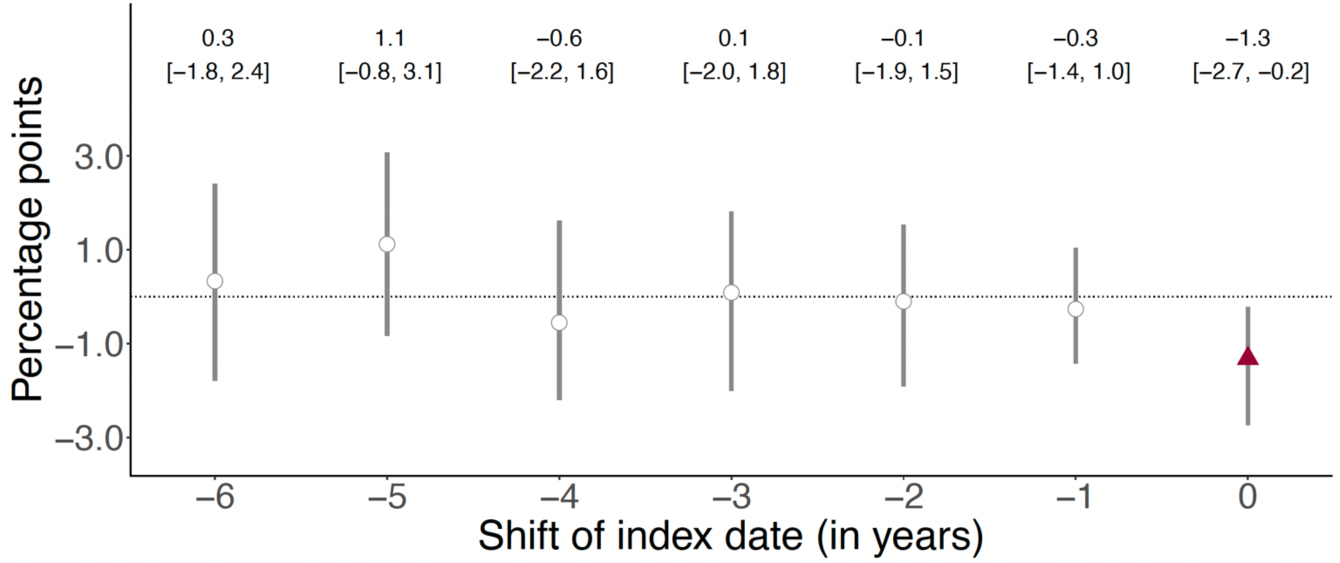

**Fig. S10**: The September 2 date-of-birth threshold only has an effect on the occurrence of dementia over a seven-year follow-up period in the year (2013) for which that date was used as eligibility threshold for zoster vaccination.[1,2,3]

[1] This analysis implements the identical analysis as for September 1 2013 (the date on which the zoster vaccine program started), for September 1 of each of the six years preceding 2013. For example, when moving the start date of the program to "-2" (i.e., September 1 2011), we started the follow-up period on September 1 2011 and compared individuals around the September 2 1931 eligibility threshold. The purpose of this analysis is to verify that the day-month (i.e., September 2) cutoff used for zoster vaccine eligibility was not also used for other interventions that affect dementia risk.

[2] White points depict statistically insignificant point estimates (p>0.05).

[3] Grey vertical bars depict 95% confidence intervals.

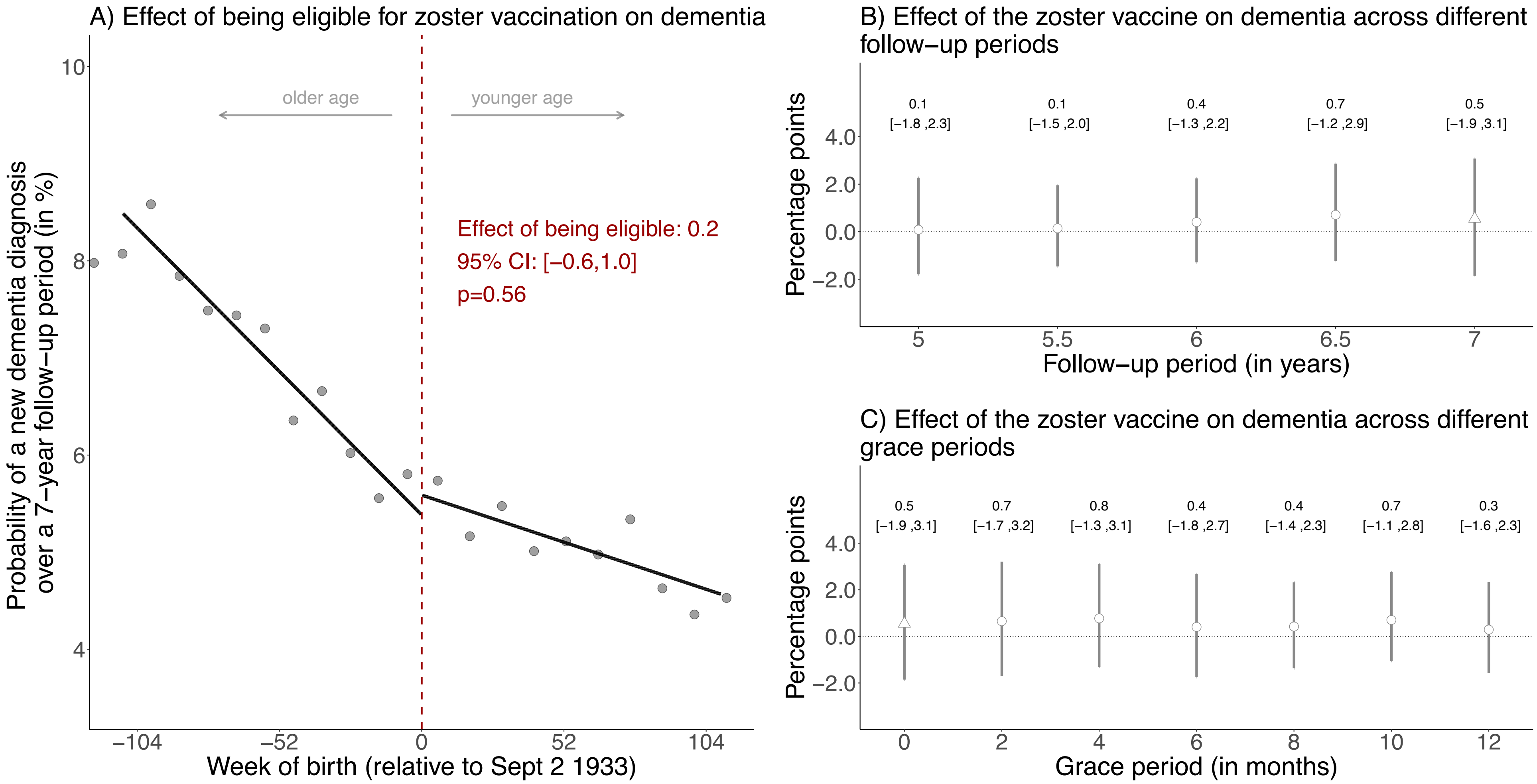

**Fig. S11**: No effects on new diagnoses of dementia diagnosed in the seven years *prior* to the start of the zoster vaccine program (i.e., between September 1 2006 to August 31 2013).[1,2,3,4,5,6]

[1] This figure shows the results from the identical analysis as implemented for our primary analysis (for which the results are shown in Fig. 3 in the main manuscript) except that we followed individuals from September 1 2006 to August 31 2013 instead of from September 1 2013 to August 31 2020. This analysis thus compares the exact same date-of-birth cohorts to each other and has the same length of follow-up as our primary analysis, but uses the seven years prior to the start of the zoster vaccine rollout as follow-up period instead of the seven years after program start. The purpose of this analysis is to verify that the same date-of-birth eligibility date used for zoster vaccine eligibility was not used for other interventions in the past that affect dementia risk.

[2] Triangles (rather than points) depict our primary specification.

[3] Red (as opposed to white) fillings denote statistical significance ($p<0.05$).

[4] With “grace periods” we refer to time periods since the index date after which follow-up time is considered to begin to allow for the time needed for a full immune response to develop after vaccine administration.

[5] Grey vertical bars depict 95% confidence intervals.

[6] Grey dots in Panel A show the mean value for each 10-week increment in week of birth.

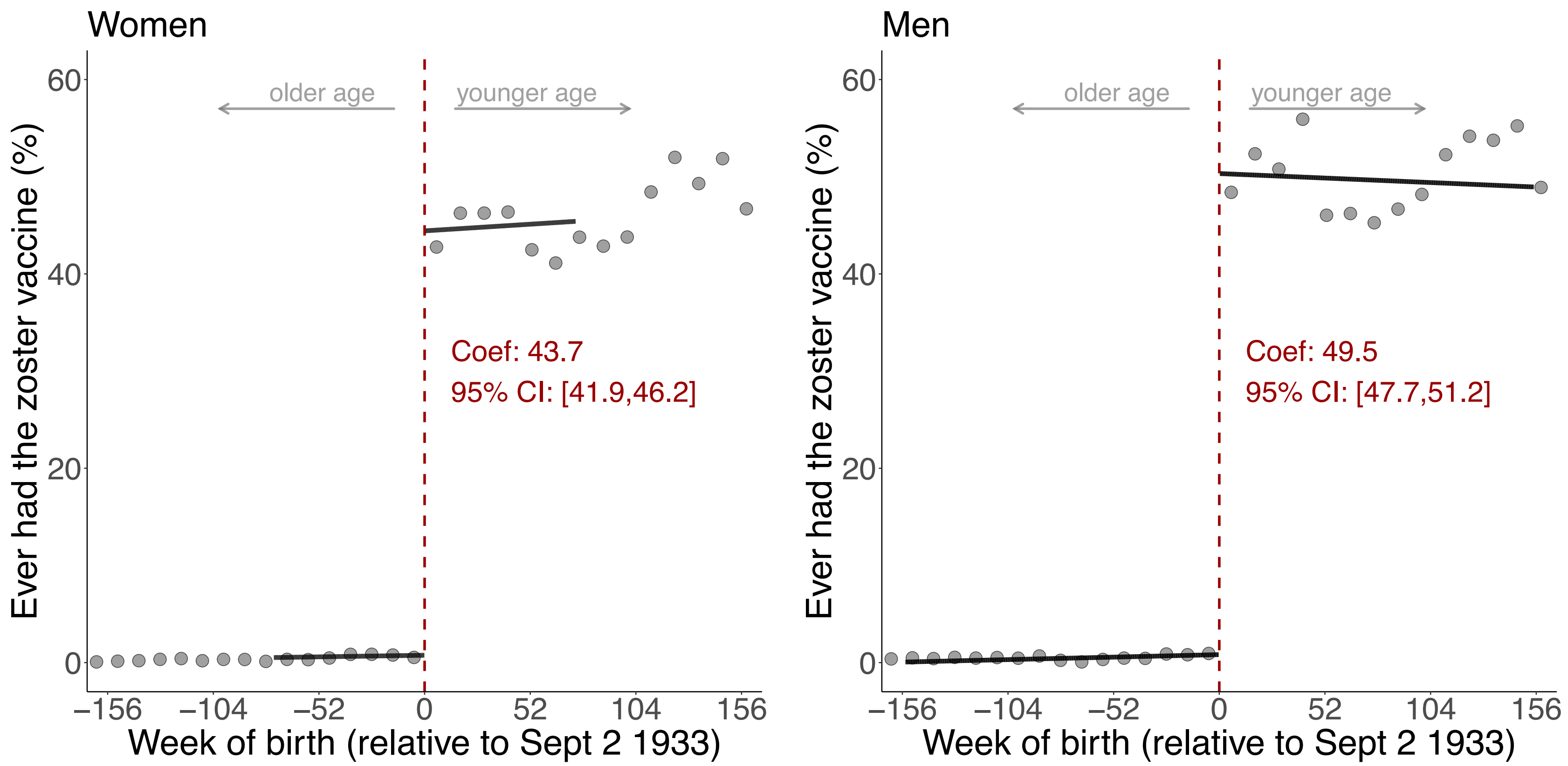

**Fig. S12**: The magnitude of the abrupt change in vaccine uptake at the September 2 1933 date-of-birth eligibility threshold was similar between men and women.[1]

[1] Grey dots show the mean value for each 10-week increment in week of birth.

Abbreviations: Coef=coefficient; CI=confidence interval

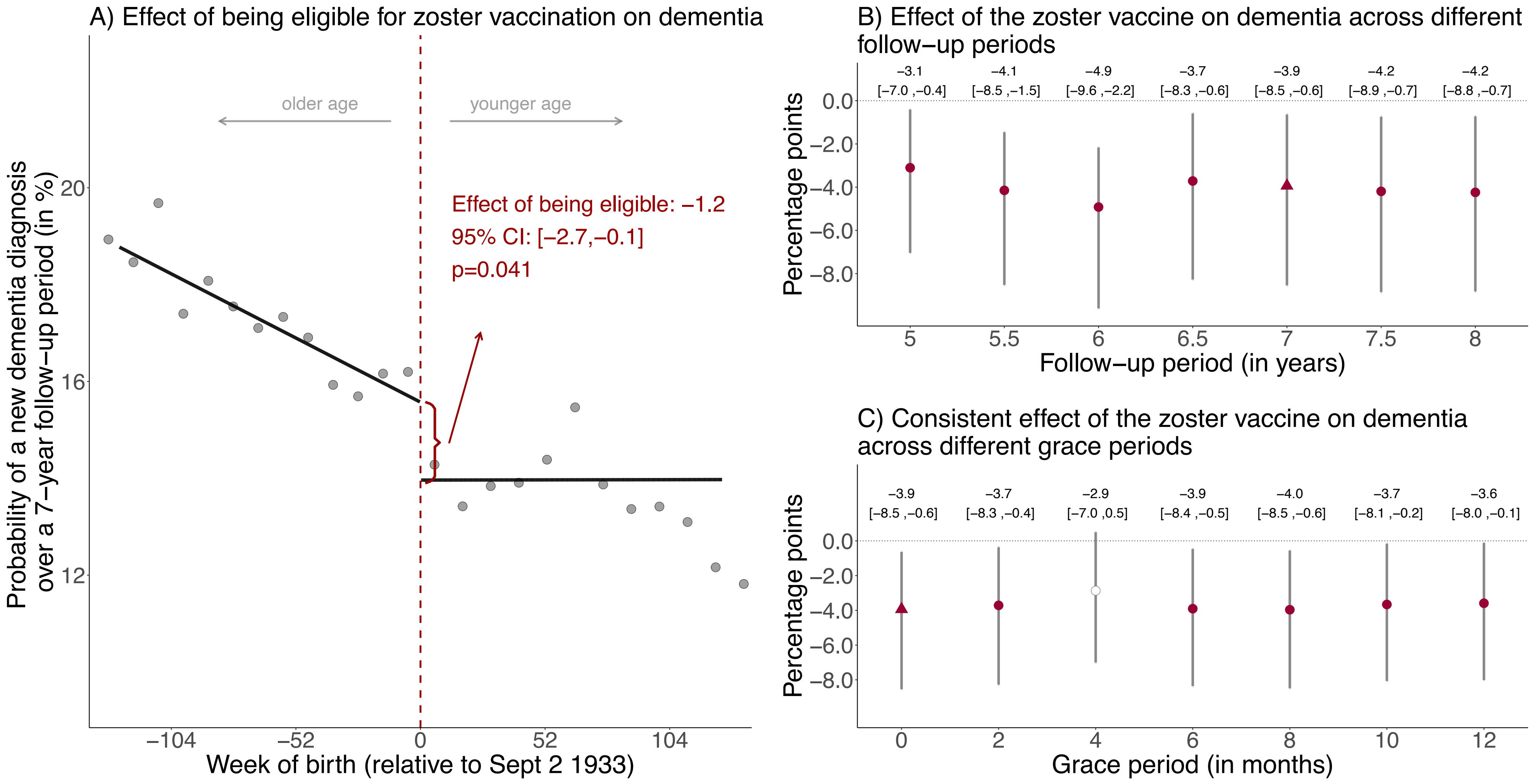

**Fig. S13**: Effect estimates of being eligible (A) and having received the zoster vaccine (B and C) on new diagnoses of dementia, taking into account the staggered roll-out and controlling for cohort fixed effects.[1,2,3,4,5,6]

[1] Instead of starting the follow-up period for all individuals on September 1 2013, we adjusted the follow-up period to account for the staggered rollout of the program by beginning the follow-up period for each individual on the date on which they first became eligible for the zoster vaccine (see Methods for details). We added cohort fixed effects to these analyses to control for between-cohort differences of the date at which the follow-up window started. That is, we defined one cohort fixed effect for ineligible individuals and the first catch-up cohort and included additional cohort fixed effects for each group of patients who became eligible at the same time.

[2] Triangles (rather than points) depict our primary specification.

[3] Red (as opposed to white) fillings denote statistical significance ($p<0.05$).

[4] With "grace periods" we refer to time periods since the index date after which follow-up time is considered to begin to allow for the time needed for a full immune response to develop after vaccine administration.

[5] Grey vertical bars depict 95% confidence intervals.

[6] Grey dots in Panel A show the mean value for each 10-week increment in week of birth.

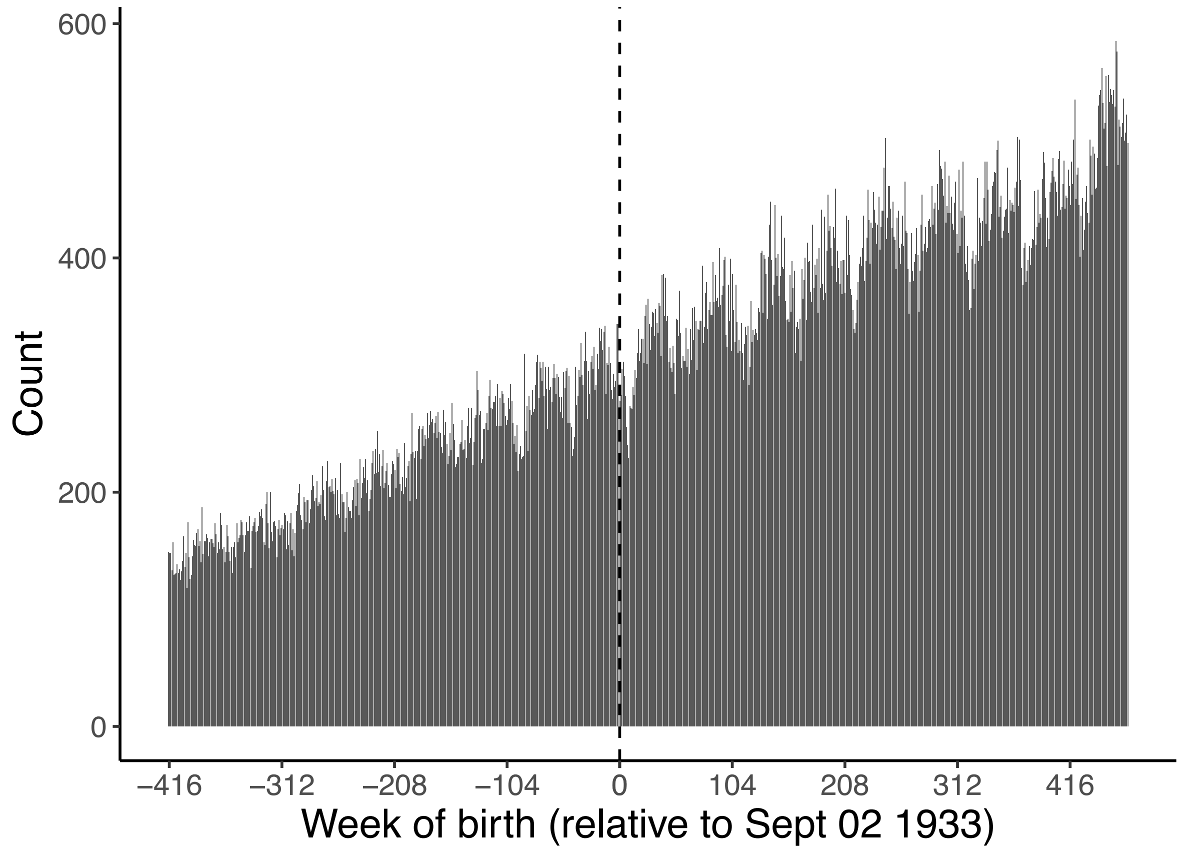

**Fig. S14**: Histogram showing the number of individuals in our dataset by week of birth.[1]

[1] As detailed in the Methods, individuals in week 0 were excluded from our dataset because it was not possible to determine whether they fell above or below the date-of-birth eligibility threshold for the zoster vaccine program.

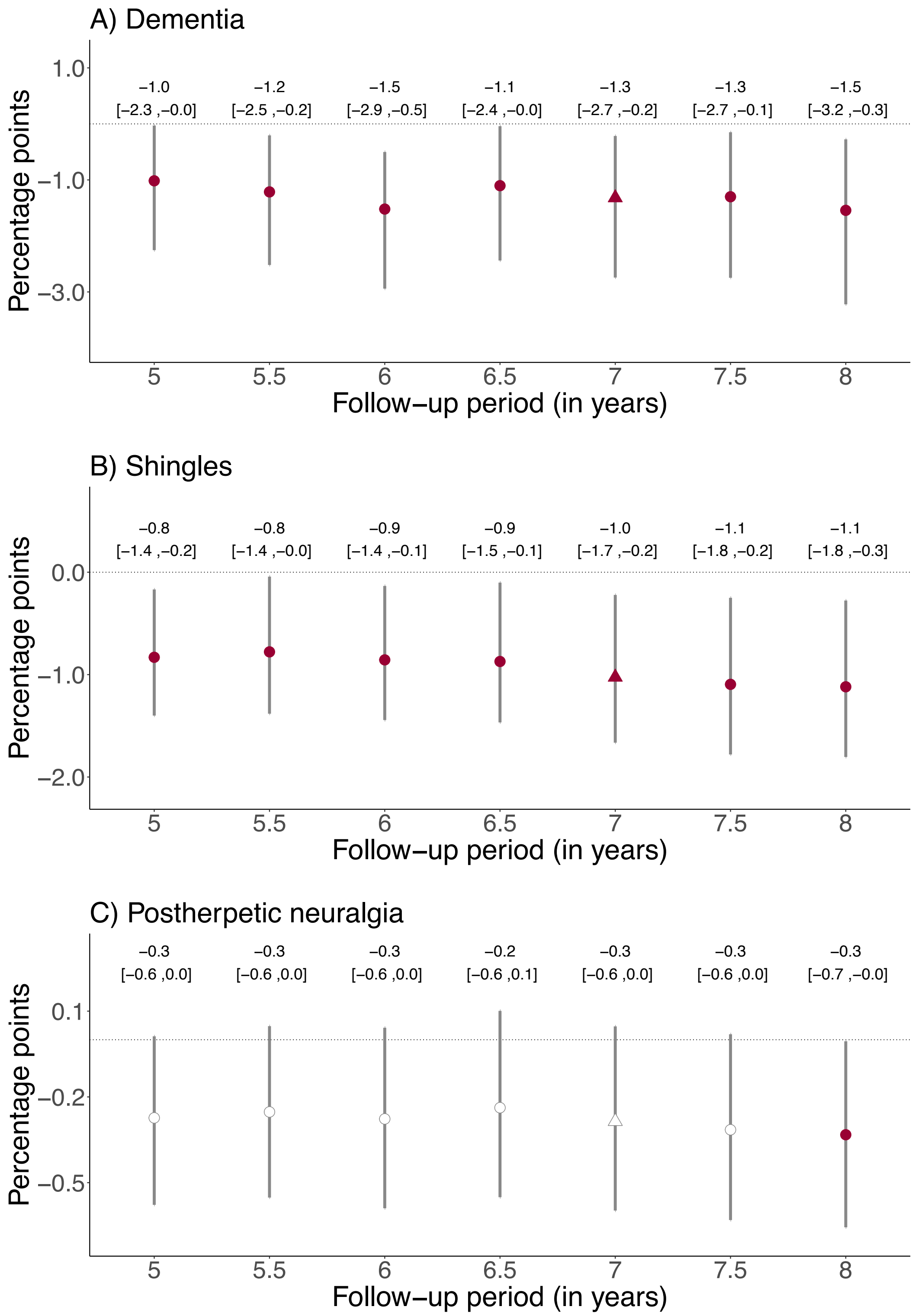

**Fig. S15**: Effect estimates of being eligible for the zoster vaccine on the probability of a new dementia diagnosis, having at least one shingles diagnosis, and having at least one diagnosis of postherpetic neuralgia across various follow-up periods.[1,2,3,4]

[1] We show the same plots for the effect estimates of receipt of the zoster vaccine (as opposed to eligibility for the vaccine) in the main manuscript.

[2] White points depict statistically insignificant point estimates (p>0.05).

[3] Grey vertical bars depict 95% confidence intervals.

[4] Triangles (rather than points) depict our primary specification.

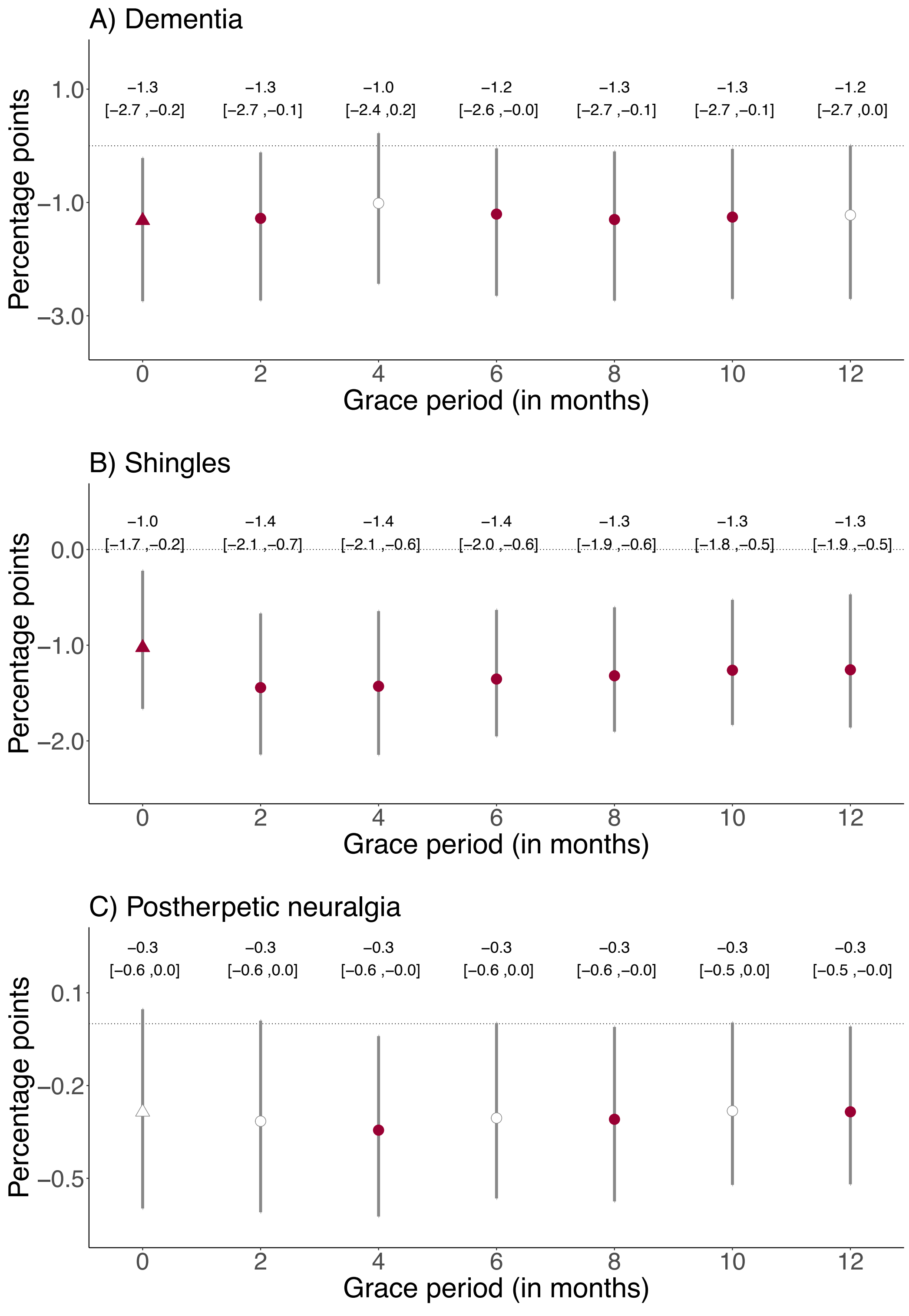

**Fig. S16**: Effect estimates of being eligible for the zoster vaccine on the probability of a new dementia diagnosis, having at least one shingles diagnosis, and having at least one diagnosis of postherpetic neuralgia across various grace periods.[1,2,3,4,5]

[1] With "grace periods" we refer to time periods since the index date after which follow-up time is considered to begin to allow for the time needed for a full immune response to develop after vaccine administration.

[2] We show the same plots for the effect estimates of receipt of the zoster vaccine (as opposed to eligibility for the vaccine) in the main manuscript.

[3] White points depict statistically insignificant point estimates (p>0.05).

[4] Grey vertical bars depict 95% confidence intervals.

[5] Triangles (rather than points) depict our primary specification.

#### Tables S1 to S4

| Variable | % | N |
|---|---|---|
| Decile of Welsh Index of Multiple Deprivation, mean (SD) | 5.8 (2.8) | |
| 1 (most deprived) | 7.9 | 22,242 |
| 2 | 9.0 | 25,420 |
| 3 | 9.2 | 26,079 |
| 4 | 9.8 | 27,741 |
| 5 | 10.7 | 30,356 |
| 6 | 10.8 | 30,555 |
| 7 | 10.0 | 28,205 |
| 8 | 10.5 | 29,697 |
| 9 | 10.6 | 29,875 |
| 10 (least deprived) | 11.5 | 32,347 |
| Gender | | |
| Male | 45.4 | 128,322 |
| Female | 54.6 | 154,218 |
| Clinical diagnoses | | |
| Past shingles | 12.2 | 34,540 |
| Ischemic heart disease | 16.3 | 46,025 |
| Chronic obstructive pulmonary disease | 11.9 | 33,511 |
| Past stroke | 7.6 | 21,454 |
| Past lower respiratory tract infection | 51.8 | 146,391 |
| History of lung cancer | 0.5 | 1,402 |
| Past fall(s) | 19.0 | 53,596 |
| History of colorectal cancer | 2.2 | 6,318 |
| History of lower back pain | 45.1 | 127,518 |
| Diabetes mellitus | 19.7 | 55,666 |
| Uptake of preventive health measures | | |
| Pneumococcal vaccine (PPV-23) | 70.7 | 199,841 |
| Recent statin use | 43.6 | 123,200 |
| Recent antihypertensive use | 59.1 | 167,093 |

**Table S1: Baseline characteristics of the cohort of individuals in our primary analyses for dementia** (n=282,541).[1,2,3,4,5]

[1] The baseline date was September 1 2013.

[2] Recent use of statins and antihypertensive drugs was defined as having received a prescription (whether new or repeat) of these medications within three months prior to September 1 2013.

[3] The clinical codes for all diagnoses are shown in Text S1.

[4] Deciles of the Welsh Index of Multiple Deprivation (WIMD) were calculated based on the 2011 WIMD survey[2].

[5] 24 and 1 individuals had missing information for the WIMD and gender, respectively.

| | Type of dementia | | | |
|---|---|---|---|---|
| | Any type | Alzheimer's | Vascular | Unspecified |
| | (1) | (2) | (3) | (4) |
| CACE (% points) | -3.5 | -1.3 | -1.1 | -1.0 |
| 95% CI | (-7.1, -0.6) | (-3.8, 0.8) | (-3.2, 0.7) | (-3.1,1.0) |
| p | 0.019 | 0.189 | 0.212 | 0.336 |
| Relative effect (%) | -19.9 | -17.9 | -18.8 | -19.1 |
| Bandwidth (in weeks) | 90.6 | 87.4 | 89.3 | 84.2 |
| Observations | 56,098 | 54,205 | 55,139 | 51,985 |
| ITT (% points) | -1.3 | -0.6 | -0.3 | -0.5 |
| 95% CI | (-2.7, -0.2) | (-1.7, 0.2) | (-1.2, 0.4) | (-1.3, 0.3) |
| p | 0.022 | 0.11 | 0.36 | 0.193 |
| Relative effect (%) | -8.5 | -9.9 | -6.1 | -9.8 |
| Bandwidth (in weeks) | 134.4 | 108.1 | 115.3 | 132.0 |
| Observations | 83,167 | 66,933 | 71,440 | 81,484 |

**Table S2: Effect estimates of being eligible and having received the zoster vaccine on new diagnoses of dementia, by type of dementia.** The CACE (complier average causal effect) refers to the estimated effect of actually receiving the zoster vaccine rather than merely being eligible for the vaccine. The ITT (intent-to-treat) effect refers to the estimated effect of being eligible for the zoster vaccine. The bandwidth is the window (in weeks) in participants' date of birth that is drawn around the September 2 1933 eligibility threshold. We used mean-squared error (MSE) optimal bandwidths[3]. Observations refer to the numbers of observations within the MSE-optimal bandwidth.

Abbreviations: CACE=complier average causal effect (i.e., the estimated effect of actually receiving the zoster vaccine rather than merely being eligible for the vaccine); ITT=intent-to-treat effect (i.e., the estimated effect of being eligible for the zoster vaccine); CI=robust bias-corrected confidence interval; p=p value

| | Type of dementia | | | | | |
|---|---|---|---|---|---|---|
| | Any type | Alzheimer's | Vascular | Unspecified | Shingles | Postherpetic neuralgia |
| | (1) | (2) | (3) | (4) | (5) | (6) |
| Difference in CACE by gender | 6.8 | 4.7 | 1.5 | 1.7 | 0.7 | -0.4 |
| 95% CI | (1.2, 12.4) | (0.8, 8.5) | (-1.9, 5.0) | (-1.9, 5.2) | (-2.2, 3.7) | (-1.6, 0.9) |
| p | 0.018 | 0.018 | 0.376 | 0.358 | 0.626 | 0.564 |
| Bandwidth (in weeks) | 90.6 | 87.4 | 89.3 | 84.2 | 116.9 | 96.6 |
| Observations | 56,098 | 54,205 | 55,139 | 51,985 | 76,316 | 63,039 |

**Table S3: Difference (in percentage points) between women and men in the effect of receipt of the zoster vaccine on new diagnoses of dementia, having at least one shingles diagnosis, and having at least one diagnosis of postherpetic neuralgia.** The CACE (complier average causal effect) refers to the estimated effect of actually receiving the zoster vaccine rather than merely being eligible for the vaccine. The reference group is women when calculating the difference in CACE. The difference in CACE by gender is estimated by running the following instrumental variable model:

$$Y_i = \alpha + \beta_1 V_i + \beta_2 \cdot (WOB_i - c_0) + \beta_3 D_i \cdot (WOB_i - c_0) + \beta_4 V_i \cdot MALE_i +$$
$$\beta_5 \cdot (WOB_i - c_0) \cdot MALE_i + \beta_6 D_i \cdot (WOB_i - c_0) \cdot MALE_i + \epsilon_i$$

where i indexes the individual. Y is a binary variable equal to one if an individual experienced the outcome (i.e. a new dementia diagnosis, at least one shingles diagnosis, or at least one postherpetic neuralgia diagnosis). The binary variable V indicates receipt of the zoster vaccine. The binary variable D indicates eligibility for the zoster vaccine (it is, thus, equal to one if an individual was born on or after the cutoff date of September 2 1933). The binary variable MALE is equal to one if an individual is recorded as male in the electronic health record data. The term (WOB-$C_0$) indicates an individual's week of birth centered around the September 2 1933 eligibility date. The interaction term D*(WOB-$C_0$) allows for the slope of the regression line to differ on either side of the date-of-birth eligibility threshold. Adding the terms (WOB-$C_0$)*MALE and D*(WOB-$C_0$)*MALE allows the slopes to vary by gender. V and V*MALE are instrumented by D and D*MALE using two-stage least squares regression. The parameter $\beta_4$ identifies the gender difference in the effect of receipt of the vaccine on the outcome; this is the parameter reported in this table. Consistent with our primary regression discontinuity models shown in the main manuscript, we used local linear triangular kernel regressions and the MSE-optimal bandwidth sizes from the respective regression model (without interaction terms by gender) in our primary analysis. Observations refer to the number of observations within the optimal bandwidth.

Abbreviations: CACE=complier average causal effect (i.e., the estimated effect of actually receiving the zoster vaccine rather than merely being eligible for the vaccine); CI=confidence interval; p=p value

| | Main | Alternative definition of dementia | | Restricted to frequent GP visitors | Controlled for health service utilization | Accounting for staggered roll-out | Quadratic fit |
|---|---|---|---|---|---|---|---|
| | | by cause of death | by prescription | | | | |
| | (1) | (2) | (3) | (4) | (5) | (6) | (7) |
| CACE | -3.5 | -1.6 | -1.9 | -3.0 | -3.3 | -3.9 | -3.8 |
| 95% CI | (-7.1, -0.6) | (-3.7, 0.2) | (-4.2, -0.1) | (-6.7, -0.2) | (-6.9, -0.5) | (-8.5, -0.6) | (-7.5, -0.8) |
| p | 0.019 | 0.073 | 0.044 | 0.036 | 0.023 | 0.023 | 0.015 |
| ITT | -1.3 | -0.8 | -0.8 | -1.2 | -1.2 | -1.2 | -1.8 |
| 95% CI | (-2.7, -0.2) | (-1.8, -0.1) | (-1.7, -0.1) | (-2.8, 0.1) | (-2.6, -0.1) | (-2.7, -0.1) | (-3.6, -0.3) |
| p | 0.022 | 0.031 | 0.02 | 0.059 | 0.028 | 0.041 | 0.017 |
| N | 282,541 | 282,541 | 282,541 | 237,196 | 282,541 | 263,475 | 282,541 |

**Table S4: Additional robustness checks**. The CACE (complier average causal effect) refers to the estimated effect of actually receiving the zoster vaccine rather than merely being eligible for the vaccine. The ITT (intent-to-treat) effect refers to the estimated effect of being eligible for the zoster vaccine.

In (1), we show the results from our primary analysis for comparison (i.e., the identical results as shown in Fig. 3 in the main manuscript). In each of the other columns, we implemented the identical analysis as for our primary analysis, except for the following differences.

In (2), we defined dementia solely as dementia being named a primary or contributory cause of death on the death certificate.

In (3), we defined dementia solely as a new prescription of donepezil hydrochloride, galantamine, rivastigmine, memantine hydrochloride, or idebenone.

In (4), we restricted the analysis cohort to only those individuals who had visited their primary care provider at least once in each of the five years preceding the start date of the zoster vaccine program.

In (5), we adjusted our regressions for the following indicators of health service utilization during the follow-up period: the probability of receiving at least one influenza vaccination and the number of i) primary care visits, ii) outpatient visits, and iii) hospital admissions.

In (6), instead of using September 1 2013 as the index date, we set the index date as the date when each date-of-birth cohort first became eligible for the zoster vaccine (see Methods for details) and adjusted our regressions for cohort fixed effects. These results are also shown in Fig. S8.

In (7), we used local squared regression instead of local linear regression. These results are also shown in Fig. S10.

Abbreviations: CACE=complier average causal effect (i.e., the estimated effect of actually receiving the zoster vaccine rather than merely being eligible for the vaccine); CI=robust bias-corrected confidence interval; p=p value; ITT=intent-to-treat effect (i.e., the estimated effect of being eligible for the zoster vaccine); N=sample size; GP=general practitioner

#### References

1. Calonico, S., Cattaneo, M. D. & Titiunik, R. Robust Nonparametric Confidence Intervals for Regression-Discontinuity Designs. *Econometrica* **82**, 2295–2326 (2014).
2. Statistical Directorate. *Welsh Index of Multiple Deprivation 2011: Summary Report*. https://www.gov.wales/welsh-index-multiple-deprivation-full-index-update-ranks-2011 (2011).
3. Imbens, G. & Kalyanaraman, K. Optimal Bandwidth Choice for the Regression Discontinuity Estimator. *The Review of Economic Studies* **79**, 933–959 (2012).
