## Supplement Materials 2 for "Causal evidence that herpes zoster vaccination prevents a proportion of dementia cases"

This file contains Read version 2 (used for all care processes in primary care electronic health record data) and ICD-10 (used for diagnoses in secondary care records and death certificates) codes for diabetes mellitus and the use of antihypertensive medications.

### Diabetes mellitus

#### *Read codes*

| | | | | | | | | | |
|---|---|---|---|---|---|---|---|---|---|
| C10.. | C100. | C1000 | | | C10001 | C100011 | C1001 | C10011 | C100112 |
| C10012 | C100z | C101. | C1010 | C1011 | C101y | C101z | C102. | C1020 | C1021 |
| C102z | C103. | C1030 | C1031 | C103y | C103z | C104. | C104.11 | C1040 | C1041 |
| C104y | C104z | C105. | C1050 | C1051 | C105y | C105z | C106. | C106.12 | C106.2 |
| C1060 | C1061 | C106y | C106z | C107. | C107.12 | C1070 | C1071 | C1072 | C1073 |
| C1074 | C107z | C108. | C108.1 | C108.12 | C108.2 | C1080 | C1081 | C1082 | C1083 |
| C1084 | C1085 | C108512 | C1086 | C1087 | C1088 | C108812 | C1089 | C108A | C108B |
| C108C | C108D | C108E | C108F | C108F12 | C108G | C108H | C108J | C108y | C108z |
| C109. | C109.1 | C109.11 | C109.12 | C109.13 | C109.2 | C109.3 | C1090 | C1091 | C1092 |
| C1093 | C1094 | C1095 | C1096 | C1097 | C1098 | C1099 | C109A | C109B | C109C |
| C109D | C109E | C109F | C109G | C109H | C109J | C109K | C10A | C10A. | |
| C10A0 | C10A1 | C10A4 | C10AW | C10AX | C10B. | | C10B0 | | C10C. |
| C10D. | C10E. | C10E.12 | C10E.2 | C10E0 | C10E1 | C10E2 | C10E3 | C10E4 | C10E5 |
| C10E6 | C10E7 | C10E8 | C10E9 | C10EA | C10EB | C10EC | C10ED | C10EE | C10EF |
| C10EG | C10EH | C10EJ | C10EK | C10EL | C10EM | C10EN | C10EP | C10EQ | C10ER |
| C10F. | C10F.1 | C10F.11 | C10F0 | C10F1 | C10F2 | C10F3 | C10F4 | C10F5 | C10F6 |
| C10F7 | C10F8 | C10F9 | C10FA | C10FB | C10FC | C10FD | C10FE | C10FF | C10FG |
| C10FH | C10FJ | C10FK | C10FL | C10FM | C10FN | C10FP | C10FQ | C10FR | C10FS |
| C10G. | C10G0 | C10H. | C10J. | C10J0 | C10K. | C10K0 | C10L. | C10M. | C10N. |
| C10N0 | C10N1 | C10P. | C10P0 | C10P1 | C10P111 | C10Q. | C10y. | C10y0 | C10y1 |
| C10yy | C10yz | C10z. | C10z0 | C10z1 | C10zy | C10zz | f11.. | f121. | f122. |
| f125. | f126. | f127. | f128. | f129. | f12a. | f12A. | f12b. | f12B. | f12C. |
| f12D. | f12e. | f12E. | f12g. | f12H. | f12I. | f12J. | f12K. | f12M. | f12N. |
| f12P. | f12Q. | f12s. | f12S. | f12T. | f12U. | f12v. | f12V. | f12W. | f12X. |
| f131. | f132. | f133. | f134. | f135. | f136. | f137. | f138. | f139. | f14.. |
| f141. | f142. | f143. | f144. | f145. | f146. | f14w. | f14x. | f14y. | f14z. |
| f151. | f152. | f153. | f154. | f155. | f15x. | f15y. | f15z. | f21.. | f211. |
| f22.. | f221. | f222. | f223. | f226. | f227. | f228. | f231. | f241. | f242. |
| f25.. | f251. | f252. | f253. | f259. | f25a. | f25A. | f25B. | f25C. | f25D. |
| f25E. | f25F. | f25G. | f25H. | f25i. | f25l. | f25m. | f25n. | f25o. | f25p. |
| f25q. | f25r. | f25s. | f25t. | f25u. | f25v. | f25w. | f25W. | f25x. | f25X. |
| f25y. | f25Y. | f25z. | f26.. | f261. | f262. | f27.. | f272. | f27a. | f27A. |
| f27b. | f27B. | f27c. | f27C. | f27d. | f27D. | f27e. | f27E. | f27f. | f27F. |
| f27g. | f27h. | f27H. | f27I. | f27j. | f27J. | f27k. | f27K. | f27l. | f27L. |
| f27m. | f27M. | f27n. | f27N. | f27o. | f27O. | f27p. | f27P. | f27Q. | f27R. |
| f27S. | f27T. | f27u. | f27v. | f27V. | f27w. | f27W. | f27x. | f27X. | f27y. |
| f27Y. | f27z. | f27Z. | f281. | f282. | f283. | f284. | f285. | f286. | f287. |
| f288. | f289. | f28A. | f28B. | f28C. | f28D. | f28E. | f28F. | f291. | f292. |
| f293. | f294. | f295. | f296. | f297. | f298. | f299. | f29A. | f29B. | f29C. |
| f2A1. | f2A2. | f2A3. | f2Ay. | f2Az. | f2B1. | f2B2. | f2B3. | f2B4. | f2B5. |

| | | | | | | | | | |
|---|---|---|---|---|---|---|---|---|---|
| f2B6. | f2C1. | f2C2. | f321. | f322. | f323. | f324. | f33.. | f331. | f332. |
| f333. | f334. | f335. | f336. | f339. | f33d. | f33e. | f33f. | f33g. | f35.. |
| f351. | f352. | f354. | f355. | f357. | f359. | f35C. | f35D. | f35w. | f35x. |
| f35y. | f35z. | f361. | f362. | f363. | f364. | f36z. | f371. | f37z. | f391. |
| f392. | f39y. | f39z. | f3a1. | f3A1. | f3A2. | f3A3. | f3a4. | f3A4. | f3A5. |
| f3A6. | f3A7. | f3A8. | f41.. | f411. | f412. | f413. | f415. | f416. | f417. |
| f418. | f419. | f41A. | f41B. | f41C. | f41D. | f41E. | f41F. | f41G. | f41H. |
| f41I. | f41J. | f41s. | f41t. | f41u. | f41v. | f41w. | f41x. | f41y. | f41z. |
| f512. | f51y. | f71.. | f711. | f71z. | f721. | f722. | f723. | f724. | f725. |
| f72w. | f72x. | f72y. | f72z. | f73.. | f731. | f735. | f736. | f73z. | ft... |
| ft11. | ft12. | ft13. | ft14. | ft21. | ft23. | ft24. | ft31. | ft32. | ft33. |
| ft34. | ft35. | ft36. | ft37. | ft38. | ft39. | ft41. | ft42. | ft43. | ft44. |
| ft45. | ft46. | ft4u. | ft4v. | ft4w. | ft4x. | ft4y. | ft4z. | ft5.. | ft51. |
| ft52. | ft53. | ft55. | ft5x. | ft5y. | ft5z. | ft61. | ft62. | ft63. | ft6x. |
| ft6y. | ft6z. | ft7.. | ft71. | ft7z. | ft8.. | ft81. | ft82. | ft83. | ft8x. |
| ft8y. | ft8z. | ft9.. | ft91. | ft92. | ft93. | ft94. | ft95. | ft9x. | ft9y. |
| ft9z. | fta1. | ftaZ. | ftb1. | ftb2. | ftby. | ftbz. | ftc1. | ftc2. | ftd1. |
| ftd2. | ftdy. | ftdz. | fte1. | ftez. | ftf1. | ftf2. | ftg1. | ftg2. | ftg3. |
| ftg4. | fth1. | fth2. | fth3. | fth4. | fti1. | fti2. | fti3. | fti4. | ftj1. |
| ftj2. | ftj3. | ftj4. | ftj5. | ftj6. | ftk1. | ftk2. | ftk3. | ftk4. | ftk5. |
| ftk6. | ftl1. | ftl2. | ftm1. | ftm2. | ftm3. | ftm4. | ftn1. | ftn2. | ftn3. |
| ftn4. | fto1. | fto2. | fto3. | fto4. | ftp2. | ftp3. | ftp4. | ftq1. | ftq2. |
| ftq3. | ftq4. | ftq5. | ftq7. | ftr2. | ftr3. | ftr4. | ftr5. | ftr6. | ftr7. |
| ftr8. | fw1.. | fw11. | fw12. | fw13. | fw14. | fw15. | fw16. | fw2.. | fw21. |
| fw22. | iz16. | p453. | p454. | p455. | ph... | ph1.. | ph18. | ph21. | ph22. |
| ph23. | ph25. | ph26. | ph29. | ph31. | ph32. | ph33. | ph34. | ph35. | ph36. |
| ph37. | ph38. | ph39. | ph3a. | ph3A. | ph3b. | ph3B. | ph3c. | ph3C. | ph3d. |
| ph3D. | ph3E. | ph3f. | ph3F. | ph3g. | ph3J. | ph3K. | ph3L. | ph3M. | ph3N. |
| ph3P. | ph3Q. | ph3R. | ph3T. | ph3U. | ph3V. | ph3Y. | ph3Z. | ph43. | ph44. |
| ph51. | ph61. | ph62. | ph7.. | ph71. | ph72. | ph73. | ph74. | ph75. | ph76. |
| ph77. | ph78. | ph79. | ph7a. | ph7A. | ph7b. | ph7B. | ph7c. | ph7d. | ph7D. |
| ph7e. | ph7E. | ph7f. | ph7F. | ph7G. | ph7H. | ph7i. | ph7I. | ph7j. | ph7J. |
| ph7k. | ph7K. | ph7l. | ph7L. | ph7m. | ph7M. | ph7n. | ph7N. | ph7o. | ph7O. |
| ph7p. | ph7P. | ph7q. | ph7Q. | ph7R. | ph7s. | ph7S. | ph7t. | ph7T. | ph7u. |
| ph7U. | ph7V. | ph7w. | ph7W. | ph7X. | ph7y. | ph7Y. | ph7z. | ph7Z. | ph81. |
| ph82. | ph83. | ph84. | ph86. | ph87. | ph88. | ph89. | ph8a. | ph8A. | ph8b. |
| ph8B. | ph8c. | ph8C. | ph8d. | ph8D. | ph8e. | ph8E. | ph8f. | ph8F. | ph8g. |
| ph8G. | ph8h. | ph8H. | ph8i. | ph8I. | ph8j. | ph8J. | ph8k. | ph8l. | ph8m. |
| ph8n. | ph8N. | ph8o. | ph8O. | ph8P. | ph8Q. | ph8r. | ph8R. | ph8s. | ph8S. |
| ph8t. | ph8T. | ph8u. | ph8U. | ph8v. | ph8V. | ph8w. | ph8Y. | ph8z. | ph8Z. |
| ph91. | ph92. | ph93. | ph94. | ph95. | ph96. | ph97. | ph98. | ph99. | ph9a. |
| ph9A. | ph9B. | ph9c. | ph9C. | ph9d. | ph9D. | ph9E. | ph9g. | ph9h. | ph9i. |
| ph9I. | ph9j. | ph9J. | ph9k. | ph9K. | ph9l. | ph9L. | ph9m. | ph9M. | ph9N. |
| ph9O. | ph9p. | ph9P. | ph9q. | ph9Q. | ph9r. | ph9R. | ph9s. | ph9S. | ph9t. |
| ph9T. | ph9u. | ph9U. | ph9V. | ph9W. | ph9X. | ph9Y. | ph9Z. | phB1. | phB2. |
| phB3. | phB4. | phB5. | phB6. | phB7. | phB8. | phB9. | phBA. | phC3. | phC4. |
| phC5. | phC6. | phC7. | phC8. | phC9. | phCa. | phCA. | phCB. | phCC. | phCD. |
| phCE. | phCF. | phCG. | phCH. | phCI. | phCJ. | phCK. | phCL. | phCM. | phCN. |
| phCO. | phCP. | phCQ. | phCR. | phCS. | phCT. | phCU. | phD1. | phD2. | phD3. |
| phD4. | phD5. | phD6. | pm16. | pm17. | pm18. | pm19. | pm1a. | pm1A. | pm1b. |
| pm1B. | pm1c. | pm1C. | pm1d. | pm1D. | pm1e. | pm1E. | pm1f. | pm1F. | pm1G. |

| | | | | | | | | | |
|---|---|---|---|---|---|---|---|---|---|
| pm1h. | pm1H. | pm1i. | pm1I. | pm1j. | pm1J. | pm1k. | pm1K. | pm1L. | pm1m. |
| pm1M. | pm1n. | pm1N. | pm1o. | pm1p. | pm1P. | pm1q. | pm1Q. | pm1r. | pm1R. |
| pm1s. | pm1S. | pm1t. | pm1T. | pm1u. | pm1U. | pm1v. | pm1V. | pm1W. | pm1x. |
| pm1X. | pm1y. | pm1Y. | pm1z. | pm1Z. | pm21. | pm22. | pm23. | pm24. | pm25. |
| pm26. | pm27. | pm28. | pm29. | pm2A. | pm2B. | pm2C. | pm2D. | pm2E. | pm2F. |
| pm2G. | pm2H. | pm2I. | pm2J. | pm2K. | pm2L. | pm2M. | pu1.. | pu11. | pu13. |
| pu21. | pu31. | pu41. | pu42. | pu51. | pu52. | pu53. | pu55. | pu56. | pu61. |
| pu71. | pu72. | pu74. | pu75. | puh.. | puh1. | puh2. | puh3. | puh4. | puh5. |
| puh6. | puh7. | puh8. | puh9. | puha. | puhb. | puhc. | puhe. | puhE. | puhf. |
| puhF. | puhg. | puhG. | puhh. | puhH. | puhi. | puhI. | puhj. | puhJ. | puhk. |
| puhK. | puhl. | puhL. | puhm. | puhM. | puhn. | puhN. | puhO. | puhp. | puhP. |
| puhq. | puhQ. | puhr. | puhR. | puhs. | puhS. | puht. | puhT. | puhu. | puhU. |
| puhV. | puhw. | puhx. | puhy. | puhY. | puhZ. | pui1. | pui8. | puig. | puiG. |
| puiI. | puiM. | puiN. | puiR. | puit. | puiX. | puj.. | puj5. | puj8. | pujE. |
| pujF. | pujH. | pujI. | pujL. | pujp. | pun.. | pun1. | pun2. | pun3. | puq7. |
| puqK. | pur.. | pur1. | pur2. | pur3. | pur4. | pur5. | pur6. | pur7. | pur8. |
| pur9. | purA. | purB. | purC. | purD. | purE. | purF. | purG. | purH. | purI. |
| purJ. | purK. | purM. | purN. | purO. | purP. | purQ. | purR. | purS. | purT. |
| purU. | purY. | | | | | | | | |

***ICD-10 codes***

| | | | | | | | | | |
|---|---|---|---|---|---|---|---|---|---|
| E10 | E10- | E10. | E100 | E101 | E102 | E103 | E104 | E105 | E106 |
| E107 | E108 | E109 | E10X | E11 | E11- | E11, | E11. | E110 | E111 |
| E112 | E113 | E114 | E115 | E116 | E117 | E118 | E119 | E11X | E12 |
| E120 | E121 | E122 | E123 | E124 | E126 | E127 | E128 | E129 | E130 |
| E131 | E132 | E133 | E134 | E135 | E136 | E137 | E138 | E139 | E13X |
| E14 | E14. | E140 | E141 | E142 | E143 | E144 | E145 | E146 | E147 |
| E148 | E149 | E14X | | | | | | | |

### Antihypertensive medications

#### *Read codes*

| | | | | | | | | | |
|---|---|---|---|---|---|---|---|---|---|
| b21.. | b211. | b212. | b213. | b214. | b215. | b216. | b217. | b218. | b219. |
| b21A. | b21B. | b22.. | b221. | b22y. | b22z. | b23.. | b231. | b232. | b23y. |
| b23z. | b251. | b25z. | b26.. | b261. | b262. | b263. | b264. | b26y. | b26z. |
| b271. | b27z. | b28.. | b281. | b282. | b283. | b285. | b286. | b287. | b288. |
| b28z. | b291. | b29z. | b2a1. | b2az. | b2b.. | b2b1. | b2b2. | b2b3. | b2bz. |
| b2c1. | b2cz. | b2d1. | b2dz. | bb3.. | bb31. | bb32. | bb33. | bb34. | bb35. |
| bb36. | bb37. | bb38. | bb39. | bb3a. | bb3A. | bb3b. | bb3B. | bb3c. | bb3C. |
| bb3d. | bb3D. | bb3e. | bb3f. | bb3F. | bb3g. | bb3h. | bb3i. | bb3j. | bb3k. |
| bb3K. | bb3l. | bb3L. | bb3m. | bb3M. | bb3n. | bb3N. | bb3o. | bb3P. | bb3Q. |
| bb3t. | bb3v. | bb3w. | bb3y. | bb3z. | bd... | bd1.. | bd11. | bd12. | bd13. |
| bd14. | bd15. | bd16. | bd19. | bd1a. | bd1A. | bd1B. | bd1C. | bd1d. | bd1D. |
| bd1E. | bd1F. | bd1g. | bd1G. | bd1h. | bd1H. | bd1i. | bd1I. | bd1j. | bd1J. |
| bd1k. | bd1K. | bd1l. | bd1L. | bd1m. | bd1n. | bd1N. | bd1o. | bd1p. | bd1P. |
| bd1q. | bd1Q. | bd1r. | bd1R. | bd1s. | bd1S. | bd1U. | bd1W. | bd1x. | bd1X. |
| bd1y. | bd1Y. | bd1z. | bd1Z. | bd21. | bd22. | bd23. | bd2w. | bd2x. | bd2y. |
| bd3.. | bd31. | bd32. | bd33. | bd34. | bd35. | bd36. | bd38. | bd3a. | bd3b. |
| bd3c. | bd3f. | bd3g. | bd3h. | bd3i. | bd3j. | bd3k. | bd3l. | bd3x. | bd3y. |
| bd3z. | bd41. | bd4z. | bd5.. | bd51. | bd52. | bd53. | bd54. | bd57. | bd58. |
| bd59. | bd5a. | bd5b. | bd5t. | bd5u. | bd5v. | bd5y. | bd5z. | bd6.. | bd61. |
| bd62. | bd64. | bd65. | bd66. | bd67. | bd6b. | bd6c. | bd6w. | bd6x. | bd6y. |
| bd6z. | bd7.. | bd71. | bd72. | bd7y. | bd7z. | bd81. | bd82. | bd83. | bd84. |
| bd8c. | bd8d. | bd8e. | bd8f. | bd8g. | bd8h. | bd8i. | bd8o. | bd8u. | bda1. |
| bda2. | bday. | bdaz. | bdc.. | bdc1. | bdc2. | bdc3. | bdc4. | bdc5. | bdcu. |
| bdcv. | bdcw. | bdcx. | bdcz. | bdd1. | bdd2. | bddz. | bde1. | bde2. | bde3. |
| bde4. | bde5. | bde6. | bde7. | bde8. | bde9. | bdea. | bdeA. | bdeb. | bdeB. |
| bdec. | bdeC. | bded. | bdeD. | bdee. | bdeE. | bdef. | bdeg. | bdeG. | bdeh. |
| bdeH. | bdei. | bdej. | bdeJ. | bdek. | bdeK. | bdel. | bdeL. | bdem. | bdeM. |
| bden. | bdeN. | bdeo. | bdeO. | bdep. | bdeP. | bdeq. | bdeQ. | bder. | bdes. |
| bdet. | bdeu. | bdev. | bdew. | bdex. | bdey. | bdez. | bdf.. | bdf1. | bdf2. |
| bdf3. | bdf4. | bdf5. | bdf6. | bdf7. | bdf8. | bdf9. | bdfA. | bdfB. | bdfC. |
| bdfD. | bdfE. | bdfF. | bdfG. | bdfH. | bdfI. | bdfJ. | bdfL. | bdfM. | bdfw. |
| bdfx. | bdfy. | bdfz. | bdh2. | bdh4. | bdi1. | bdi2. | bdj1. | bdj2. | bdj3. |
| bdj4. | bdj5. | bdl.. | bdl1. | bdl2. | bdl3. | bdl4. | bdl5. | bdl6. | bdl7. |
| bdl8. | bdm.. | bdm1. | bdmy. | bdmz. | bdn1. | bdn2. | bdn3. | bdn4. | bdn5. |
| bdn6. | bh11. | bh12. | bh13. | bh14. | bh1y. | bh1z. | bh2y. | bh2z. | bh31. |
| bh3y. | bh41. | bh42. | bh43. | bh44. | bh45. | bh46. | bh47. | bh48. | bh49. |
| bh4v. | bh4w. | bh4x. | bh4y. | bh4z. | bh51. | bh52. | bh53. | bh54. | bh55. |
| bh56. | bh5y. | bh5z. | bh6.. | bh61. | bh62. | bh63. | bh64. | bh65. | bh66. |
| bh67. | bh68. | bh69. | bh6A. | bh6B. | bh6C. | bh6D. | bh6E. | bh6F. | bh6G. |
| bh6H. | bh6y. | bh6z. | bi... | bi1.. | bi11. | bi13. | bi15. | bi16. | bi17. |
| bi18. | bi19. | bi1a. | bi1b. | bi1d. | bi1f. | bi1F. | bi1g. | bi1G. | bi1H. |
| bi1I. | bi1j. | bi1J. | bi1k. | bi1K. | bi1l. | bi1p. | bi1q. | bi1s. | bi1v. |
| bi1w. | bi1x. | bi1y. | bi1z. | bi2.. | bi21. | bi22. | bi23. | bi24. | bi25. |
| bi26. | bi27. | bi28. | bi29. | bi2a. | bi2A. | bi2b. | bi2B. | bi2C. | bi2D. |
| bi2E. | bi2F. | bi2G. | bi2H. | bi2J. | bi2K. | bi2L. | bi2M. | bi2t. | bi2u. |
| bi2v. | bi2w. | bi2x. | bi2y. | bi2z. | bi3.. | bi31. | bi32. | bi33. | bi34. |
| bi35. | bi36. | bi37. | bi38. | bi39. | bi3a. | bi3b. | bi3c. | bi3d. | bi3e. |

| | | | | | | | | | |
|---|---|---|---|---|---|---|---|---|---|
| bi3f. | bi3g. | bi3h. | bi3i. | bi3j. | bi3k. | bi3l. | bi3m. | bi3n. | bi3p. |
| bi3q. | bi3r. | bi3s. | bi3t. | bi3y. | bi41. | bi42. | bi43. | bi44. | bi45. |
| bi46. | bi47. | bi48. | bi49. | bi4A. | bi4D. | bi5.. | bi51. | bi52. | bi53. |
| bi54. | bi55. | bi56. | bi57. | bi58. | bi6.. | bi61. | bi62. | bi63. | bi64. |
| bi65. | bi66. | bi67. | bi68. | bi69. | bi6A. | bi6B. | bi6C. | bi6D. | bi6E. |
| bi6F. | bi6G. | bi6o. | bi6t. | bi6u. | bi6v. | bi6w. | bi6x. | bi6y. | bi6z. |
| bi71. | bi72. | bi73. | bi74. | bi81. | bi82. | bi83. | bi84. | bi85. | bi86. |
| bi87. | bi88. | bi89. | bi8a. | bi91. | bi92. | bi93. | bi94. | bi95. | bi96. |
| bi9A. | bi9z. | biA1. | biA2. | biA3. | biA4. | biB1. | biB2. | biB3. | biBx. |
| biBy. | biBz. | biC1. | biC2. | biC3. | biC4. | biC5. | biC6. | biC7. | biC8. |
| bk3.. | bk31. | bk32. | bk33. | bk34. | bk35. | bk36. | bk37. | bk38. | bk39. |
| bk3A. | bk3B. | bk3C. | bk3D. | bk3E. | bk3y. | bk3z. | bk4.. | bk41. | bk42. |
| bk43. | bk44. | bk45. | bk46. | bk47. | bk48. | bk49. | bk4A. | bk4B. | bk4s. |
| bk4t. | bk4u. | bk4v. | bk4w. | bk4x. | bk4y. | bk4z. | bk5.. | bk51. | bk52. |
| bk53. | bk54. | bk55. | bk56. | bk57. | bk58. | bk59. | bk5x. | bk5y. | bk5z. |
| bk61. | bk62. | bk7.. | bk71. | bk72. | bk73. | bk74. | bk75. | bk76. | bk77. |
| bk78. | bk79. | bk7z. | bk81. | bk82. | bk83. | bk84. | bk85. | bk86. | bk87. |
| bk88. | bk8w. | bk8x. | bk8y. | bk8z. | bk91. | bk92. | bk93. | bk9x. | bk9y. |
| bk9z. | bkB.. | bkB1. | bkB2. | bkB3. | bkB4. | bkB5. | bkB6. | bkC1. | bkC2. |
| bkC3. | bkCx. | bkCy. | bkCz. | bkD1. | bkD2. | bkD3. | bkDx. | bkDy. | bkDz. |
| bkF1. | bkF2. | bkFy. | bkFz. | bkH1. | bkH2. | bkH3. | bkHx. | bkHy. | bkHz. |
| bkI2. | bkI3. | bkI4. | bkI5. | bkJ1. | bkJ2. | bkJ3. | bkJ4. | bkJ5. | bkJ6. |
| bkL1. | bkL2. | bkL3. | bkL4. | bkL5. | bkL6. | bl5.. | bl51. | bl53. | bl54. |
| bl55. | bl56. | bl57. | bl58. | bl59. | bl5a. | bl5A. | bl5b. | bl5B. | bl5c. |
| bl5C. | bl5d. | bl5D. | bl5e. | bl5E. | bl5f. | bl5F. | bl5g. | bl5G. | bl5h. |
| bl5H. | bl5I. | bl5J. | bl5k. | bl5K. | bl5l. | bl5L. | bl5m. | bl5M. | bl5n. |
| bl5N. | bl5o. | bl5O. | bl5p. | bl5P. | bl5q. | bl5Q. | bl5r. | bl5R. | bl5s. |
| bl5S. | bl5t. | bl5u. | bl5U. | bl5v. | bl5V. | bl5w. | bl5W. | bl5x. | bl5y. |
| bl5Y. | bl5z. | bl5Z. | bl71. | bl72. | bl73. | bl74. | bl7w. | bl7x. | bl7y. |
| bl7z. | bl8.. | bl81. | bl82. | bl83. | bl84. | bl85. | bl86. | bl8A. | bl8b. |
| bl8B. | bl8C. | bl8D. | bl8e. | bl8E. | bl8f. | bl8F. | bl8g. | bl8G. | bl8h. |
| bl8H. | bl8i. | bl8j. | bl8J. | bl8k. | bl8K. | bl8l. | bl8L. | bl8m. | bl8M. |
| bl8n. | bl8o. | bl8O. | bl8p. | bl8P. | bl8q. | bl8r. | bl8R. | bl8S. | bl8t. |
| bl8T. | bl8u. | bl8U. | bl8v. | bl8V. | bl8w. | bl8W. | bl8x. | bl8X. | bl8y. |
| bl8Y. | bl8z. | bl8Z. | bla1. | bla2. | blb.. | blb1. | blb2. | blb3. | blb4. |
| blb5. | blb6. | blc.. | blc1. | blc2. | blc3. | blc4. | blc5. | blc6. | blc7. |
| blc8. | blc9. | blca. | blcb. | blcc. | blcd. | blce. | blch. | blci. | blcj. |
| blck. | blcl. | blcm. | blcn. | blco. | blcp. | blcr. | blcs. | blct. | ble1. |
| ble2. | ble3. | ble4. | blg1. | blg2. | blg3. | blg4. | blg5. | blg6. | blh1. |
| blh2. | blh3. | blh4. | bli1. | bli2. | bli3. | bli4. | blj1. | blj2. | blj3. |
| blj4. | blj5. | blj7. | blj8. | blj9. | blja. | bljA. | bljb. | bljB. | bljC. |
| bljD. | bljE. | bljF. | bljJ. | bljK. | bljL. | bljM. | bljN. | bljO. | bljP. |
| bljQ. | bljR. | bljS. | bljT. | bljU. | bljV. | bljX. | bljY. | bll1. | bll2. |
| bll3. | bll5. | bll6. | bll7. | blla. | bllb. | bllc. | blld. | bllf. | bllg. |
| bllh. | bllk. | blll. | dt13. | dt14. | | | | | |

***ICD-10 codes***

Not applicable
